# Disease trajectories in hospitalized COVID-19 patients are predicted by clinical and peripheral blood signatures representing distinct lung pathologies

**DOI:** 10.1101/2023.09.08.23295024

**Authors:** João Da Silva Filho, Vanessa Herder, Matthew P. Gibbins, Monique Freire dos Reis, Gisely Cardoso Melo, Michael J. Haley, Carla Cristina Judice, Fernando Fonseca Almeida Val, Mayla Borba, Tatyana Almeida Tavella, Vanderson de Sousa Sampaio, Charalampos Attipa, Fiona McMonagle, Marcus Vinicius Guimaraes de Lacerda, Fabio Trindade Maranhão Costa, Kevin N. Couper, Wuelton Marcelo Monteiro, Luiz Carlos de Lima Ferreira, Christopher Alan Moxon, Massimo Palmarini, Matthias Marti

## Abstract

Linking clinical biomarkers and lung pathology still is necessary to understand COVID-19 pathogenesis and the basis of progression to lethal outcomes. Resolving these knowledge gaps enables optimal treatment approaches of severe COVID-19. We present an integrated analysis of longitudinal clinical parameters, blood biomarkers and lung pathology in COVID-19 patients from the Brazilian Amazon. We identified core signatures differentiating severe recovered patients and fatal cases with distinct disease trajectories. Progression to early death was characterized by rapid and intense endothelial and myeloid activation, presence of thrombi, mostly driven by SARS-CoV-2^+^ macrophages. Progression to late death was associated with systemic cytotoxicity, interferon and Th17 signatures and fibrosis, apoptosis, and abundant SARS-CoV-2 ^+^ epithelial cells in the lung. Progression to recovery was associated with pro-lymphogenic and Th2-mediated responses. Integration of ante-mortem clinical and blood biomarkers with post-mortem lung-specific signatures defined predictors of disease progression, identifying potential targets for more precise and effective treatments.

## Introduction

COVID-19, as with many infectious diseases, displays a wide spectrum of clinical outcomes in infected patients. Current understanding of disease pathogenesis indicates that the host responses to SARS-CoV-2, the causative agent of COVID-19, in hospitalized patients with severe disease is highly complex, heterogenous, and temporally dynamic (Del Valle et al., 2020; Kuri-Cervantes et al., 2020; Laing et al., 2020; Lucas et al., 2020; Mathew et al., 2020; Tay et al., 2020).

There is strong evidence that both immune hyperactivation and immunosuppression are involved in disease pathogenesis. Immune hyperactivation is indicated by high levels of inflammatory and tissue damage markers, including C-reactive protein (CRP), ferritin and D-dimer, high neutrophil-to-lymphocyte ratio (NLCR) (Del Valle et al., 2020; Herold et al., 2020; Mehta et al., 2020; Wu et al., 2020) and increased levels of inflammatory cytokines and chemokines (Del Valle et al., 2020; Mehta et al., 2020; Tay et al., 2020; Zhang et al., 2020). In the lung infiltration of proinflammatory myeloid cells has been described, associated with tissue damage (Mann et al., 2020; Moore and June, 2020; Tay et al., 2020). The role of hyperinflammation is supported by improved outcome in clinical trials using glucocorticoids (dexamethasone), and inhibitors of the IL-6 receptor (e.g., tocilizumab) or Janus kinases (JAK, e.g., baricitinib) (Gordon et al., 2021; Group et al., 2021; Kalil et al., 2021). Reciprocally several studies in severe COVID-19 patients demonstrate defective lymphoid responses, associated with lymphopenia, impaired T-cell function, impaired/delayed interferon (IFN) antiviral responses and immune senescence leading to failure to control virus replication. These immunosuppressive responses are also associated with lung tissue damage (Bost et al., 2021; Chen and John Wherry, 2020; Diao et al., 2020). Immune hyper-*vs* hypo responsiveness may be dominant at different phases of the disease in the same patient or may represent distinct disease response trajectories in different patients. These contrasting scenarios of hyper or hypo immune activation emphasize the complex pathophysiology of COVID-19 and challenges the “one size fits all” therapeutic approaches currently available for treatment of severe COVD-19 ( and Consortium, 2022).

Characterizing the specific responses longitudinally in different patients and identifying biomarkers that define the critical processes driving lung disease in a particular patient with a particular disease trajectory would enable us to stratify patients to the best treatments. To do this we need a way to link measurements that can be made from clinical observations and biomarkers in blood to the underlying processes occurring in the lung parenchyma. However, the lung is generally inaccessible during life and to date, in cases that have been characterized post-mortem, there are limited clinical or blood biomarker data during life. Thus, 3 years after the beginning of the pandemic there is still limited data on the direct association between the longitudinal changes in clinical parameters and peripheral blood protein markers and the lung-specific pathological responses driving disease progression (i.e., speed of recovery or death).

Here, we investigated a longitudinal cohort of severe COVID-19 patients, from hospital admission up to outcome (recovery or death), in the Brazilian Amazon during the first wave of the pandemic in whom lung tissue was collected post-mortem in fatal cases. We had two main aims: firstly, to characterize clinical and peripheral blood (PB) biomarkers over the course of acute illness and identify how specific systemic signatures at admission can predict disease progression and outcome; secondly to characterize pathological responses developing in the lung and to link these back to the clinical and PB data to assess what these accessible data during life tell us about the underlying disease processes in the lung. We undertook detailed peripheral blood profiling at multiple time points during hospitalization and linked this with clinical data taken throughout the course of the disease up to hospital discharge (recovered patients) or time of death (fatal cases). This comprehensive longitudinal data was complemented with histopathological analysis and a spatially resolved single-cell atlas of matched post-mortem lung samples (fatal cases). By using machine learning and systems-biology based approaches, we integrated the clinical, peripheral blood, and lung data, which deconvolved the host signatures driving distinct trajectories in COVID-19 disease progression. Crucially, these models defined predictors of the trajectories in disease progression during SARS-CoV-2 infection. To our knowledge, this is the first study linking detailed investigation of serial clinical data and peripheral blood samples taken during life, to detailed histopathological and spatially-resolved single-cell investigation of lung samples in death in any acute respiratory infection.

Thus, our study can contribute to the development of more precise, targeted, and tailored treatment approaches for COVID-19 patients, which might be especially beneficial in health systems with limited resources.

## Results

### Clinical and peripheral blood parameters at hospital admission stratify disease progression of COVID-19 patients

We followed a prospective cohort of 142 hospitalized adult patients admitted at the Delphina Rinaldi Emergency Hospital (HPSDRA), in collaboration with the Tropical Medicine Foundation Dr Heitor Vieira Dourado (FMT-HVD) (Manaus, Brazilian Amazon Region). Patients had confirmed laboratory diagnosis of COVID-19 by RT-PCR testing of nasopharyngeal swab samples (Delafiori et al., 2021; Huang et al., 2020; Oliveira et al., 2022; Qin et al., 2020). This study was conducted during the first wave of the pandemic (March-May 2020) and therefore therapies including remdesivir, anti-interleukin-6 (IL-6) receptor monoclonal antibody, dexamethasone or anti-coagulants were not administered as usual care for COVID-19. Of the 142 patients, 58 recovered while 84 had a fatal outcome (**Figure 1A**). The primary cause of death of the fatal cases was respiratory failure alone or in the context of multiorgan failure. Patients were followed from the day of hospital admission for up to 28 days of hospitalization. Complete autopsies were performed on 34 out of the 84 patients with fatal outcome. Representative formalin-fixed paraffin embedded (FFPE) lung samples from complete autopsies of 30 fatal cases were obtained and scored independently by four pathologists (Blueprint; Delafiori et al., 2021; Farias et al., 2022; Freire Santana et al., 2020; Oliveira et al., 2022; Santana et al., 2021; WHO). FFPE samples were used for histopathological, imaging and spatially resolved single-cell proteomic tissue analysis. (**Figure 1A**). For analysis we divided patients into two groups based on outcome: those who died during hospital admission (fatal cases) and those who recovered and were discharged home from hospital within 28 days of hospitalization (recovered cases) (**Figures 1A, 1B)**.

**Figure 1:**
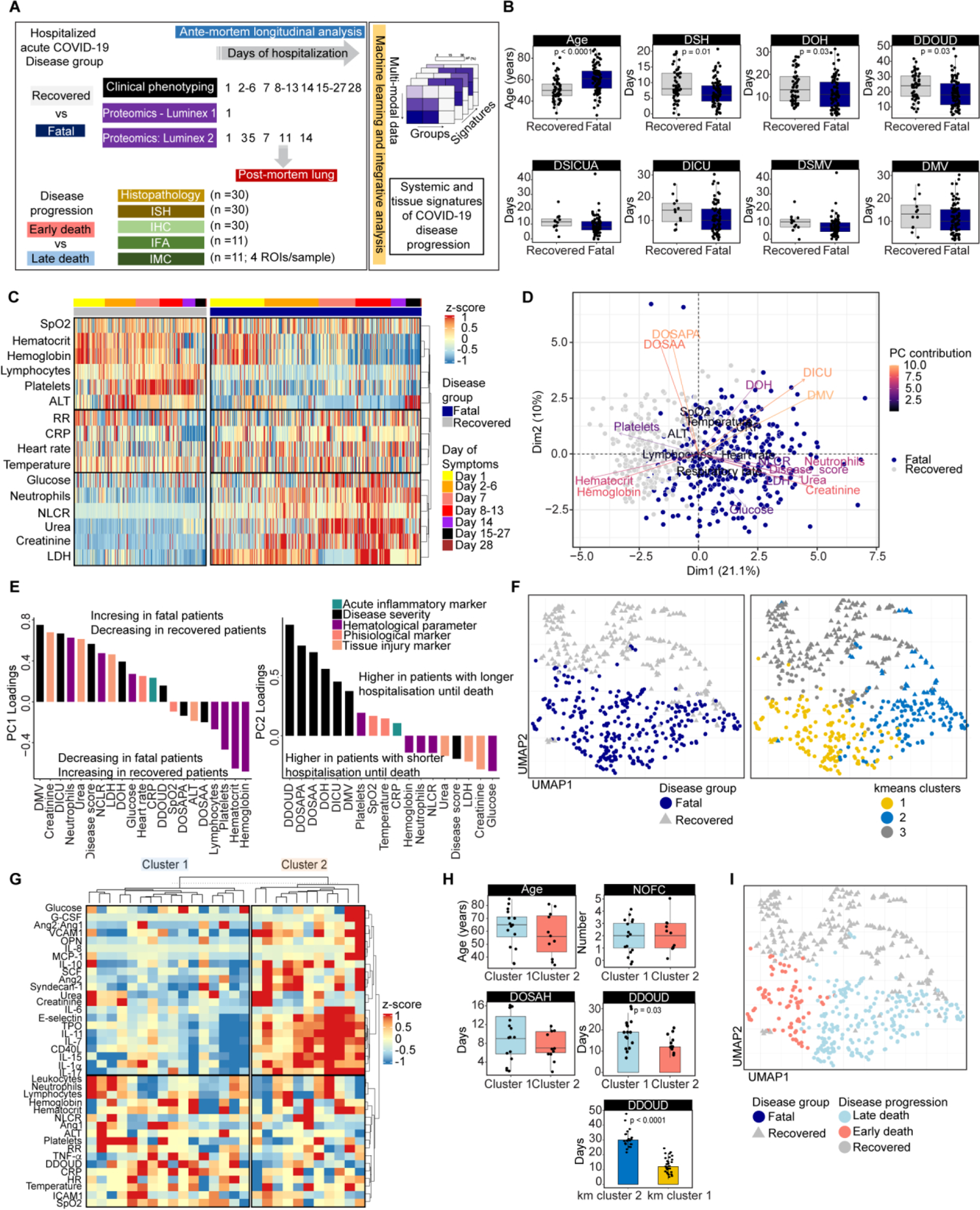
Clinical characterization of hospitalized COVID-19 recovered and fatal patients. **(A)** Cohort, study design and workflow of experimental and analytical approaches. The scheme shows the number of patients assayed per each experimental approach. Hospitalized COVID-19 cases (n=142) were grouped according to disease outcome up to 28 days of hospitalization: fatal cases (n=84) and recovered cases (n=58) were. Clinical phenotyping was recorded on the same day and during admission at 7-days intervals and at least once more between the 7-days intervals. Blood sampling, for collection of plasma, and naso/oropharyngeal secretion sampling for detection of SARS-CoV-2 by RT-PCR were collected at admission (day 1) and days 3, 5, 7, 11 and 14 after admission. The plasma samples were used in multiplex-plasma profiling, which was performed using 2 different panels. Panel 1 included plasma samples collected on day 1 of admission (27 fatal cases). Panel 2 included plasma samples collected up to 14 days of hospitalization (11 recovered cases and 11 fatal cases). Representative lung FFPE samples from full autopsies of 30 fatal cases were used for histopathological characterization and scoring of tissue lesions by H&E and quantitative investigation of the cellular landscape and virus-infected cells by in situ hybridization (ISH) and immunohistochemistry (IHC). From this set of 30 samples, 11 patient samples were further analyzed by immunofluorescence (IFA) and spatially resolved single-cell proteomic tissue analysis with Imaging Mass Cytometry by Time of Flight (Imaging CYTOF - IMC). **(B)** Demographic and temporal data of hospitalized COVID-19 recovered and fatal cases. **(C)** Heatmap representing clinical parameters longitudinally measured during hospitalization of COVID-19 recovered (grey bar on top) and fatal patients (blue bar on top). The data was scaled and the z-score value for each parameter was input in hierarchical clustering (clustering for rows only). Columns represent each patient, and they were grouped according to disease outcome (recovered=58; fatal=84) and the day of sampling from hospital admission up to 28 days, as indicated by the top bar. **(D)** Principal component analysis (PCA) shows the relationship between the clinical parameters measured during hospitalization with the individual measurements in the recovered (grey dots) and fatal (dark blue dots) COVID-19 patients. The contribution of the clinical variables in accounting for the variability in a given principal component (Dim1 and Dim2) are colour coded and their directionality in the PCA plot are represented by the arrows. Each dot represents a sample from each patient collected up to 28 days of hospitalization and colour coded following the disease outcome – dark blue dots for fatal patients and grey dots for recovered patients. **(E)** Loadings of clinical parameters on principal component 1 (PC1) and PC2 for hospitalized COVID-19 cases**. (F)** Embedding by uniform manifold approximation and projection (UMAP) and unsupervised k-means clustering of the clinical parameters of COVID-19 patients, color-coded (left to right) by disease group and k-means clustering. **(G)** Hierarchical clustering of proteins measured in the peripheral blood (PB) sample collected on hospital admission from 27 patients, who later had a fatal outcome. Measurements were normalized across all patients and the heatmap of the z-score values is represented. Euclidean distance and hierarchical clustering (ward.D) were used to cluster columns (representing individual samples) and rows (representing the measured parameters) to determine the patients’ clusters (Cluster 1, n=16; Cluster 2, n=11). **(H)** Different time of disease until death between clusters 1 and 2 defines early and late death progression of severe COVID-19 patients with fatal outcome. Age and number of comorbidities were not found as cofounders determining the differences between the 2 clusters of COVID-19 fatal patient. Importantly, the interval between onset of symptoms until death (DDOUD) is significantly lower in the cluster 2 group when compared to the cluster1, with the median of the full disease course of patients in cluster 2 being 12 days and 19 days for patients in cluster 1. Independently, k-means clustering based only on clinical parameters, confirmed that fatal patients’ separation was also associated with disease progression, with patients in the k-means cluster 1 (82% overlap with cluster 2) showing the median of the full disease course being 12 days and 30 days for patients in the k-means cluster 2 (50% overlap with cluster 1). **(I)** Embedding by uniform manifold approximation and projection (UMAP) of the clinical parameters of COVID-19 patients, color-coded by disease progression.

Longitudinal analysis revealed different clinical phenotypes between fatal versus recovered groups that could be separated by principal component analysis (PCA) (**Figures 1C, D)**. Compared to recovered patients, fatal cases had reduced respiratory function, indicated by significantly lower median SpO_2_ and increased median duration of mechanical ventilation and ICU admission during hospitalization (**Figures 1B, 1C, 1D**). These patients were also characterized by lower median neutrophil and lymphocyte counts resulting in higher median neutrophil-to-lymphocyte ratio (NLCR), a recognized feature of severe COVID-19 (Kuri-Cervantes et al., 2020; Laing et al., 2020; Lucas et al., 2020; Mann et al., 2020). Fatal cases also had increased levels of tissue injury markers, (e.g., urea, creatinine and LDH) in peripheral blood, and were more anaemic during hospitalization (reduced hematocrit and hemoglobin levels, **Figures 1E left panel, S1A**). The high variability across clinical parameters in fatal cases suggests higher heterogeneity in the trajectories of disease progression leading to a fatal outcome. Indeed, PCA (principal component analysis) and UMAP (Uniform Manifold Approximation and Projection) embedding followed by unsupervised k-means clustering confirmed separation of hospitalized COVID-19 patients into recovered and fatal outcomes and revealed stratification of fatal cases into 2 clusters (**Figures 1F, S1A left panel**).

To further investigate this putative stratification in the group of patients with fatal outcome, multiplex profiling using a bead-based assay **(Luminex panel 1 – see Table S1)** was performed with plasma samples collected at admission from 27 fatal cases, where post-mortem lung FFPE samples were also available (**Figure 1A**). Overall, clinical records and PB samples were collected on the same day as hospital admission, or no more than two days after. Unsupervised hierarchical clustering stratified patients with fatal outcome into two clusters (cluster 1, n=16; cluster 2, n=11), independent of age, sex, number of comorbidities and days of symptoms at first sampling (**Figures 1G, 1H),** and confirmed the k-means clustering based only on clinical parameters (**Figure 1F**). The main features distinguishing the two clusters were peripheral blood (PB) protein markers of myeloid response and chemoattraction, and endothelial cell (EC) activation and vascular damage (**Figures 1G, S1B, S1C)**. Interestingly, the two clusters of fatal patients were also discriminated based on disease progression, i.e., the interval between onset of symptoms until death (DDOUD – days of disease onset until death), which was significantly shorter in the patients in cluster 2 compared those in cluster 1 (median of 12 days compared to 19 days) (**Figure 1H**). The same discrimination was observed when comparing the two clusters from unsupervised k-means clustering (median of 12 days compared to 30 days) (**Figures 1F, 1H**). Thus, two independent clustering approaches deconvolve the heterogeneity in disease progression within the fatal cohort into two distinct clusters: i) fast disease progression and rapid/early increase in plasma levels of markers related to myeloid activation and recruitment, EC activation, vascular damage and coagulopathy (k-means cluster 2, hierarchical cluster 1), and ii) slower disease progression (k-means cluster 1, hierarchical cluster 2). Based on these analyses, we defined patients with DDOUD < 15 days as following an early death disease progression (n=34), while patients with DDOUD > 15 days as following a late death progression (n=50) (**Figures 1H, 1I, S1A right panel**). For downstream analysis, we have therefore stratified cases into “recovered”, “early death” and “late death” groups unless stated otherwise.

### Clinical and peripheral blood signatures in hospitalized COVID-19 patients predict disease outcome and progression

Analyses described above have stratified patients into those following an early death progression (n= 34); those following a late death progression (n= 50); and those that recovered (n= 58). To characterize the drivers of these distinct trajectories in disease progression in our cohort, we first applied Exploratory Factor Analysis (EFA) to the clinical data recorded at and up to 28 days of hospitalization. We identified three clinical signatures based on factor loadings and mapped how they varied during disease progression (Figures 2A**, S1D**). Clinical signature 1 (CS1) is positively associated with (i) markers of tissue injury, creatinine, urea, LDH and CRP and (ii) neutrophil counts and NLCR. Meanwhile, this signature is negatively associated with SpO2, ALT and the hematological parameters, platelets, and lymphocyte counts. Clinical signature 2 (CS 2) is strongly positively associated with hematocrit and hemoglobin. Finally, clinical signature 3 (CS 3) is positively associated with temperature and heart rate (Figure 2A**, left panel).** EFA clinical signatures indicate that patients following the fatal trajectory (both early and late death) show higher values of CS 1 and a rapid decrease of CS 2 compared to recovered patients during the whole period of hospitalization. However, within the fatal group, the early death progression showed higher levels of clinical signature 1 in the first days of hospital admission and a significantly faster drop of the clinical signature 2 during the first week of hospitalization, when compared to the late death cases (Figures 2A**, S1D)**. Next, we applied EFA to multiplex plasma profiling using bead-based assays **(Luminex Panel 2 – Table S1)** on a series of plasma from PB samples collected every 2-3 days during hospitalization (at and up to 14 days of hospitalization) from 11 COVID-19 fatal patients (early death=4; late death=7) and 11 recovered patients (Figures 1A**, 2B, S1E, S1F)**. The EFA output showed a rapid and sustained increase of PB signatures 1 and 2 in the early death progression during hospitalization (Figures 2B**, S1F)**. The parameters with high positive factor loadings for these signatures (i.e., characterizing the early death progression) were related to myeloid cell response, myeloid chemoattraction, EC activation, vascular damage, coagulopathy, inflammasome activation and inhibition of T-cell responses (Figures 2B**, S1F)**. In comparison, the late death progression was characterized by a slower (when compared to the early death group) and sustained increase (when compared to the recovered group) of PB signature 1. However, discriminating the late death progression from the other groups was a delayed increase of markers in the PB signatures 2 and 3 (EC activation – E-selectin, ICAM-1, IL-1β, ADAMTS13; thrombopoiesis – IL11, TPO; fibrosis – FGFb; cytotoxicity – Granzyme B; type I, II and III IFN – IFNα, IFNβ, IFNλ3, IFNγ; and Th17 responses – IL-17), as they were detected only days later during hospitalization. The recovered patient trajectory was characterized by PB signature 5, which consists of the anti-inflammatory cytokine IL-10, Th2 response (IL-4, IL-5, IL-13), lymphopoiesis (IL-7, IL-15) and myelopoiesis (G-CSF, GM-CSF) (Figures 2B**, S1F)**.

**Figure 2:**
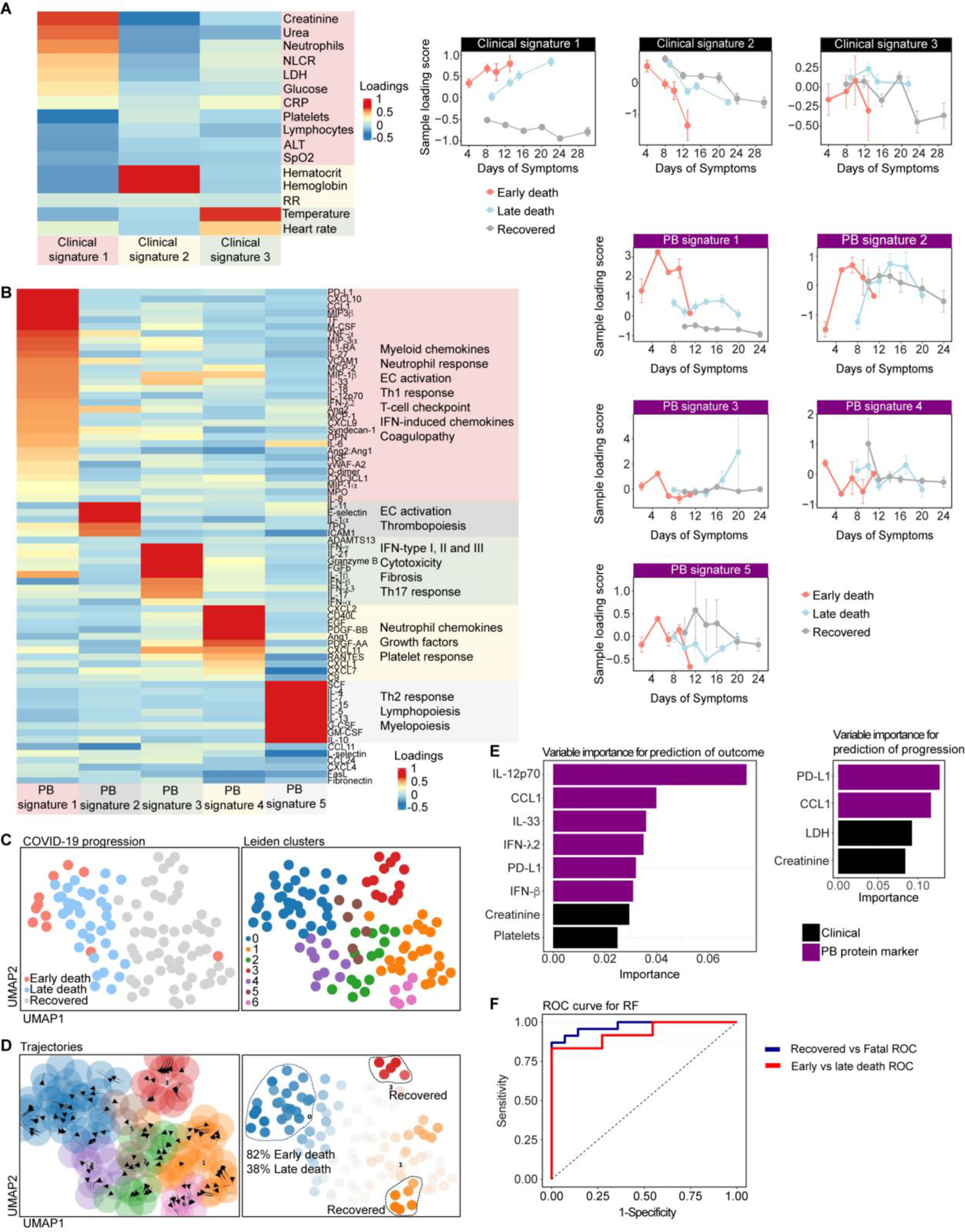
Disease trajectories of hospitalized COVID-19 patients characterized by longitudinal clinical and peripheral blood (PB) signatures. **(A)** Heatmap representing the composition (left panel) and line plots (right panels) representing the longitudinal trajectories of the 3 clinical signatures identified by Exploratory Factor Analysis (EFA). The heatmap shows the values of the factor loadings of each clinical parameter and the composition of each clinical signature after sorting the clinical parameters across each factor. The line plots show the variation of each clinical signature along the days of symptoms per each disease progression (recovered in grey, early death in light red and late death in light blue). **(B)** Heatmap representing the composition (left panel) and line plots (right panels) representing the longitudinal trajectories of the different peripheral blood (PB) signatures based on Exploratory Factor Analysis (EFA) along the days of symptoms of COVID-19 patients grouped by disease progression. The values of the factor loadings in the heatmap in the left panel provide a score for each parameter on each factor, which here we called PB (peripheral blood) signatures. Protein markers with no association with the corresponding PB signature are expected to have values close to zero, whereas markers with strong association with the PB signature are expected to have large absolute values. The sign of the factor loading indicates the direction of the effect: a positive value indicates that the marker has higher levels (enriched) in the samples with positive PB signatures values, and vice-versa. The line plots in the right panels show the variation of each PB signature along the days of symptoms per each disease progression (recovered in grey, early death in light red and late death in light blue). **(C)** Computation of transition probabilities based on similarities among patients using a KNN graph. Intermediate and terminal macrostates of our patient landscape and the drivers for each trajectory, based on the longitudinal trajectories of combined clinical parameters and PB biomarkers, were also determined. First, non-linear dimensionality reduction (UMAP) and leiden clustering were performed. Each dot represents a patient sample representing the clinical data and PB biomarker measured up to 14 days of hospitalization. **(D)** Determination of intermediate and terminal states. Cellrank’s connectivity kernel was applied to compute the transition probabilities based on similarities among patients using a k-nearest neighbours (KNN) graph. Then, terminal states were determined and plotted in the UMAP embedding. **(E)** Top clinical parameters and peripheral blood protein markers for prediction of disease outcome (left panel) and progression (right panel) in hospitalized COVID-19 patients. Set of clinical and PB biomarker variables, ranked by feature selection steps for prediction using random forests (RF)-based algorithms, generating a predictive model for COVID-19 outcome in hospitalized patients, with low redundancy and sufficient for excellent (AUROC > 0.9) or good prediction performances (AUROC > 0.7). **(F)** Area under the receiver operating characteristic curves (AUROC) representing the performance of the RF models trained for prediction of disease outcome (fatal vs recovery - blue curve) and progression (early death vs late death - red curve) in the test datasets using the top clinical and PB biomarkers for prediction of each condition.

To further characterize the trajectories in disease progression, we combined clinical and PB parameters and computed intermediate and terminal states of our patient landscape to identify the drivers for each trajectory (Figures 2C**, 2D**). After non-linear dimensionality reduction (UMAP) and clustering (Figure 2C), we computed transition probabilities based on similarities among patients using a k-nearest neighbours (KNN) graph (Bergen et al., 2020; Lange et al., 2022; Lukas Heumos, 2023). Then, we determined the intermediate and terminal states, which defined the leiden clusters 0 (82% samples from early death patients), as well as 1, 3 and 6 (100% samples from recovered patients) as progressing towards terminal states. Leiden clusters 2, 4 and 5 (62% samples from late death patients) were defined as intermediate/transitional states (Figure 2D). Analysis of the major clinical and PB protein markers driving these terminal states corroborated the EFA outputs (**Figures S2A, S2B)**. We also applied machine learning (ML) to define clinical and PB protein markers that predict disease outcome (recovered vs fatal) or progression (early vs late death). First, the combined clinical and PB dataset was partitioned in training, validation and test sets for building models using 171 ML algorithms in SIMON (Sequential Iterative Modeling ‘‘Over Night’’) (Tomic et al., 2019; Tomic et al., 2021) to identify the best performing ML predictive models and order the best clinical/PB parameters for prediction by feature selection. Based on metrics of performance, we selected the random forest (RF) model and sorted the clinical/PB parameters in descending order by their permutation-based importance (mean decrease accuracy or increase out-of-bag error), averaged over 50 RF runs (**Figures S2C-F**) (Ganggayah et al., 2019; Jiang, 2020; Ludemann et al., 2006; Speiser et al., 2019; Tuleau-Malot, 2022; Wickham, 2020; Wiener, 2002). This step determined clinical/PB parameters with low redundancy, sufficient for excellent/good (AUROC > 0.9/ > 0.7) prediction performances of disease outcome or disease progression [31, 38]. The best predictors of disease outcome were protein markers in PB signature 1 (IL12-p70, CCL1, IFN-λ2, PD-L1, IL-33, IFN-β) and clinical parameters in clinical signature 1 (creatinine and platelet counts) (Figures 2E **left panel, S2C)**. A similar set of parameters, plus LDH, best predicted disease progression leading to fatal outcome (early vs late death) (Figures 2E **right panel, S2E**). Next, we validated the practical value of these parameters in predicting disease outcome or progression when measured at or early after hospital admission. For this purpose, the RF model was trained on 70% of the dataset and then evaluated in a test dataset, using only the clinical and PB variables measured at and up to 3 days of hospitalization. The RF model had high sensitivity and specificity to predict outcome (AUROC train set 0.97; test set 1.00) and disease progression (AUROC train set 0.93; test set 0.82) (Figure 2F) (Robin et al., 2011; Sing et al., 2005).

On the bases of the data obtained, we provided examples of decision trees and cut-off values for clinical and PB parameters that could be used in clinical settings to prioritize care to patients most likely to deteriorate and, potentially, to identify precision medicine approaches to treatment (**Figures S2D, S2F**). The RF models outputs emphasize the association of the clinical signature 1 and PB signature 1 with faster clinical deterioration leading to fatal outcome in hospitalized COVID-19 patients, as both signatures are significantly higher in early death patients. Together, deconvolution of clinical and PB signatures through different systems biology approaches allowed us to deconvolve the biological processes and responses underlying these 3 distinct trajectories in disease progression in hospitalized COVID-19 patients. Patients with fatal outcome following the early death progression are characterized by dysfunctional immune response marked by combined T-cell immunosuppression and myeloid-mediated inflammation leading to EC activation and coagulopathy, present at hospitalization, indicating a coordinated myeloid and vascular inflammatory response associated with faster progression of severe disease. Late death progression is characterized by a delayed increase of a pro-inflammatory response characterized by EC activation and thrombopoiesis, fibrosis, cytotoxicity, IFN and Th17 responses. Finally, recovered patients develop a pro-lymphogenic, Th2 and anti-inflammatory-mediated responses. These data reinforce the opportunities provided by precision medicine and patient stratification for timed and targeted therapeutic approaches.

### Disease progression correlates with distinct patterns of lung lesions

Next, we aimed to verify how the trajectories of systemic signatures (clinical and PB) measured during life are associated with tissue-specific responses in the lung (Figure 1A). First, histopathological characterization was performed on H&E stained FFPE lung sections from full autopsies of 26 fatal cases (early death= 15; late death = 11). Parameters for lung evaluation and scoring were selected based on the current literature (Ackermann et al., 2020; Barton et al., 2020; Cai et al., 2020; Farias et al., 2022; Freire Santana et al., 2020; Gibson-Corley et al., 2013; Klopfleisch, 2013; Magro et al., 2020; Menter et al., 2020; Santana et al., 2021; Tian et al., 2020; Wichmann et al., 2020; Xu et al., 2020). Early and late death progression showed different frequencies and severities of different lesions in the lung (**Figure S3A**). The early death progression was associated with marked alveolar damage with type II pneumocyte (PTII) hyperplasia, venous thrombi, high immune cell infiltration (cellularity) and fibrin deposition (Figures 3A **and S3A**). In contrast the late death progression was associated with a greater degree of fibrosis and granulation and with hemorrhages, microthrombi and arterial thrombi **(Figure S3A)**, although less fibrin deposition or venous thrombi than the early death progression. Then, to assess whether lesions fit into patterns that reflect shared pathogenic processes, we grouped lesions into five morphological signatures (MS) (Figure 3A). Interestingly, early, and late death progression were associated with different MS. Early death was mainly associated with MS1 (46.7% cases *vs* 9% in late death) and MS2 (14% cases vs 9% in late death), with the frequency of MS3 slightly higher in the late death group (40% cases vs 45.4% in late death) (Figure 3B); MS1 is characterized by diffuse alveolar damage (DAD) and high infiltration of macrophages (Figure 3A**, S3A**), MS2 by diffuse alveolar damage, fibrin deposition, microthrombi and low levels of cell infiltration and MS3 is characterized by high neutrophil infiltration (Figure 3A). These findings fit with the strong procoagulation phenotype (PB signature 1) observed in the plasma of early death patients during life (Figures 2B**, S1F).** Morphological signatures 4 and 5 were only found in the lung of fatal patients following the late death progression (9% and 27.6% of late death cases, respectively) (Figure 3B). MS4 is characterized by pronounced fibrosis and collagen-rich granulation tissue almost replacing the entire lung tissue (Figures 3A**, S3A**) and MS5 by alveolar hemorrhage (Figure 3A, **S3A**).

**Figure 3:**
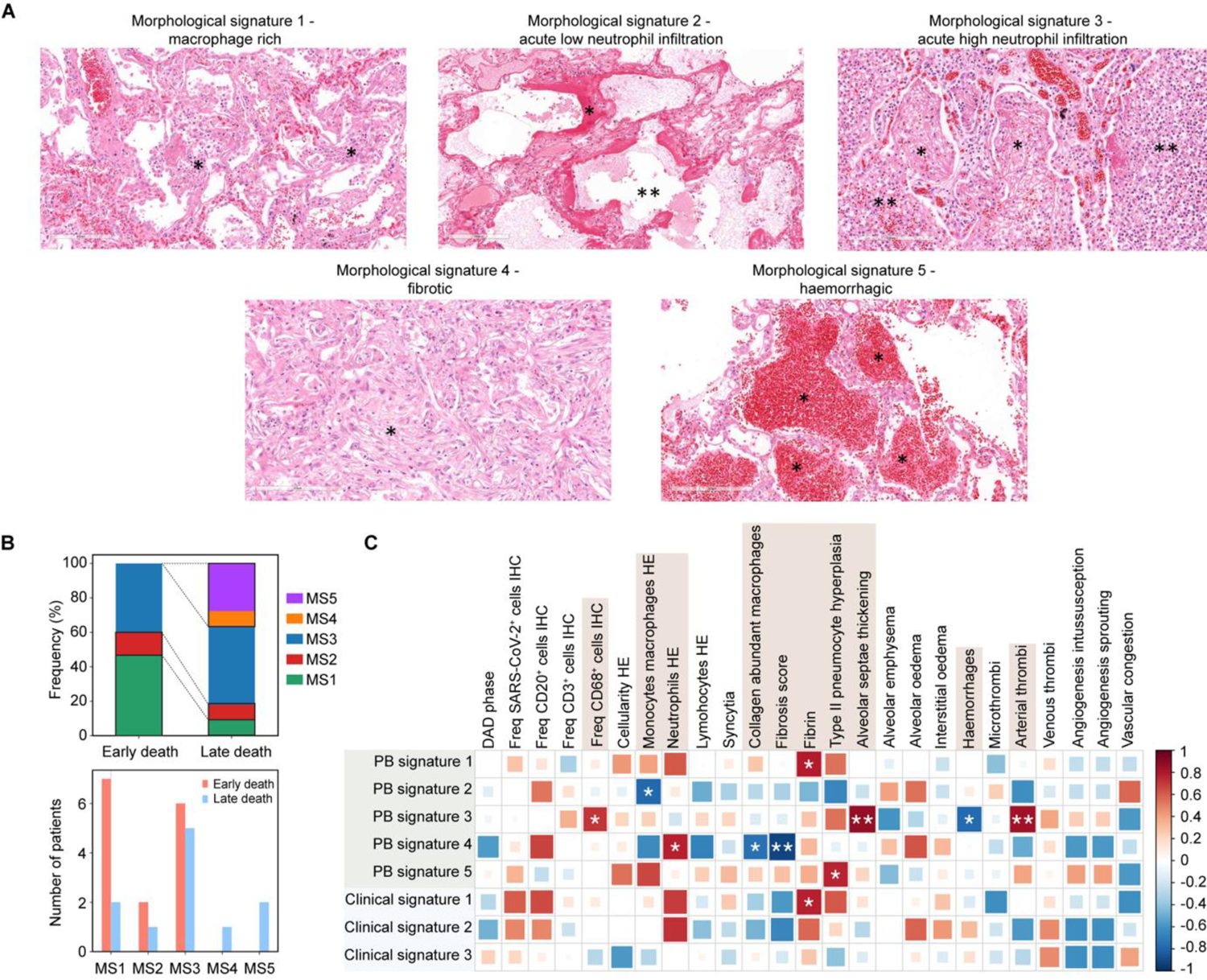
Early and late death disease trajectories exhibit distinct patterns of lesions (morphological signatures) in the lung. **(A)** Histopathological patterns (morphological signatures) of lesions found in the post-mortem lung in COVID-19 fatal cases. The macrophage rich present in the morphological signature 1 shows diffuse alveolar damage (DAD) and a dominating mononuclear cell infiltration with lymphocytes, plasma cells and alveolar macrophages (* shows mononuclear cells, including macrophages, lymphocytes and plasma cells). The morphological signature 2 shows a very acute disease phase with dominant diffuse alveolar damage, as well as fibrin and microthrombi and an overall very low cell infiltration (* shows DAD and fibrin, ** shows ruptured alveoli with alveolar oedema). The morphological signature 3 shows similar morphological findings as in the 1 but additionally displays a prominent infiltration of neutrophils in the lung (* shows fibrin in alveoli, ** shows viable and degenerated neutrophils). Morphological signature 4 represents the chronic disease stage with a pronounced fibrosis and collagen-rich granulation tissue almost replacing the entire lung tissue (* shows collagen-rich granulation tissue - fibrosis). The morphological signature 5 represents a hemorrhagic phenotype, with abundant erythrocytes in the alveoli (* shows alveoli filled with extravasated erythrocytes - hemorrhages). In summary, the histological patterns were created based on the chronicity of the disease characterized by the dominating inflammation and therefore from 1 to 4 the chronicity is increasing. Scale bar= 100μm. **(B)** Early and late death disease trajectories exhibit distinct patterns of lesions in the lung. By evaluating the histological patterns in the lung of COVID-19 fatal patients following the early and late death trajectories, we observed similar frequencies of both groups presenting inflammation dominated by neutrophil infiltration (morphological signature 3), representative of an acute disease stage. However, we observed a significant predominance of the morphological signature 1 in the lung of COVID-19 fatal patients in the early death progression (early death = 46.7% vs late death = 9%). Absent in the lung of these patients, COVID-19 fatal patients in the late death progression were also characterized by the significant presence of the morphological signatures 4 and 5 (% and 27.6%, respectively in late death patients), representing the chronic phase of disease and hemorrhagic lesions. **(C)** Correlation of the peripheral blood and clinical signatures close to death with the lung tissue lesions. *p<0.05; **p<0.01; ***p<0.001.

We further analyzed the levels of infiltration of myeloid and lymphoid cells by H&E and immunohistochemistry (IHC) **(Figures S3B, S3C).** Early death patients had higher median levels of macrophages (determined by H&E and CD68^+^ cells by IHC) **(Figure S3B)**. In contrast, no differences in the infiltration of CD3^+^ (T lymphocyte) and CD20^+^ (B lymphocyte) cells were observed between early and late death cases (**Figure S3C**). Extramedullary hematopoiesis in the lung of a patient following the early death progression was also observed **(Figure S3D)**. To verify whether clinical and PB variables measured close to death could reflect the lesions scored in the matched post-mortem lung, Spearman’s rank correlation coefficient between the systemic signatures (clinical and PB) and the scored lung lesions were determined (Figure 3C). This analysis revealed that PB signature 1 and clinical signature 1 were positively correlated with fibrin deposition (Figure 3C), a hallmark lesion of MS 1 and the most frequent pattern present in patients following the early death progression (Figure 3B). Thus, the correlative analysis provides the first orthogonal validation for how clinical and PB parameters (measured ante-mortem during hospitalization) may be able to predict the underlying specific lesions and pathogenic processes developing in the lung in patients progressing to a fatal outcome.

### Disease progression towards fatal outcome correlates with distinct viral localization and cellular landscapes in the lungs

To further characterize the pathogenic processes underlying the different trajectories leading to fatal outcome, we constructed a spatially resolved single-cell atlas of the post-mortem lung using Imaging Mass Cytometry (IMC) (Figures 1A**, 4A, S4-S6**). The integrative analysis of H&E, IHC and IFA guided the definition of regions of interest (ROIs) for subsequent IMC. We designed a 38-antibody panel for IMC **(Table S1)**, which included phenotypic markers of cell type (epithelial, endothelial, stromal and immune cells), cell function (activation, antigen presentation, cytotoxic activity and apoptosis), and an antibody specific to the SARS-CoV-2 spike (S) protein. The panel was designed to also complement the protein markers measured in the PB by Luminex **(Table S1,** Figures 1G**, 2B, S1E)**. A total of 43 highly multiplexed images at 1μm resolution were analyzed representing a total of 65 mm^2^ of tissue and 249,164 single-cells across all lung samples. After quality control checks and normalization of signal intensity, a supervised cell assignment algorithm was applied to assign single-cells to all major cell types (Geuenich et al., 2021). Next, we projected the single cells from all disease groups into a two-dimensional UMAP space and clustered them based on the expression of their marker proteins, which resulted in 22 clusters representing the cell atlas of the post-mortem lung in our cohort of fatal COVID-19 patients (Figures 4A**, S4, S5 and S6)**. In parallel, we performed a differential cell type abundance test (without relying on cell clustering) by applying Milo (Dann et al., 2022), a tool quantifying cell type enrichment based on k-nearest neighbor graphs (KNN) and implementation of negative binomial generalized linear model (GLM) (Figure 4B**, S6A**). Both approaches revealed differential abundance of cell types between the early and late disease progression groups. The early death progression is characterized by enrichment of SARS-CoV-2 antigen-positive (Ag^+^) alveolar macrophages (SARS-CoV-2^+^ AM), activated endothelial cells (ECs) and classical monocytes in the post-mortem lung (Figures 4A**, 4B, 4F, S6C, S6G)**. In contrast, the late death progression is characterized by higher abundance of SARS-CoV-2^+^ epithelial cells, CD66b^Low^ neutrophils and different apoptotic populations, such as epithelial cells, apoptotic fibroblasts, apoptotic smooth muscle cells (SMCs) and apoptotic neutrophils (Figures 4A**, 4B, 4F, S6C, S6E, S6I, S6K)**. To independently validate our findings, we re-analysed the IMC dataset of a fatal COVID-19 cohort of North American patients (Rendeiro et al., 2021) which were also stratified into early (DDOUD<30 days) and late (DDOUD>30 days) death groups **(Figures S7).** Cell clusters were determined using the same analysis pipelines as in our dataset **(Figure S7A)**. Indeed, we observed that the early death progression patients had higher frequency of SARS-CoV-2 antigens detected in the myeloid compartment, in particular in monocytes and neutrophils. In contrast, the late death progression group had a higher frequency of virus antigens in the epithelial compartment **(Figures S7B, S7C)**.

**Figure 4.**
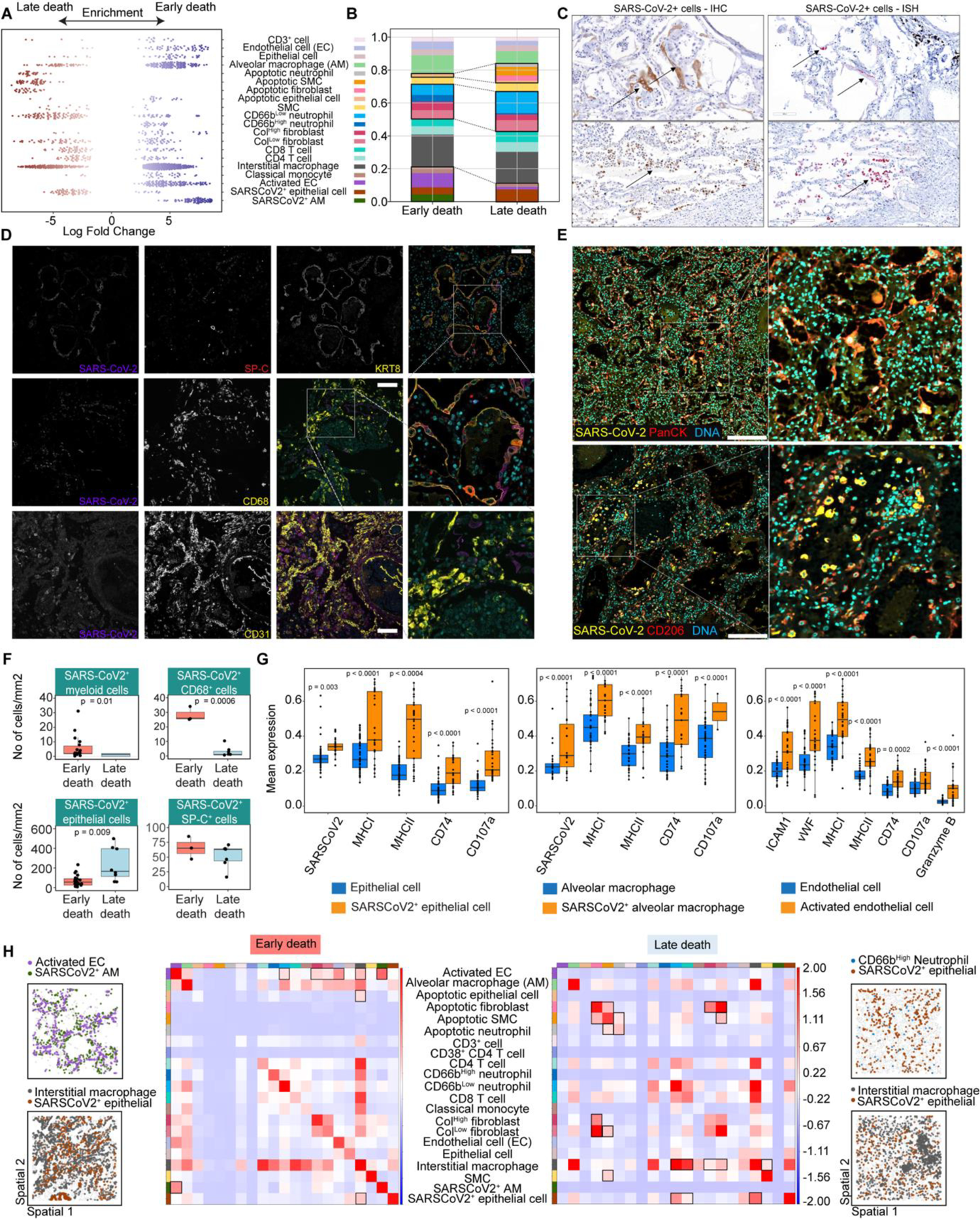
Severe COVID-19 disease progression in hospitalized patients with fatal outcome is associated with distinct spatial cellular landscapes and interactions in the lung. **(A, B)** Cell type enrichment analysis (left plot) and frequency (right plot) of the cell populations identified in the post-mortem lung COVID-19 samples using Imaging Mass Cytometry (IMC). Cell type enrichment analysis, tested with Milo, shows the logarithm of the fold-change comparing the abundance of each cell type in the post-mortem lungs from early death vs late death progression. To correct for multiple testing, the spatial false discovery rate (FDR) was calculated and only dots with spatialFDR < 0.05 are shown. In (B), the stacked bar plot shows the averaged frequency of the cell types by grouping the values from regions of interest (ROIs) according to the disease progression (early vs late death). Black boxes and dashed lines highlight groups of cell populations that are significantly different between the disease progression groups. **(C)** Representative images of post-mortem lung sections stained by immunohistochemistry (IHC - top panels) with anti-SARS-CoV-2 Nucleocapsid antibody for detection and quantification of SARS-CoV-2^+^ cells. The lower panels are representative images of post-mortem lung sections stained by *in situ* hybridization (ISH) with probes specifically targeting virus RNA for detection and quantification of SARS-CoV-2^+^ cells. Scale bar= 100μm. **(D)** Representative images of post-mortem lung sections stained by immunofluorescence (IFA) with anti-SARS-CoV-2 Nucleocapsid antibody for detection and quantification of SARS-CoV-2^+^ cells. Top panels: Grayscale representative images of lung section co-stained for virus and pneumocyte markers: SARS-CoV-2 antibody, Prosurfactant Protein C (SP-C) and Cytokeratin 8 (KRT8) and a color merged image including DAPI staining (cyan). Middle panels: Grayscale representative images of lung section co-stained for virus and macrophage markers: SARS-CoV-2 antibody, CD68 and a color merged image including DAPI staining (cyan). Bottom panels: Grayscale representative images of lung section co-stained for virus and endothelial cell markers: SARS-CoV-2 antibody, CD31 and a color merged image including DAPI staining. Scale bar= 100μm. **(E)** Representative images of post-mortem lung sections showing SARS-CoV-2^+^ cells identified by Imaging Mass Cytometry (IMC). Top panel: SARS-CoV-2 N protein (yellow), epithelial cell marker Pan-cytokeratin (PanCK in red) and DNA marker (cyan). Bottom panel: SARS-CoV-2 N protein (yellow), macrophage marker CD206 (red) and DNA marker (cyan). Scale bar= 260μm. **(F)** Quantification of SARS-CoV-2^+^ cells in the lung tissue according to disease progression. The bar plots show the number of SARS-CoV-2^+^ epithelial and myeloid cells per mm^2^ of tissue, as quantified in different ROIs (represented by each dot) by IMC, and SARS-CoV-2^+^CD68^+^ (macrophages) and SARS-CoV-2^+^SP-C^+^ (epithelial) cells by IFA, grouped by disease progression. **(G)** Functional phenotype of SARS-CoV-2^+^ alveolar macrophages, SARS-CoV-2^+^ epithelial cells and activated endothelial cells (ECs) versus non-infected or non-activated counterparts based on average expression per ROI. The ranking sum of the highly differentially expressed marker proteins for each cell population, averaged by ROI (each dot), was computed. Differentially expressed proteins were defined by using the Wilcoxon method followed by the Benjamini-Hochberg correction method for multiple comparisons. **(H)** Spatial statistics analysis reveals that distinct trajectories in disease progression result in different patterns of cellular interactions in the lung. This analysis informs on the neighbor structure of the tissue by calculating the enrichment score on spatial proximity of clusters based on a spatial graph. Here the spatial graph was built using the radius method, with radius = 20μM. The 2 left panels from the heatmap visualize the spatial organization between specific cell types based on the neighborhood graph for specific ROIs from each disease progression group, as indicated.

To further characterize the identification of SARS-CoV-2^+^ cells in the lung by IMC (Figures 4A**, 4B, 4E, S4**), we performed IHC and IFA staining using a SARS-CoV-2 nucleocapsid protein antibody and *in situ* hybridization (ISH) targeting the *spike* gene of the virus (Figures 4C**, 4D)**. These experiments confirmed that viral RNA and protein are predominantly in epithelial cells (SP-C^+^ and KRT8^+^ cells), and to a lesser extent in macrophages (CD68^+^ cells) but were absent in endothelial cells (CD31^+^CD45-) (Figures 4C, 4D, 4F). We also observed significantly higher numbers of SARS-CoV-2^+^ myeloid cells in the lungs of early death patients, while virus antigens were enriched in the epithelial compartment in the lungs of late death cases (Figure 4F). In our cohort, SARS-CoV-2^+^ alveolar macrophages (SARS-CoV-2^+^ AM) expressed significantly higher levels of the proteins associated with antigen presentation (e.g., MHCI, MHCII, CD74 and the marker of cytotoxic granules, CD107a) compared to virus antigen-negative alveolar macrophages (Figure 4G). Similar differences were observed between SARS-CoV-2^+^ epithelial cells compared to virus antigen-negative epithelia (Figure 4G). Finally, there were higher levels of EC activation in the lungs of early death patients, including higher expression of canonical activation markers (ICAM-1, vWF) as well as of proteins associated with antigen presentation (MHCI, MHCII, CD74) and the cytotoxic marker, Granzyme B (Figure 4G).

### Distinct patterns of single-cell interaction networks in the lungs characterize COVID-19 disease progression

We next generated a quantitative map of cellular interactions in the lungs of patients who died as a result of COVID-19 according to disease progression. We applied different statistical methods to determine homotypic (within cell-types) and heterotypic (between cell-types) proximal spatial enrichment as a proxy for cellular interactions. By using different spatial statistics approaches, we observed significant enrichment of homotypic interactions between SARS-CoV-2^+^ macrophages and activated ECs, and heterotypic interactions between SARS-CoV-2^+^ alveolar macrophages and activated ECs in the lungs of early death patients (Figures 4H**, S8A, S8B).** There was a strong correlation between the frequencies of activated ECs and SARS-CoV-2^+^ alveolar macrophages (**Figure S9A**), which indicates that the proximity of these cell types to each other could either be spatially coordinated or stochastic. Correction of rates of cellular interactions for differences in cellular abundances still shows significant spatial proximity between these cell types confirming their interactions are coordinated and not simply by chance (**Figure S8A**). The neighborhood graph of the lung tissue of late death patients shows enriched interactions of SARS-CoV-2^+^ epithelial cells with neutrophils and interstitial macrophages. In addition, we also observed enriched interactions between fibroblasts, SMCs and neutrophils with their apoptotic counterparts (Figures 4H**, S8C, S8D**). The lack of correlation between the frequency of these cell types indicate that their interactions are spatially coordinated rather than stochastic (**Figure S8C, S9B**). Apoptosis and fibrosis pathways, such as increased frequency of lung fibroblasts, (**Figures S7B, S7C**) were also observed to be upregulated in COVID-19 late death cases in a North American cohort (Rendeiro et al., 2021).

Interestingly, analysis of changes in protein expression due to specific cell interactions showed that in the early death lungs, upregulation of endothelial cell activation markers is induced when ECs are in close proximity of SARS-CoV-2^+^ alveolar macrophages, while only in the late death lungs EC activation is induced by EC proximity to CD66b^High^ neutrophils (**Figures S8E, S8F**). Thus, the single-cell spatially resolved data increases the granularity of tissue features characterizing lung pathology and the biological processes underlying and discriminating the distinct disease progression groups.

### Integrated systemic and tissue signatures reveals hallmarks of severe COVID-19 progression leading to fatal outcome

Thus far, our dataset contains parallel characterization of the systemic *in life* signatures (longitudinal clinical records and PB protein markers data) and then post-mortem tissue features (patterns of tissue lesions and spatially-resolved single-cell data). This raises the opportunity to identify the systemic signatures (clinical and PB protein markers), that when measured during hospitalization (accessible in life), could predict the underlying lung pathological responses. For this, we sought to integrate the clinical data, PB profiling and tissue features (based on IMC) from fatal cases, by using multi-omics integrative and tensor approaches (Argelaguet et al., 2020; Argelaguet et al., 2018; Chang et al., 2021; Fanaee and Thoresen, 2019; Hore et al., 2016; Josse and Husson, 2016; Pagès, 2014; Velten et al., 2022), which disentangles the driving sources of variation across the disease trajectories leading to fatal COVID-19 outcome. First, we applied Multiple Factor Analysis (MFA) (Figures 5A**, 5B, S10A, S10B)**. The MFA output showed a distinct separation of the early and late death patients in the dimensionality reduction plot (Figures 5A**, S10A)**. The tissue signatures enriched in the early death progression comprise those highly positively correlated to Dim1 and contributing to Dim 1, such as (i) abundance of and cellular interactions involving SARS-CoV-2^+^ alveolar macrophages, (ii) mean expression of SARS-CoV-2 antigens in these cells; (iii) abundance of and cellular interactions involving activated ECs and mean expression of ICAM1, vWF, MHCI in these cells (Figures 5B**, S10A, S10B)**. PB biomarkers capturing most of the variances defining the early death progression (highly positively correlated and contributing to Dim 1) were mostly those in the PB signature 1 (Figure 2B), such as markers of activated ECs, myeloid cells and coagulopathy (Figures 5B**, S10A, S10B**), which cross-validate tissue and systemic data. Other parameters showed to be increased in early death were related to clinical signature 1 (Figure 2A), such as tissue injury markers, creatinine, urea, and ALT (Figures 5B**, S10A, S10B)**. Meanwhile, the MFA data shows that the late death progression is mostly driven by (i) tissue features associated with abundance of and cellular interactions involving SARS-CoV-2^+^ epithelial cells, counts of apoptotic epithelial cells, apoptotic neutrophils, apoptotic SMCs (smooth muscle cells), apoptotic fibroblasts, and fibrosis (counts and frequencies of SMCs and fibroblasts) in the lung (Figures 5B**, S10A)**; (ii) reduced levels of clinical parameters and PB markers present in the clinical signature 1 and PB biomarker signature 1 (opposite direction in the plot) (Figures 5B**, S10A)**. By also applying MFA, we could independently validate similar relationships between clinical parameters and post-mortem lung tissue features driving early and late death progression in a fatal COVID-19 north-American cohort (**Figure S7E**) (Rendeiro et al., 2021).

**Figure 5:**
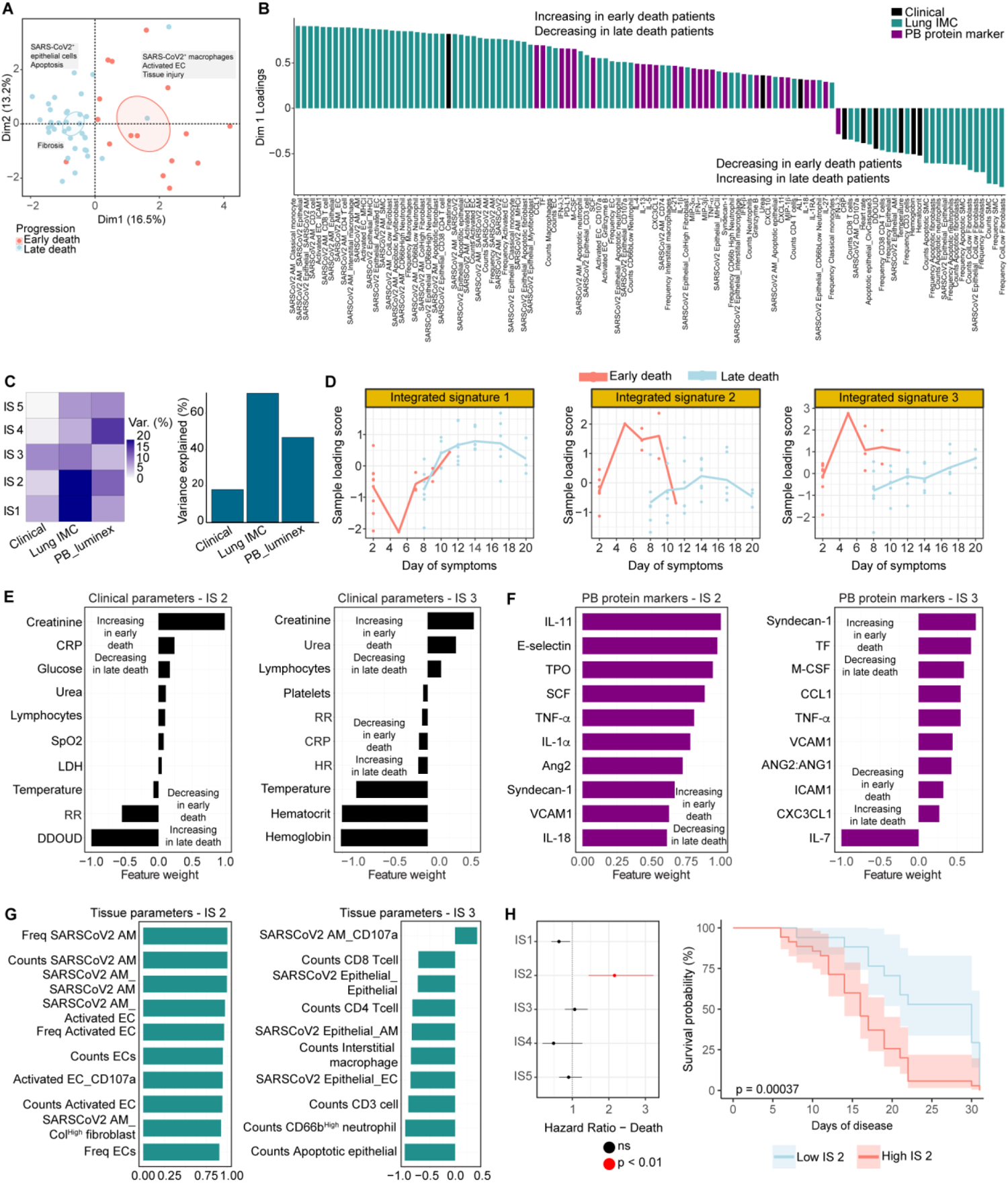
Integration of systemic and tissue signatures reveal the molecular signatures underlying the distinct disease trajectories leading to fatal outcomes in COVID-19. **(A)** Multiple Factor Analysis (MFA) based on longitudinal clinical, peripheral blood and lung tissue signatures show the underlying factors driving the early and late fatal disease trajectories. To characterize the driving sources of variation across the disease trajectories leading to fatal COVID-19 outcome we used multi-omics tensor approaches. First, MFA was applied. The following tissue features were extracted from IMC and used as inputs: (i) frequency (freq) and absolute counts of different cell types; (ii) scores of neighborhood enrichment of cell populations with SARS-CoV-2^+^ alveolar macrophages (SARSCoV2 AM) and SARS-CoV-2^+^ epithelial cells (SARSCoV2 Epithelial); (iii) expression levels of markers related to antigen presentation (MHCI, MHCII, CD74), cytotoxicity (CD107a, Granzyme B), apoptosis (ClvCaspase3) and endothelial cell activation (ICAM-1, vWF) in SARS-CoV-2^+^ alveolar macrophages, SARS-COV-2^+^ epithelial cells and activated ECs In the plot, each dot represents a patient sample (collected up to 14 days of hospitalization or at time of death) and they are color-coded accordingly to the disease progression (light red, early death; light blue, late death). The ellipses (color coded accordingly to disease progression) represent point concentration ellipses with their size defined by the 95% confidence interval around the group mean points. A summary label of the underlying factors driving the early and late fatal progression are indicated in the boxes. **(B)** Loadings of tissue and systemic parameters on dimension 1 of the MFA. The first dimension (Dim1) of the MFA plot shows the top tissue and systemic parameters capturing most of the variance defining and contributing to the separation of early and late death progression. The loading scores of these parameters were plotted. Post-mortem lung features (green bars), clinical parameters (black bars) and PB biomarkers (purple bars) with positive loading scores for PC1 are expected to show higher values (enriched) in patients following the early death progression. Meanwhile, negative values in PC1 loading scores indicate those parameters expected to be enriched in patients following the late death progression. Freq = frequency; the underscore sign “_” between cell type labels indicates interacting cell pairs; the underscore sign “_” between a cell type label and a protein marker indicates protein expression in the corresponding cell type. **(C)** Variance decomposition by factor (integrated signature – IS) and total variance explained by view. The plot in the left panel (variance decomposition by factor) summarizes the sources of variation in a complex heterogeneous dataset by showing the percentage of variance explained by each IS across each data modality (clinical, PB biomarker and IMC). The plot in the right panel represents the total variance explained per view, and it shows the total variance captured by all integrated signatures from each source approach. **(D)** Sample loading scores for each integrated signature (IS) along the days of symptoms. The loading scores of each patient sample (dots) for each IS were plotted against the days of symptoms in line plots. The lines represent mean values, and they were color coded according to the disease progression (early death – light red; late death – light blue). **(E)** Inspection of the top clinical parameters associated with the integrated signatures 2 (left panel – IS 2) and 3 (right panel – IS 3), those driving most of the variance underlying the early and late death progression. The plots show the weights of each clinical parameter in the integrated signatures 2 (right panel) and 3 (left panel). Clinical parameters with no association with the corresponding factor are expected to have weights close to zero, whereas parameters with strong association with the factor are expected to have large absolute values. The sign of the weights indicates the direction of the effect: a positive weight indicates that the parameter has higher levels in the samples with positive factor values, and vice-versa. Finally, groups with different signs manifest opposite phenotypes along the inferred axis of variation, with higher absolute value indicating a stronger effect. **(F)** Inspection of the top PB biomarkers associated with the integrated signatures 2 (left panel) and 3 (right panel), those driving most of the variance underlying the early and late death progression. **(G)** Inspection of the top tissue features associated with the integrated signatures 2 (left panel) and 3 (right panel), those driving most of the variance underlying the early and late death progression. Freq = frequency; the underscore sign “_” between cell type labels indicates interacting cell pairs; the underscore sign “_” between a cell type label and a protein marker indicates protein expression in the corresponding cell type. **(H)** Systemic and tissue signatures underlying integrated signature 2 significantly predict patients’ survival rate during disease progression. The left panel represents the ratio of the absolute coefficients for the hazard of death and p-values for each integrated signature computed by Cox proportional hazards model. Factors with significant influence on fatal outcome show high absolute coefficient in the Cox model. If the coefficient is positive, patients with large values for that specific factor have an increased hazard (of death) compared to patients with lower values. The right panel represents Kaplan-Meier plots, showing the % of survival probability over time, after patients were split into two groups based on the values of integrated signature 2 using the maximally selected rank statistics.

Next, we applied MOFA (Multi-Omics Factor Analysis) and MEFISTO (Method for the Functional Integration of Spatial and Temporal Omics data) (Argelaguet et al., 2020; Argelaguet et al., 2018; Velten et al., 2022) to generate a multi-omics factor analysis model (Figures 5C-F**, S10C-E)**. The MOFA factors 1-3, henceforth “integrated signatures” (IS) 1-3, capture variation between 20% and 70% of the total variance across all data modalities (Figures 5C**, S10C).** Most of the IMC variability driving differences between early and late death progression are well represented in IS 1 and 2, the PB biomarkers variability is mostly captured by IS 2, and finally, clinical data variability is mainly captured by IS 3 (Figure 5C).

Integrated signature 1 (IS1) is enriched in patients following the late death and reduced in those following the early death progression. The left plot and the heatmap (right panel) show the following top tissue features, extracted from the IMC, with positive weights in the IS1: (i) frequency and counts of apoptotic smooth muscle cells (SMC), frequency and counts of fibroblasts and apoptotic fibroblasts; frequency and counts of apoptotic neutrophils. Meanwhile, the following tissue features show negative weights in IS1 (thus, reduced in the late death, increased in the early death progression): cellular interactions with SARS-CoV-2^+^ alveolar macrophages and SARS-CoV-2^+^ epithelial cells; expression levels of ICAM1 in activated ECs. Freq = frequency; the underscore sign “_” between cell type labels indicates interacting cell pairs; the underscore sign “_” between a cell type label and a protein marker indicates protein expression in the corresponding cell type.

The inferred integrated signatures values for each disease progression were plotted against the days of symptoms (Figure 5D) and days of sampling **(Figure S10D)** during hospitalization. Values of IS 2 and 3 are significantly higher in COVID-19 fatal patients following the early death progression, while IS 1 is enriched in the late death progression. Once fitted, the model allows the identification of the biological features underlying each MOFA integrated signature, by inspecting the top parameters with positive and negative weights (Figures 5E-G**, S10E)**. Because IS 2 and 3 show higher values in the early death progression (Figures 5D**, S10D)**, patients following this trajectory show enrichment/higher values of the clinical parameters, PB protein markers and tissue features with positive weights, as well as lower levels of those parameters with negative weights for IS 2 and 3 (Figures 5E-G**, S10E)**. Note that independent MOFA/MEFISTO analysis corroborates the MFA outputs (Figures 5A**, 5B, S10A)** and the EFA analysis, as most of the clinical parameters and PB markers with positive weights in IS 2 and 3 overlap with clinical and PB signatures 1, elevated in early death patients (Figures 2B**, S1D, S1F, 5A, 5B, 6F right panel, S10A, S10D)**. The integrated signature 1 (IS1), which variance is mostly derived from IMC data, confirms higher abundance of fibroblasts and apoptotic populations (fibroblasts SMCs and neutrophils) in the lung of late death progression patients **(Figure S10E),** as well as cellular interactions involving SARS-CoV-2^+^ alveolar macrophages and EC activation as predominant processes in the lung of the early death progression (**Figure S10E**). We further validated the MOFA/MEFISTO outputs by applying other systems biology integration approaches, such as machine learning (ML) with random forests-based feature selection (Jiang, 2020; Speiser et al., 2019; Tuleau-Malot, 2022; Wiener, 2002) (**Figure S11**) and sparse decomposition of arrays (SDA) (Chang et al., 2021; Fanaee and Thoresen, 2019; Hore et al., 2016) (**Figure S12**). We also applied MOFA to integrate the systemic and tissue signatures of the Amazonian and north-American fatal COVID-19 cohorts (Rendeiro et al., 2021). Integrated signatures from both cohorts corroborated our observations by showing SARS-CoV-2 positivity in the myeloid compartment and EC activation as the predominant biological processes in the lungs in the early death progression, while apoptosis and fibrosis as tissue hallmarks of the lungs in the late death progression (represented by IS 1; components 2 and 4) (**Figures S11A-C, S12**).

Finally, the factors inferred by MOFA could be used to predict clinical outcomes such as time to treatment or survival times (Argelaguet et al., 2020). Integrated signature 2 was the only factor with significantly high and positive coefficient in the Cox model (Figure 5H**, left panel)**. Next, using the maximally selected rank statistics, samples were split into two groups based on the integrated signature 2 values (low IS 2 and high IS 2), and we calculated the estimate survival probabilities along the days of disease (Figure 5G**, right panel**). The survival analysis confirms our observations that hospitalized COVID-19 patients showing elevated levels of the clinical and PB parameters in the integrated signature 2 have a faster disease progression, thus, shorter survival times (early death) (Figure 5H**, right panel)**. To validate and to verify the practical value of the top 10 clinical and PB parameters in the IS 2 (here referred as “big panel”) (Figures 5E**, 5F**), in predicting disease progression, when measured at or early after hospital admission (up to 3 days), we trained a RF model and then evaluated its prediction performance in a test dataset (**Figures S11E, S11F)**. Aiming at translating discoveries into a clinically actionable assay that is broadly accessible, we sought to determine the minimum threshold of markers that could be used to predict progression without compromising accuracy, with the goal of reducing our panel to approximately three clinical parameters and three PB parameters, which is more likely to be amenable to clinical routine. Thus, based on the ranking of factor weights in the IS 2, we combined the top two, three, four or five markers, that can be measured by laboratory tests in a blood sample, and tested whether combinations could predict progression in our validation dataset with high accuracy. Using this approach, we achieved similar high accuracy (big panel AUROC train set 0.78; test set 0.83; small panel AUROC train set 0.84; test set 0.84) when we used three clinical parameters (creatinine, CRP and urea) and three PB protein markers (IL11, E-selectin and TPO) in the integrated signature 2 (here referred as “small panel”), in comparison to big panel (**Figures S11E, S11F**). Examples of decision trees and cut-off values for the top three clinical and PB biomarker parameters measured up to three days of hospitalization exemplify how these parameters, such as creatinine, urea, E-selectin and TPO, could assist in prediction of disease progression and patient stratification in the clinical setting with the aim to evaluate the best therapeutic strategies (**Figure S11E**).

## Discussion

Severe and life-threatening complications from SARS-CoV-2 results mainly from parenchymal lung pathology leading to severe hypoxia. Therapeutic interventions focus on improving the outcome of critical processes occurring in the lungs, which cannot be fully understood from analyses of blood samples. Thus, it is critical to understand how blood biomarkers (taken at hospital admission) could both (i) predict the development of lung pathology, and (ii) stratify patients into appropriate therapeutic interventions. Studies on the host responses in the lungs of COVID-19 patients have lagged behind those in peripheral blood. Most data so far is derived from single-cell and single nucleus RNA sequencing (Delorey et al., 2021; Melms et al., 2021; Wendisch et al., 2021), which added valuable insights in the understanding of COVID-19 host responses in the lung at the transcriptomic level, but are limited by lacking spatial resolution.

In this study, we applied multimodal approaches to longitudinal data from clinical records, PB profiling and high-dimensional spatial characterization of lung lesions, to link systemic responses with pathological processes developing in the lung. This approach allowed us to disentangle and characterize phenotypes associated with distinct trajectories of disease progression in hospitalized COVID-19 patients. We identified early predictors of clinical outcomes and disease progression, which could assist in patient stratification at hospital admission to determine the most appropriate therapeutic regimes. To our knowledge, this is the first study linking detailed investigation of serial clinical data and peripheral blood samples taken during life, to detailed histopathological and spatially-resolved single-cell investigation of lung samples in death in any acute respiratory infection. This approach therefore has implications for understanding COVID-19 host responses and disease progression but also as a proof of concept for other lung conditions.

Analyses of the longitudinal trajectories of clinical data suggested heterogeneity within the cohort of patients with fatal outcome. One important driver of this heterogeneity was differences in the time between disease onset until time to death. Patients with faster disease progression (DDOUD<15 days; early death), showed rapid/early increase (detected at hospital admission) in plasma levels of markers related to myeloid activation and recruitment, EC activation, vascular damage, and coagulopathy than patients with slower disease progression (DDOUD>15 days; late death). Further analysis of systemic and clinical features indicated that patients with faster disease progression to fatal outcome presented with increased tissue injury, neutrophilia and lymphopenia (i.e., higher neutrophil-to-lymphocyte ratio, NLCR) and anemia during hospitalization (clinical signature 1). Notably, higher median NLCR is a recognized feature of worse prognosis in severe COVID-19 (Kuri-Cervantes et al., 2020; Laing et al., 2020; Lucas et al., 2020; Mann et al., 2020). The underlying host responses detected in the PB characterizing the early death progression (during hospitalization) are related to a rapid and sustained increase in markers of myeloid response, chemoattraction, EC activation, vascular damage, coagulopathy, inflammasome activation and inhibition of T-cell responses/exhaustion (PB signature 1). Biological processes characterizing the late death progression, detected systemically only 2 weeks after hospitalization in these patients, included an increase in markers of EC activation and thrombopoiesis, fibrosis, cytotoxicity, IFNs and Th17 responses. The progression leading to recovery was characterized by a predominant Th2 response, lymphopoiesis and myelopoiesis. Random forests (RF)-based predictive models confirmed that, when only systemic parameters (clinical records and PB profiling) are available, the best predictors of disease outcome (recovery vs death) were protein markers in the PB signature 1, such as IL12-p70, CCL1, IFN-λ2, PD-L1, IL-33, IFN-β, and clinical parameters in the clinical signature 1, such as creatinine and platelet counts. A similar set of parameters, plus the tissue injury marker LDH, were the best parameters to predict the 2 trajectories in disease progression to fatal outcome (early vs late death). Examples of decision trees and cut-off values for clinical and PB parameters resulted from the RF models are a proof-of-concept in how they could be useful in clinical settings to assist in patient stratification for precision medicine approaches to treatment and for prioritizing care to patients most likely to deteriorate. Interestingly, trajectory inference analysis identified the early death progression and recovery, but not the late death, as terminal states. The observation that patients with early death progression heading towards a terminal state when presenting at hospital (i.e., even before admission to intensive care) indicates they already have severe immune dysregulation. This dysregulation is characterized by excessive myeloid response, EC activation and coagulopathy along with lymphopenia and T-cell inhibition/exhaustion. This suggests that treatment likely needs to be aggressive and given quickly if it is to modulate outcome using agents such as inhibitors of EC activation/coagulation and immunomodulatory therapies targeting specific cytokine signaling pathways, such as IL-6, IFNs or chemokines involved in myeloid recruitment (Gracia-Hernandez et al., 2020). Interestingly, trajectory inference analysis defining the late death progression as an intermediate state indicates that patients in this trajectory could progress either way, towards recovery or death. This analysis suggests that differences between patients with severe disease/late death outcome and those with severe disease/recovery outcome might become more pronounced with time, and that these dynamic changes reflect underlying disease processes developing in the lung during hospitalization, which cannot be characterized by clinical and PB data only. Access to matched post-mortem lung samples from a set of patients with fatal outcome is a unique opportunity to further investigate how these trajectories of systemic signatures (clinical and PB protein markers) measured ante-mortem during hospitalization, are associated with, and are predictive for biological processes, responses and lesions developing in the lung.

Based on histopathological features, we defined five distinct patterns of lung lesions in the human lung after SARS-CoV-2 infection (Ackermann et al., 2020; Barton et al., 2020; Cai et al., 2020; Farias et al., 2022; Freire Santana et al., 2020; Gibson-Corley et al., 2013; Klopfleisch, 2013; Magro et al., 2020; Menter et al., 2020; Santana et al., 2021; Tian et al., 2020; Wichmann et al., 2020; Xu et al., 2020). The lung of early death patients was characterized by marked alveolar damage with type II pneumocyte (PTII) hyperplasia, venous thrombi, high macrophage infiltration and fibrin deposition. These features reflected the biological processes that we detected by the PB profiling. In contrast, lungs in late death progression were characterized by higher levels of fibrosis, hemorrhages, microthrombi, arterial thrombi and granulation tissue. By developing a spatially resolved single-cell atlas of the post-mortem lung, we further disentangled the pathogenic processes underlying the different trajectories leading to fatal outcome. The IMC data demonstrated that post-mortem lungs in the early death progression are characterized by higher numbers of SARS-CoV-2^+^ alveolar macrophages (SARS-CoV-2^+^ AM), corroborating the macrophage-rich pattern defined by H&E analysis. They are also characterized by higher numbers of activated endothelial cells (ECs) and their interactions with SARS-CoV-2^+^ AM. Upregulation of EC activation markers is observed in ECs in close proximity to SARS-CoV-2^+^ AM. These cellular interactions together with integration of PB and tissue data indicate that lung endothelial cell activation and injury might be triggered by macrophage infiltration and activity (such as phagocytosis of SARS-CoV-2 antigens and antigen presentation). These processes might trigger pro-coagulant pathways and thrombin-mediated inflammation (Burzynski et al., 2019; McGonagle et al., 2020), which results in further activation of the clotting cascade, high levels of fibrin deposition, as observed in the lungs (Dorward et al., 2021). These coagulopathic processes ultimately result in elevated coagulation markers in PB including elevated D-dimer levels through the degradation of fibrin rich thrombi (Teuwen et al., 2020). The post-mortem lungs in the late death progression are characterized by high amounts of SARS-CoV-2^+^ epithelial cells, neutrophil infiltration and a high frequency of different apoptotic cell populations (epithelial cells, fibroblasts, smooth muscle cells (SMCs) and neutrophils). The increase in apoptotic cells in the late death progression is consistent with chronic lesions observed in the H&E analysis in the lungs of late death patients, such as fibrosis, granular tissue, and hemorrhagic lesions. Different from early death, EC activation in the late death lungs seems to be induced by close proximity of ECs to CD66b^High^ neutrophils. This indicates that thrombotic processes observed in the lungs in the late death progression might be the result of the high neutrophilic infiltration and neutrophil-mediated damage from degranulation or neutrophil extracellular traps, which could also contribute to endothelial injury and coagulation, as observed in other settings (de Bont et al., 2019; Gupta et al., 2010; Huang et al., 2020; Perdomo et al., 2019) and demonstrated in COVID-19 patients (Middleton et al., 2020). Combined, our results show different potential pro-coagulant triggers that could contribute to vascular and thrombotic lesions in the lung at different stages of disease. Hence, the pathological processes developing in the lungs of early death patients are mostly driven by macrophage-mediated inflammatory damage and activation of ECs along with interactions involving these cell types, which substantiate and add granularity to previous reports identifying the importance of mononuclear phagocyte dysfunction in severe COVID-19 (Bernardes et al., 2020; Bost et al., 2021; and Consortium, 2022; Mann et al., 2020; Schulte-Schrepping et al., 2020). Thus, patients following early death progression might benefit from therapeutic approaches targeting myeloid activation and chemoattraction (Gracia-Hernandez et al., 2020), EC activation and coagulopathy.

In contrast, our data indicate that much of the underlying pathology of the chronic lesions in the post-mortem lung in the late death progression might be due to an inadequate early response to clear the virus, then persistence of high amounts SARS-CoV-2 antigens in the epithelial compartment (even after 20 days of disease). This might lead to high levels of apoptosis, inefficient/dysregulated alveolar epithelial and stromal repair responses, resulting in increased fibrosis (Melms et al., 2021; Olajuyin et al., 2019; Praveen Weeratunga, 2022). Importantly, our analyses cannot determine whether virus positive cells reflect an active (chronic) infection or simply reflect the presence of high levels of residual viral antigen in the absence of virus replication, although the latter seems more likely. Notably, early in hospitalization, late death patients do not show a distinguishing enrichment of any of the PB signatures detected by our exploratory analysis, they are identified as an “intermediate” state in the trajectory analysis, until 2 weeks of hospitalization when specific signatures start to predominate. Thus, integrated lung and systemic data indicates that the clinical response of patients following late death progression do not have a hyperactivated immune state at admission and thus may not benefit from immunosuppressive therapies like dexamethasone. In contrast, in this late death group, early administration of therapies to stimulate virus-targeted immune responses and rapidly clear the high levels of virus, may be beneficial and shift the disease trajectory towards recovery.

Re-analysis of the lung IMC dataset and associated clinical data of a fatal COVID-19 cohort of north American patients (Rendeiro et al., 2021) that were also stratified into early and late death groups independently validated several of the conclusions of this study. However, the pathological processes and responses progressed slower in the North American population: early vs late death trajectories are defined by patients with disease course up to 30 days (early) vs longer than 30 days (late) in the US cohort. Hence, the course of severe disease seems to be much faster, with narrower intervals between symptoms onset to fatal outcome in the Amazonian cohort. To what extent this is due to differences in treatment in this lower resource setting versus population-specific differences is unclear.

By applying RF-based predictive models based on the integrated systemic and lung tissue data, we were able to identify a set of parameters that can be measured by laboratory tests in a blood sample at the time of hospital admission to predict disease progression. The set identified here is comprised of three clinical parameters related to tissue injury (creatinine, CRP and urea) and three PB protein markers related to EC activation and coagulopathy (IL11, E-selectin and TPO). Decision trees and cut-off values for these parameters measured up to 3 days of hospitalization exemplify how they could assist in prediction of disease progression and patient stratification in a clinical setting with the aim to evaluate the most suitable therapeutic strategies. Validating our findings, a study in a cohort of severe COVID-19 patients in China independently identified CRP as one of the top predictors distinguishing severe COVID-19 patients with or without fatal outcome (Shu et al., 2020), while different clinical trials utilized CRP as a biomarker of severity (Bolouri et al., 2021; Bronte et al., 2020; Group, 2021; Herold et al., 2020). Peripheral blood protein markers related to endothelial injury and coagulation, such as Ang2, vWF-A2 and D-dimer, and platelet activation were also found elevated in hospitalized COVID-19 patients, in particular in those with fatal outcome, in cohorts in China and in the UK (Ren et al., 2021; Thwaites et al., 2021). The importance of examining NLCR in patients for prognosis in COVID-19 is well known (Fu et al., 2020; Terpos et al., 2020) but the assessment of neutrophil to T-cell ratio is more stringent (Liu et al., 2020).

An important aspect to highlight is that the added value of integrating ante-mortem longitudinal systemic signatures and post-mortem lung data is the improvement in the lists of parameters identified as predictors of disease progression. If we had only access to PB and clinical data, targets such as T-cell inhibition/exhaustion or type III IFN responses would be the main predictors and preferential targets for treatment. However, when the integration with the lung tissue data is added in the predictive models, we can observe that EC activation and coagulopathy markers become the main predictors of disease progression. Incorporating these tissue-specific markers into the identification of treatment targets may be important for optimal treatments.

An important limitation of our study is that the data generated is from the first wave of the COVID-19 outbreak in a cohort of immunologically naive patients. We do not know to what extent our findings would be applicable in the context of newer variants, and more importantly patients with a variable degree of background immunity provided by vaccination or previous infections.

Taken together our study provides the first integration of longitudinal systemic signatures and post-mortem lung data in matched samples, enabling cross-validation and analyses of high-dimensional spatially-resolved single-cell tissue data alongside clinical and peripheral blood data. The study represents the first comprehensive longitudinal investigation of this kind performed in a non-Western or Asian population and it is a proof-of-concept in how patients could be stratified for timely and targeted treatment for a precision medicine approach applied to infectious diseases.

## Data Availability

All data produced in the present study are available upon reasonable request to the authors.

## Acknowledgments

We would like to thank the families who, in times of grief, allowed the autopsies to be conducted and thus provide further insight into COVID-19 disease. Also, we thank all the personnel from the CloroCovid research team, Hospital e Pronto Socorro Delphina Rinaldi Abdel Aziz, RB Patologia, and Departamento de Patologia e Medicina Legal (Universidade Federal do Amazonas) for their assistance with data collection. We would like to thank the support of the team in the Fundação de Medicina Tropical Dr. Heitor Vieira Dourado (FMT-HVD) in Manaus, Brazil. The authors also gratefully acknowledge the help and assistance provided by the Central Laboratory of High-Performance Technologies (LaCTAD, University of Campinas) and the Institute of Infection, Immunity and Inflammation Flow Core Facility and Glasgow Imaging Facility for the generation of some of the data reported in this manuscript. The authors also thank the team of the Veterinary Diagnostic Service of University of Glasgow with preparing FFPE sections at highest standards for this study. We thank Dr Gareth Howell and the Flow Cytometry Core Facility at the University of Manchester for enabling the Imaging Mass Cytometry experiments performed in this study. Purchase of the Imaging Mass Cytometer system was supported by the BBSRC (BB/S019324/1 to K.N.C.). M.M. is supported by Wellcome Trust Center award (number 104111) and Global Challenges Research Fund (GCRF 2020/21). J.LS.F. was supported by the Sao Paulo Research Foundation (FAPESP grant 2019/01578-2 and 2016/12855-9) and it is currently supported by the UK Medical Research Council (MRC) (MR/W018802/1). F.T.M.C is supported by the Sao Paulo Research Foundation (FAPESP grants 2020/05369-6 and 2017/18611-7).

W.M.M. is supported by the Amazonas Research Foundation (FAPEAM N. 005/2020 - PCTI-EMERGESAÚDE – AM). M.V.G.L., G.C.M., W.M.M. and F.T.M.C. are CNPq research fellows. M.P. is supported by the Wellcome Trust grant (206369/Z/17/Z) and UK Medical Research Council grant (MC_UU_00034/9). C.A.M. is a UKRI MRC Future Leaders Fellow (MR/V025856/1). K.N.C. was supported by the MRC (MR/R010099/1). MR/R010099/1 was jointly funded by the UK Medical Research Council (MRC) and the UK Department for International Development (DFID) under the MRC/DFID Concordat agreement and was also part of the EDCTP2 program supported by the European Union.

## Author contributions

J.L.S.F., M.F., G.C.M., M.V.G.L., F.T.M.C., W.M.M., L.C.F., K.N.C., C.M. M.P. and M.M. conceived study concept and design. V.H., M.P.G. and M.H. contributed to study concept. J.L.S.F., V.H., M.P.G., M.H., K.N.C., C.M., M.P. and M.M contributed to literature search, data interpretation, and writing the initial manuscript. All authors contributed to reviewing and editing of the manuscript. M.F., F.F.AV., M.B., C.C.J., T.T. and V.S.S. conducted sample collection and processing. J.L.S.F., V.H., M.P.G., M.H., C.C.J., T.T., V.S.S., C.A., F.M. performed experiments, data analysis and interpretation. J.L.S.F, V.H., M.P.G., and M.M. made figures and tables.

## Lead contact

Further information and requests for resources and reagents should be directed to and will be fulfilled by the lead contact, Joao Silva-Filho.

## Methods

## 1 EXPERIMENTAL MODEL AND SUBJECT DETAILS

### 1.1 Study Cohort

The study was designed to combine longitudinal changes in clinical signatures and peripheral blood biomarkers measured during life (at and during hospitalization) in hospitalized patients with severe COVID-19, with lung-specific pathological responses, which are inaccessible for analyses during life, but are indicative of disease progression. We combined clinical and blood immunological profiling with high resolution tissue imaging and spatially-resolved single-cell proteomics in a prospective cohort of hospitalized COVID-19 patients in the Brazilian Amazon region at the outset of the pandemic (April-July 2020). Samples used were derived from 142 patients admitted and treated at the Delphina Rinaldi Abdel Aziz Emergency Hospital (HPSDRA), in collaboration with the Tropical Medicine Foundation Dr Heitor Vieira Dourado (FMT-HVD), Manaus, Western Brazilian Amazon (Delafiori et al., 2021; Farias et al., 2022; Freire Santana et al., 2020; Santana et al., 2021). This was the largest public reference unit dedicated exclusively to the treatment of severe COVID-19 cases in the state, with an intensive care unit (ICU) capacity of 100 beds. At the beginning of the study, autochthonous SARS-CoV-2 transmission had already been recorded in Manaus, and the city became a major site of SARS-CoV-2 transmission in Brazil within a few weeks. The study was approved by the local Research Ethics Committee at FMT-HVD #CAAE: 30152620.1.0000.0005 and #CAAE: 32077020.6.0000.0005). Severe cases were defined according to the WHO Ordinal scale, by requiring hospitalization for more than 10 days with recovery or death as the outcome, and presenting one or more of the following clinical symptoms: respiratory rate higher than 24 breaths per minute and/or heart rate higher than 125 bpm (in the absence of fever) and/or peripheral oxygen saturation (SpO_2_) ≤ 93% in ambient air and/or shock (i.e., arterial pressure lower than 65 mmHg, with the need for vasopressor medicines, oliguria, or a lower level of consciousness in the last 7 days (Blueprint; WHO). The primary cause of death of the fatal cases was respiratory failure, or sometimes multiorgan failure inclusive of respiratory system failure. Patients were followed from the day of hospital admission up to 28 days of hospitalization, from which 58 had a non-fatal (recovered) outcome and 84 had a fatal outcome (Figure 1A). Complete autopsies were performed on 34 patients.

### 1.2 Hospitalized COVID-19 patients

Hospitalized patients were included if they had clinical and/or radiological suspicion of COVID-19. Suspicion of COVID-19 was defined by the presence or history of fever and any respiratory symptom, e.g., cough or dyspnea and/or ground-glass opacity or pulmonary consolidation observed on a computed tomography (CT) scan. Patients included (18 years of age or older at the time of inclusion) either had SpO2 94% with room air, or required supplementary oxygen, or required invasive mechanical ventilation. Patients were enrolled as soon as laboratory diagnosis for COVID-19 was confirmed via RT-PCR testing of a nasopharyngeal swab sample, as previously described (Santana et al., 2021). Co-prescription of agents hypothesized or proven to be effective in the management or treatment of COVID-19 were logged for all patients with COVID-19. These agents included azithromycin, ceftriaxone, cefalexin, clarithromycin, vancomycin, cefepime, and others. Given that enrolment occurred early in the pandemic prior to the broad recommendation of targeted therapies that may be expected to alter the disease course, patients were not administered medicines such as remdesivir, anti-interleukin-6 (IL-6) receptor monoclonal antibody, anti-coagulants or dexamethasone.

### 1.3 Autopsy

Lung tissues were collected at post-mortem from thirty-four patients, from whom we had also analyzed ante-mortem clinical and blood data, as previously described (Santana et al., 2021). Lung tissue sampling was performed systematically from ten areas of the lungs (Powers, 1995; Santana et al., 2021). The fragments were fixed in 10% neutral buffered formalin, embedded in a paraffin block, and 2-3μm sections were stained with hematoxylin and eosin staining. Special stains for acid-fast bacteria or fungi were made when necessary (Ziehl Neelsen or Grocott-Gomori, respectively). Evaluation of hematoxylin and eosin sections was performed independently both locally in Brazil by two pathologists, and in the UK by two pathologists in the UK. Main pathologic findings were reached by consensus.

Representative FFPE blocks of the main pathologic findings were shipped to the MRC-University of Glasgow Centre for Virus Research for further downstream analysis. Lung samples from autopsy cases occurring before the first COVID-19 case diagnosed in Brazil, were used as non-COVID negative control samples for antibody staining optimizations. Samples showing signs of tuberculosis were excluded from the study (4 cases). The final number of FFPE lung samples (n=30) were then used for characterization and scoring of tissue lesions by H&E, in situ-hybridization (ISH) and immunohistochemistry (IHC) and 11 patient samples were further analyzed by immunofluorescence (IFA) and Imaging Mass Cytometry (IMC).

### 1.4 Clinical phenotyping

#### 1.4.1 Patient demographics

The hospital has all source documents registered online in an electronic healthcare record (EHR) system (Medview). Clinical analyses, laboratory examinations, and routine computed tomography scanning are also available on site. Demographic data, such as age, self-reported sex, weight, and height were recorded at admission.

#### 1.4.2 Patient medical history and risk factors

Smoking, alcoholism, and tuberculosis status was derived from clinical clerking, or direct patient or next-of-kin questioning wherever possible. Pre-existing medical conditions were defined by the study clinical teams using clinical records and patient or relative questioning. Pre-existing conditions were only assigned if they were present at admission, and not if they appeared during hospitalization. After defining pre-existing comorbidities, patients were classified as having the following conditions: hypertension, chronic respiratory disease, chronic cardiovascular disease, chronic kidney disease, liver disease, diabetes, chronic hematological disease, rheumatological condition, dementia, neoplasia, significant immunosuppression and HIV.

#### 1.4.3 Hospital admission and progression timescales

Length of hospital stay (days of hospitalization - DOH) was defined using hospital records for all hospitalized patients. Length of intensive care admission (days in ICU – DICU) and duration of mechanical ventilation (days in mechanical ventilation - DMV) and days of symptoms until outcome (DDOUO), i.e. hospital discharge (recovered cohort) or death (fatal cohort) were defined for hospitalized patients using electronic healthcare records (EHR). All intervention and maximal severity time points were defined according to the date of onset of symptoms for each patient. This was defined by independent clinicians through review of the clinical notes or direct questioning of the patient according to any unusual symptoms related to the current clinical condition.

COVID-19 was defined by presence of at least one symptom consistent with COVID-19 and a positive diagnostic test. Time between symptom onset and sampling was measured in days. In cases where death occurred, this was defined as maximum severity of illness and time between symptom onset and maximum severity (days of disease onset until death - DDOUD) was defined thereafter.

#### 1.4.4 Clinical and peripheral blood data capture

A range of clinical data was collected and stored for downstream analysis using structured methodology from the hospitalized patients included in the study. Here, registered clinical data were grouped as follows: (i) hematological parameters: hematocrit, red blood cell (RBC) counts, platelet counts, lymphocyte counts, neutrophil counts and neutrophil-to-lymphocyte ratio (NCLR); (ii) markers of acute inflammation and tissue injury measured in the blood: C-reactive protein (CRP), lactate dehydrogenase (LDH), creatinine, urea, alanine transaminase (ALT) and glucose. Physiological observations related to lung and heart function were available from EHR for all hospitalized patients, such as SpO2, respiratory rate, heart rate, arterial pressure, capillary filling, corrected QT Interval (QTc) Fridericia and Bazett methods. These clinical parameters were recorded at admission (day 1 - at the same day of hospital admission and no greater than 2 days), days 7, 14 and 28. Between the 7-days intervals, clinical data was also measured and recorded at least once: day 2-6; day 8-13; day 15-27. Blood sampling, for collection of plasma, and naso/oropharyngeal secretion sampling were performed from all cases, which had SARS-CoV-2 infection confirmed by RT-PCR. These samples were collected at admission (day 1) and days 3, 5, 7, 11 and 14 after admission (or of hospitalization). Processed peripheral blood samples (aliquoted plasmas) were shipped to the Wellcome Centre for Integrative Parasitology, University of Glasgow, UK. These samples were used in multiplex-plasma profiling, which was performed using 2 panels **(Table S1)**: for Panel 1 only plasma samples collected at day 1 of project admission from 27 fatal cases were included. Panel 2 included plasma samples collected at hospital admission and up to 14 days (days 1, 3, 5, 7, 11 and 14) from 11 recovered cases and 11 fatal cases. For the fatal cohort, the average time between the last blood sampling/clinical data recording sample and the time of death was of 72h.

#### 1.4.5 Inclusion / exclusion criteria

Patients < 18 years old were not included due to their known lower morbidity/mortality from COVID-19 (Lu et al., 2020). Patients with active malignancy or receiving significant immunosuppression (greater than an equivalent of 40mg once a day of prednisolone) prior to admission, or those with a clear alternative cause for symptoms and hospital presentation were excluded from analyses. For most modalities, samples were prioritized to match for age, sex and severity of illness.

## 2 METHODS DETAILS

### 2.1 Blood sample processing and preparation of poor platelet plasma

Whole blood from hospitalized patients were sampled into EDTA buffered vacutainers (Fisher Scientific) for processing within 4 hours of sampling. Whole blood was centrifuged at 180 g for 19 min at room temperature, without brake, for gradient formation and to obtain a platelet-rich plasma (PRP). PRP was centrifuged at 100 g for 10 min for removal of residual leukocytes, and subsequently centrifuged at 800 g for 20 min to obtain the platelet pellet. Prostaglandin E1 at 300nM was used to minimize platelet aggregation. The supernatant was centrifuged at 1000 g for 10 min to obtain the platelet-poor plasma (PPP)

#### 2.1.1 Multiplex bead array (Luminex) assay

The concentrations of selected proteins in the plasma (peripheral blood biomarkers) were quantified using customized multiplex suspension detection systems (R&D Systems) containing the following 76 analytes (Table S1):

i. EC activation and damage: Angiopoietin-1 (Ang1), Angiopoietin-2 (Ang2), E-Selectin/CD62E, ICAM1, VCAM1, Syndecan-1;
ii. Platelet activation and pro-coagulation markers: ADAMTS13, CD40 Ligand (CD40L), CXCL4/PF4, CXCL7/NAP-2, D-dimer, Fibronectin, IL-11, Thrombopoietin (TPO), Tissue Factor (TF), Plasminogen activator inhibitor-1 (PAI-1), von Willebrand factor A2 (vWF-A2);
iii. Chemokines: CCL1/I-309, CCL2/MCP-1, CCL3/MIP-1α, CCL4/MIP-1β, CCL5/RANTES, CCL8/MCP-2, CCL13/ MCP-4, CCL19/MIP-3β, CCL20/MIP-3α, CCL11/Eotaxin, CCL24/Eotaxin-2, CX3CL1/Fractalkine, CXCL1/GROα, CXCL2/GROβ/MIP-2/, CXCL9/MIG, CXCL10/IP-10, CXCL11/I-TAC;
iv. Myeloid response: Granulocyte colony-stimulating factor (G-CSF), Granulocyte-macrophage colony-stimulating factor (GM-CSF), Macrophage colony-stimulating factor (M-CSF), IL-27, L-selectin/CD62L, Myeloperoxidase (MPO);
v. Cytokines: TNF-α, IL-2, IL-4, IL-5, IL-6, IL-7, IL-8/CXCL8, IL-10, IL-12 p70, IL-13, IL-15, IL-17/IL-17A, IL-21, IL-23;
vi. IL-1 family and Inflammasome response: IL-1α, IL-1β, IL-1RA, IL-18, IL-33;
vii. Type I, II and III Interferons (IFNs): IFN-α, IFN-β, IFN-γ, IFN-λ2, IFN-λ3; viii.Growth Factors: Epidermal growth factor (EGF), Basic fibroblast growth factor (FGFb/FGF2), Hepatocyte growth factor (HGF), Platelet-derived factor AA (PDGF-AA), Platelet-derived factor BB (PDGF-BB);
viii. Cytotoxicity: Fas Ligand (FasL), Complement Component C2, Complement Component C5a, Complement Component C9, Granzyme B;
ix. Immune checkpoint/T-cell exhaustion: Programmed Cell Death Ligand 1 (PD-L1/B7-H1);
x. Hematopoiesis-related cytokines: Osteopontin (OPN), Stem cell factor (SCF/c-kit Ligand).

The assays were conducted according to the manufacturer’s instructions in a Bio-Rad Bio-Plex 200 Systems. Fluorescence intensity (FI) data from the assays were used for further analysis.

### 2.2 Processing and preparation of FFPE lung sessions

#### 2.2.1 Hematoxylin and eosin (H&E) stains

From each FFPE lung sample shipped to the UK, 2-3 µm thick sections were cut and mounted on glass slides. After H&E staining, each lung was scored according to histological criteria applied in other autopsy lung studies in COVID-19 patients (Ackermann et al., 2020; Ashcroft et al., 1988; Barton et al., 2020; Cai et al., 2020; Franks et al., 2003; Gibson-Corley et al., 2013; Klopfleisch, 2013; Magro et al., 2020; Menter et al., 2020; Tian et al., 2020; Wichmann et al., 2020; Xu et al., 2020) on a whole scanned slide (40x, Aperio Versa 8, Leica). The semi-quantitative scoring covered the following parameters or cells: monocyte/macrophage, lymphocyte, plasma cell, neutrophil, eosinophil, megakaryocyte, analysis of the diffuse alveolar damage (DAD), fibrin deposition, alveolar epithelial hyperplasia, thickening of alveolar septae, hemorrhages, microthrombi, arterial thrombi, venous thrombi, leukocytoclastic vasculitis, fibrinoid vessel wall necrosis, vascular amyloidosis (without Congo red), intussusception: angiogenesis, sprouting: angiogenesis, granulation tissue, collagen abundant macrophages, serositis and activated mesothelial cells, squamous metaplasia, broncho-pneumonia, cellular fibromyxoid exudate, bronchial mucosal oedema, alveolar emphysema, alveolar oedema, interstitial oedema, vascular congestion and syncytia and necrotic lung tissue.

#### 2.2.2 Phenotyping by immunohistochemistry (IHC)

Lung sections were stained with antibodies targeting CD3 (1:100; pressure cooking at pH6 buffer antigen retrieval; DAKO/Agilent; cat. number: A0452), CD20 (1:600, pressure cooking at pH6 buffer antigen retrieval; Invitrogen; cat. Number: PA5-16701;) and CD68 (1:200; pressure cooking at pH6 buffer antigen retrieval; DAKO/Agilent; cat. Number: 76550). For detection, EnVision+/HRP, Mouse, HRP kit (Agilent DAKO, cat. Number: K400111-2) or EnVision+/HRP, Rabbit, HRP kit (Agilent DAKO, cat. Number: K4003) were used according to manufacturer’s instructions. Viral nucleocapsid protein was detected using an anti-SARS-CoV-2 nucleocapsid antibody (5μg/ml, 1:200; pressure cooking at pH6 buffer antigen retrieval; Novus Biologicals, cat. Number: NB100-56576) with an EnVision+/HRP, Rabbit, HRP kit (Agilent DAKO, cat. Number: K4003) detection system. Slides were stained in an autostainer (Autostainer Link 48, Agilent Technologies).

#### 2.2.3 Quantification of immune cells

The number of immune cells (CD3^+^: T cells, CD20^+^: B cells and CD68^+^: macrophages) in the tissue were quantified by manually outlining the lung and tuning an algorithm applying the software Image Scope Aperio (Leica) to detect individual immune-positive stained cells. Results are shown as positive cells per cell population in the lung (%).

#### 2.2.4 Detection of viral RNA in FFPE tissue

According to manufacturer’s instructions, viral RNA was detected by RNAscope using a SARS-CoV-2 spike gene-specific probe and a kit (Advanced Cell Diagnostics, 848561 and 322372) as previously described (Gavin R Meehan, 2023; Herder et al., 2021).

#### 2.2.5 Phenotyping by immunofluorescence assays (IFA)

Lung sections were cut onto positively charged microscope slides (Klinipath) and baked overnight at 42°C. For the immunofluorescent co-stains, sections were stained for SARS-CoV-2 nucleocapsid protein first and then either: (i) a marker for endothelial cells (CD31); (ii) macrophages (CD68); (iii) or pneumocytes (Prosurfactant Protein C [SP-C]; Cytokeratin 8 [KRT8]) in a triple stain, using a tyramide dye-based (Akoya) visualisation method. Slides were heated at 60°C for 1 hour to melt the wax around the sections. Sections were cleared by dipping in Xylene twice (5 min each) before rehydrating in decreasing concentrations of ethanol (2 x 100% ethanol, 2 x 90% ethanol, 2 x 70% ethanol; 3 min each) and a final incubation in distilled water (3 min). Sections were circumscribed with a hydrophobic pen, and incubated with endogenous peroxidase inhibitor (Bloxall, Vector Laboratories) for 10 minutes before incubating in Tris Buffered Saline + 0.05% Tween 20 (TBST) for 3 minutes. Slides were then placed in a benchtop autoclave for heat induced antigen retrieval. A small 10-200ul tip box (without lid), was filled with 250ml 1x citrate pH6 antigen retrieval buffer (TCS Biosciences) and slides submerged in the buffer using a metal rack. The tip box with buffer and slides was placed in autoclave, with steam outlet closed and cycle started waiting until the temperature reached 126°C before turning off the autoclave at the main electricity plug. Autoclave was left to cool and re-equilibrate pressure for 10 minutes before opening the valve to relieve the pressure. The tip box was removed carefully and cooled using a running water bath, letting the box float on the water. When cool, the slides were removed from the buffer and placed in distilled water and then TBST for 3 minutes each. In a darkened humidified chamber, blocking solution was added to the sections (2.5% normal horse serum; Vector Laboratories) supplemented with 2.5% normal human serum (Invitrogen)) and incubated for 1 hour at room temperature.

Tapping off the blocking solution, then 5 μg/ml of primary antibody targeting the SARS-CoV-2 nucleocapsid protein (Novus Biologicals, cat. Number: NB100-56576) was added and incubated at 4°C overnight. The primary antibody was tapped off and slides placed in TBST thrice for 3 minutes each time. Neat ImmPRESS HRP Horse anti-Rabbit IgG Polymer (Vector Laboratories) secondary antibody was added to the sections, straight from the bottle, and incubated in the humidity chamber at room temperature for 30 minutes. Slides were then washed again with TBST thrice for 3 minutes. Then TSA Plus-Cy5 (Akoya) reagent was reconstituted with diluent at 1:50 and added to the sections, incubating for 10 minutes at room temperature. Slides were then washed again with TBST thrice for 3 minutes and antigen retrieval repeated, as before, to strip off the previous primary and secondary antibodies but maintain the tyramide dyes. Following the stripping of antibodies, blocking buffer was added as before (1 hour, RT) followed by incubation with primary antibody. Where co-staining for CD31, antibody was added at 2μg/ml (Novus Biologicals, cat.

Number: NB100-2284); for SP-C, antibody was added at 4.17μg/ml (Novus Biologicals, cat. Number: NBP1-60117); and for CD68 antibody was added at 5μg/ml (Agilent, cat. Number: M0876). Slides were incubated in the humidity chamber overnight at 4°C. The next steps of washing and secondary antibody incubation were followed as before using the appropriate ImmPress for each co-stain (mouse for CD68 staining; rabbit for CD31 and SP-C stainings) diluted 1:10 with blocking buffer before addition. Sections were washed before adding the tyramide reagents. For developing the CD31 signal, TSA Plus-Fluorescein (Akoya) was used at 1:200 for 8 minutes and at 1:50 for 10 minutes for CD68 co-stains. Opal 570 (Akoya) was used to stain for SP-C at 1:200 for 6 minutes.

When identifying pneumocytes, a third antibody was also used to visualise KRT8^+^ cells (rabbit anti-human KRT8 used at 2.5ug/ml, Abcam, cat. Number: ab59400). After washing with TBST (3 minutes, thrice), antigen retrieval (citrate pH6) was performed for a third time to strip the second set of antibodies and then the same regime of staining and washing was executed overnight at 4°C, followed by secondary antibody staining (neat rabbit ImmPress) at room temperature for 30 minutes and TSA Plus-Fluorescein at 1:200 for 6.5 minutes. Double or triple co-stains were then carried forward to be counterstained and mounted. After the last tyramide development, slides were placed in TBST for 3 minutes twice, and then once in TBS (no Tween) for 3 minutes. Sections were incubated with 2.5μg/ml DAPI diluted in TBS for 10 minutes at RT in the humidified chamber before placement in TBS for 3 minutes twice. Following rinsing, sections were incubated with TrueView autofluorescent quencher (Vector Laboratories) for 3 minutes, before being placed in TBS for 5 minutes. Sections were mounted with self-hardening Vectashield Vibrance (Vector Laboratories) and coverslipped.

#### 2.2.6 Imaging

Sections were viewed and imaged on a Nikon A1R confocal microscope with Galvo detector or a Leica DiM8 confocal microscope. Images were analyzed using FIJI. Images were analyzed to determine the mean fluorescence intensity (MFI) of SP-C and KRT8 and count the number of co-stained cells (SARS-CoV-2/CD68; SARS-CoV-2/CD31; SARS-CoV-2/SP-C/KRT8) using thresholding to determine positive signal and background staining based on methods from Shihan et al. (Shihan et al., 2021). To quantify the MFI of SP-C and KRT8, three ROIs from patient samples showing the most consistency in terms of staining were analyzed. Three steps in the 15 step Z-stacks which showed the highest intensity were produced as average intensity projections. After splitting the channels, thresholds were defined using FIJI default settings for virus, SP-C and KRT8 signal to generate cell selections with positive signal.

We also used the MFI to quantify the frequency and number of SARS-CoV-2^+^ cells within the epithelial (SP-C^+^ or KRT8^+^ cells), the myeloid (CD68^+^ cells) and endothelial (CD31^+^ cells) compartments as follows: to count infected macrophages, endothelial cells and pneumocytes, maximum intensity projections were produced across the whole of the Z-stack. After splitting the channels, thresholds were defined using FIJI default settings for CD68 (macrophages), SP-C and/or KRT8 (pneumocytes) and virus to define cell selections with positive signal. Following a remerge of channels and the same high levels of contrast for all images to ensure co-localization rather than artefacts, a number of considerations were used to objectively count SARS-CoV-2^+^ cells. Considerations for counting SARS-CoV-2^+^ macrophages included: positive SARS-CoV-2 nucleocapsid signal, positive CD68 signal, morphology and signal surrounding a nucleus. Random speckles or non-cellular signals are not considered genuine SARS-CoV-2^+^ cells. Considerations for counting SARS-CoV-2^+^ epithelial cells (pneumocytes) included: positive KRT8 signal and/or positive SPC signal, positive SARS-CoV-2 nucleocapsid signal, pneumocyte shape/morphology (Type I and II) but not bronchiolar cell morphology and signal surrounding a nucleus. Random speckles or non-cellular signals are not considered genuine infected cells.

#### 2.2.7 Imaging Mass Cytometry (IMC) – Definition of regions of interest

For the high-multiplexed imaging analysis using Imaging Mass Cytometry (IMC), 4 regions of interest (ROIs) of 1.5 μM^2^ in lung sections of 12 fatal COVID-19 cases were selected based on immunohistochemistry and immunofluorescence analysis of subsequent sections. From the H&E analysis, large vessels filled with mononuclear cells and red blood cells (RBCs) were excluded as well as areas showing artefacts due to fixation issues.

#### 2.2.8 Imaging Mass Cytometry (IMC) – Panel

An antibody panel (**Table S1**) was designed to target epitopes specific for the SARS-CoV-2 nucleocapsid protein (Novus Biologicals, cat. Number: NB100-56576), markers to identify epithelial, vascular (endothelial, stromal, and RBCs), myeloid and lymphoid cell types, as well as, markers related to activation and functional states, such as, antigen presentation, apoptosis and cytotoxicity. Primary antibodies were conjugated to lanthanide metals using Maxpar X8 antibody labeling kit (Standard BioTools) as per manufacturer’s instructions.

#### 2.2.9 Imaging Mass Cytometry (IMC) – Preparation and staining

Tissue sections (5 µm thickness) were stained with metal-conjugated antibodies as per manufacturer’s instructions (https://www.standardbio.com/products/instruments/hyperion). Sections were dewaxed in xylene and rehydrated in a graded series of alcohol (ethanol:deionized water 100:0, 90:10, 80:20, 70:30, 50:50, 0:100; 5 min each). In a 95°C water bath, heat-induced epitope retrieval was conducted in Tris-EDTA buffer at pH 8.6 for 30 min. The sections were immediately cooled to 70°C and then blocked with 3% BSA in PBS for 45 min at room temperature. Samples were incubated in humidity chamber overnight at 4°C in primary antibody diluted in PBS, 0.5% BSA. Tissue samples were washed twice with PBS 0.2% Triton X-100, once with PBS, incubated with 191Iridium/193Iridium (1:400 in PBS) for 30 min at room temperature for DNA staining, washed with water and dried before IMC measurements.

#### 2.2.10 Imaging Mass Cytometry (IMC) – Image acquisition

Images were acquired using a Hyperion Imaging System (Standard BioTools) as per manufacturer’s instructions. Regions of interest were laser-ablated in a rastered pattern in a series of 1µm^2^ pixels at a rate of 200Hz. Staining was reviewed using MCD Viewer (Standard BioTools), and all successful image acquisitions were processed and exported for downstream analysis. A total of 58 images were acquired.

## 3 QUANTIFICATION AND STATISTICAL ANALYSIS

### 3.1 Parameters correction using linear mixed effect models

To ensure that the differences observed between patient samples were due to disease progression/outcome and not due to the differences in age, self-reported sex (fixed effects) and days of symptoms at admission (offset) of the patients (random effects), their effect was tested using nested linear mixed models (using the function lmer of the lme4 R package v 1.1.32) (Douglas Bates, 2015), with the following formula: lmer(variable ∼ age + sex + (1 | patient) + offset(days of symptoms at hospitalization), data = dataset). For the cases when not enough samples were available to estimate patient effects, a linear model was used instead with the removeBatchEffect function from the limma R package (v 3.57.6) (Ritchie et al., 2015). For parameters with significant sex or age influence, estimates of predicted age/sex influence (fitted values) were subtracted from the raw parameter values and residuals were used for downstream statistical testing.

### 3.2 Analysis of clinical parameters and peripheral blood (PB) biomarkers

Demographic and temporal data of the study cohorts as well as the concentrations of 76 circulating proteins in plasma measured and presented as the median concentration in pg/mL ± min and max values were analyzed using the following statistical tests. Data normality was checked by the results of the D’Agostino & Pearson, Anderson-Darling, Kolmogorov-Smirnov and Shapiro–Wilk tests. Student’s t-test was used to compare medians between disease outcome (fatal vs recovered) or progression (early death vs late death) with normally distributed data, and data sets with non-normal distributions were compared using Mann–Whitney test. Fisher’s exact test was used for categorical data. All tests were performed two-sided using a nominal significance threshold of p<0.05 unless otherwise specified. When appropriate to adjust for multiple hypothesis testing, Bonferroni, False Discovery Rate or Benjamini–Hochberg (BH), two-stage step-up (Benjamini, Krieger and Yekutjeli) correction methods were applied to test significance at the p-value<0.05 threshold. Analyses were performed and the graphs generated in GraphPad Prism 9 (v 9.5.1 [528], 2023) and RStudio software (v 2023.03.0+386; 2023). Hierarchical clustering, Principal Component Analysis (PCA), for linear dimensionality reduction (Jolliffe and Cadima, 2016; Josse and Husson, 2016), Uniform Manifold Approximation and Projection (UMAP) (McInnes, 2018), for non-linear dimensionality reduction, consensus k-means clustering, and Exploratory Factor Analysis (EFA) (Rouvel and Schaefer, 1990) and machine learning approaches (Ganggayah et al., 2019; Jiang, 2020; Speiser et al., 2019; Tomic et al., 2019; Tomic et al., 2021; Tuleau-Malot, 2022; Wiener, 2002) were used to analyze the clinical parameters and PB biomarkers. To avoid variable-specific bias, the data was preprocessed by applying a scaling to zero mean and unit variance (z-score) to all parameters with the scale function in R. To ensure results reproducibility, we set the seed for the random number generator in all analysis.

#### 3.2.1 Hierarchical clustering and heatmap

First, clinical parameters and PB biomarkers recorded during hospitalization until outcome were clustered using the Ward.D cluster method and Euclidean distance metric. We also ran hierarchical clustering of 27 COVID-19 fatal patients based on the PB biomarkers measured in plasma samples, collected at admission, and combined with the matched clinical data. Here, clustering was repeated 40 times and cluster stability and composition analyzed using reference-based consensus spectral and K-means clustering (k) to identify the best number of clusters using the M3C function of the M3C R package (v 1.18.0), with 25x Monte Carlo iterations, 100x inner replications, objective=’PAC’ and clusteralg = ‘km’ (John et al., 2020). This analysis indicated k=2 generates the most stable clusters (higher jaccard index). The resulting clusters and the z-score values of each parameter were represented in heatmaps (using the function Heatmap of the R package Complex Heatmap v 2.14.0) (Francois Husson, 2017; Gu et al., 2016). The color-scale is bounded at z-score = ± 1, with an increased z-score shaded red; decreased z-score shaded blue; unchanged z-score shaded yellow.

#### 3.2.2 Dimensionality reduction: PCA and UMAP

We carried out principal component analysis (PCA) on the normalized data using the PCA function of the FactoMineR R package (v 2.8) with default parameters (Sebastien Le, 2008). For each principal component (PC), we determined the most significantly associated variables with a given principal component (using the function dimdesc), and the contribution of each variable for each PC (using the function get_pca_var). For visualization of PCA results, ggplot2 (v 3.4.2), factoextra (v 1.0.7), and corrplot (v 0.92) R packages were used. The UMAPs were calculated using the function umap of the umap R package with default parameters.

#### 3.2.3 K-means clustering

Next, we ran reference-based consensus K-means clustering (k) to identify the best number of clusters (for k = 2,3,4,5,6, 7, 8, 9, 10 clusters), using the M3C function of the M3C package in R (with 25x Monte Carlo iterations, 100x inner replications, objective=’PAC’ and clusteralg = ‘km’) (John et al., 2020). This function works by resampling and clustering the dataset using k-means clustering with varying k (i.e., the number of clusters as indicated). Then it calculates a NXN consensus matrix, where each element represents the fraction of times two samples clustered together. This process is repeated 100 times. It also computes several scores to verify the stability of these consensus matrices to decide k, such as the empirical cumulative distribution (CDF), the Proportion of Ambiguous Clustering (PAC) and relative cluster stability index (RCSI). The analysis of these scores indicated k=3 as the optimal number of K-means clusters when patients were clustered based on the longitudinal clinical data only. In parallel, we performed K-means clustering followed by 1000x bootstrapping (using the functions kmeans; clusterboot of the fpc package 2.2.10), which also produced the most stable clusters (highest jaccard_index) with k=3. We then performed an unsupervised consensus k-means clustering analysis of the hospitalized COVID-19 patients based on their longitudinal clinical data on the principal components (using the functions kmeans with the patients coordinates in the PCA plot). The output k-means clusters were also visualized in the UMAP embedding.

#### 3.2.4 Exploratory factor analysis (EFA)

Exploratory factor analysis (EFA) was used to reduce the dimensionality of the clinical and PB biomarkers data into a smaller set of features, without losing information, to identify the driving clinical and PB biomarker “signatures” of variance over time, which we analyzed according to the days of symptoms or days of sampling during hospitalization. This was done by sorting the clinical and PB variables into factors that represent hidden features not measured directly, which here we called clinical and PB signatures, respectively, which is a combination of the variables in some weightage (not essentially adding or removing information in this step but only transforming it). For this, we first analysed the factorability of the dataset by measuring the Kaiser-Meyer-Olkin (KMO) score using the function KMO in the psych R package (v 2.1.9) (Revelle, 2023). According to Kaiser’s guidelines (Dziuban, 1974; Kaiser, 1974), a suggested cut-off for determining the factorability of the sample data is KMO≥60. In our dataset KMO=65 indicating sample adequacy for this analysis. This was confirmed by the Bartlett’s Test of Sphericity using the functions cortest.bartlett and det in the psych R package, which generated very small p-values (p<0.05). Next, we conducted factor analysis using the function fa in the psych R package with default parameters. To determine the number of factors (here called “signatures”) to be retained, we based on the Kaiser’s ‘eigenvalue rule’ of retaining eigenvalues larger than 1 (analysis using a scree plot) and that collectively explain 90-99% of the variance, which means that the new factors explain more variance than one original variables. This analysis resulted in 3 factors (“signatures”) to be retained when reducing the clinical data and 5 factors to be retained when reducing the PB biomarker data. The final factor analysis was then run with these number of factors to be extracted and fine-tuning some parameters in the fa function (changed the type of factor analysis to principal axis factoring with fm=‘pa’; number of iterations for convergence max.iter=100) with other parameters used as default. Resulted factors were sorted in decreasing order of the variances they retain and factor loadings (values of clinical or PB biomarker parameters per each factor) and sample loading scores (values of each patient sample per each factor) were extracted.

The factor loading values of each clinical parameter and PB biomarker per each factor is used to define the parameters that are better represented by a particular factor (which here we called clinical and PB signatures). For instance, clinical parameters with no association with a corresponding signature are expected to have factor loadings close to zero, whereas parameters with strong association are expected to have large absolute loading values. The composition of each clinical and PB signature after sorting parameters across them based on the factor loadings are represented in heatmaps and the mean±SEM of the sample loading scores per patient, grouped according to disease progression (recovered patients in grey, early death in salmon and late death in light blue), are represented along the days of symptoms or days of sampling during hospitalization in line plots. The sign of the sample loading score indicates the direction of the effect: samples with high positive score values indicate that the parameters composing that clinical or PB signature are enriched in those samples, whereas samples with high negative values indicate reduced levels of the parameters composing that particular clinical or PB signature.

#### 3.2.5 Trajectory Inference Analysis

To further characterize the disease progression of hospitalized COVID-19 patients, we used trajectory inference, using the Python packages cellrank (v 1.5.1), scvelo (v 0.2.5) and ehrapy (v 0.2.0) (Bergen et al., 2020; Lange et al., 2022; Lukas Heumos, 2023), to compute the intermediate and terminal macrostates of our patient landscape based on the longitudinal trajectories of combined clinical parameters and PB biomarkers. This analysis also identified the drivers for each trajectory, inferred fate probabilities towards the terminal states for each patient while accounting for the continuous nature of fate determination. Missing data was imputed using the functions estim_ncpPCA and ImputePCA from the missMDA R package (v 1.18), with default parameters and six components maximum to predict the missing entries (ncp=6) (Josse J, 2012; Josse and Husson, 2016). Then, the complete imputed longitudinal clinical and PB biomarker dataset was loaded into a pandas dataframe (fuction read_csv in the pandas Python’s package) was converted into an anndata object (function df_to_anndata in the anndata Python’s package). Next, the data was preprocessed by applying a scaling to zero mean and unit variance to all parameters (function pp.scale_norm in the ehrapy Python’s package). Then, PC and UMAP dimensionality reduction with default parameters and leiden clustering with resolution 1 were calculated (functions pp.pca, pp.neighbors, pp.umap, tl.leiden in the ehrapy Python’s package) and visualisation plots generated (pl.umap). Because with this type of data is not clear to define the cluster of origin in a trajectory, the cellrank’s ConnectivityKernel function was used to compute transition probabilities based on similarities among patients using a KNN graph, by defining a kernel, computing the transition matrix and a projection on top of the UMAP (functions ConnectivityKernel, compute_transition_matrix, compute_projection in the ehrapy Python’s package). To visualize the trajectories forwards in time we used scvelo (function pl.velocity_embedding_stream). Finally, intermediate and terminal macrostates (functions tl.estimators.GPCCA, compute_macrostates, plot_macrostates, set_terminal_states_from_macrostates, plot_terminal_states in cellrank and ehrapy Python’s packages) and the clinical and PB biomarker parameters driving these transitions and terminal states determined and visualized (functions compute_lineage_drivers, plot_lineage_drivers in cellrank and ehrapy Python’s packages).

#### 3.2.6 Machine learning (ML) approaches for building predictive models

We applied machine learning (ML) to search for clinical and PB biomarkers predicting disease outcome (recovered vs fatal) or progression (early vs late death) in hospitalized COVID-19. First, the complete (imputed missing data as described above) dataset containing the longitudinal clinical and PB biomarker measurements from recovered and fatal COVID-19 patients (108 samples in total) was partitioned into 70% training and 30% test set, with balanced class distribution of fatal and recovered patients using the function initial_split from the tidymodels package (v 1.0.0) (Wickham, 2020). Briefly, the dataset is split into groups based on quartiles, and sampling is done randomly within these subgroups to balance the class distributions. Test sets were held out for evaluation of model performance on unseen datasets to prevent overfitting as described previously (Tomic et al., 2019). Next, 171 ML algorithms were trained and tested in the datasets using SIMON (Sequential Iterative Modeling ‘‘Over Night’’) (Tomic et al., 2019; Tomic et al., 2021), to identify the best performing ML models and important variables for feature selection. Since the entire ML process in SIMON is unified, resulting models built with different algorithms can be compared and the best performing models can be selected. First, models are built on training set and the performance is evaluated using a 10-fold cross-validation repeated five times and cumulative error rate is calculated. To prevent overfitting, in the second step, each model is evaluated on the withheld test set. The performance of classification models was determined by standard performance measurements such as accuracy, sensitivity, specificity, precision, recall, area under the receiver operating characteristic curve (AUROC), precision-recall area under curve (prAUC), and logarithmic loss (LogLoss) on training and holdout test sets. After evaluation of these metrics of model performance, the random forest (RF) model was the best-performing ML algorithm on the training as well as on the holdout test set for prediction of disease outcome or progression. In addition, SIMON outputted the contribution of each clinical and PB biomarker feature to the model as variable importance score (scaled to maximum value of 100). Next, another feature selection step for prediction, to cross-validate SIMON outputs, was performed now using RF-based algorithms (randomForest v 4.7.1, randomForestExplainer v 0.10.1, LongituRF v 0.9 and VSURF v 1.2.0 packages in R) (Capitaine et al., 2021; Ganggayah et al., 2019; Jiang, 2020; Speiser et al., 2019; Tuleau-Malot, 2022; Wiener, 2002). First, we performed fine tuning of the RF model by determining: (i) the best number of trees (ntree); (ii) the best number of variables randomly sampled at each stage (mtry); (iii) the best number of maxnodes and the K-fold cross validation. The final RF model (mtry=5, maxnodes = 15, ntree=1000, nodesize=5) was trained in 70% of the samples (randomly assigned) and performance evaluated on the test dataset (30% of the samples). Using VSURF, the three steps of variable selection procedure for supervised classification and regression problems based on the fine-tuned random forest model (functions VSURF, VSURF_interp and VSURF_pred) were run to build predictive models for disease outcome (recovered vs fatal) or progression (early death vs late death). These steps comprehend a first “thresholding step”, where RFs are computed using the function randomForest with arguments importance=TRUE and the values for ntree and mtry as initially trained. Then variables are sorted according to their mean variable importance (VI), in decreasing order and a minimum threshold (min.thres), i.e. based on the predicted value of a pruned CART tree fitted to the curve of the standard deviations of VI, is computed. Next, the actual “thresholding” is performed, where only variables with a mean VI larger than nmin * min.thres are kept. The “interpretation step” aims to select all variables related to the response (here disease outcome or progression) for interpretation purpose. The variables passing the threshold in the first step are embedded in RF models that grow starting with the random forest build with only the most important variable and ending with all variables selected in the first step. Then, the minimum mean out-of-bag (OOB) error (err.min) of these models and its associated standard deviation (sd.min) are computed. The smallest set of corresponding variables is selected when then mean OOB error is less than the product of err.min + nsd * sd.min. Finally, the third step (”prediction”) refines the selection for prediction purpose, by eliminating redundancy in the set of variables passing the interpretation step. Variables are added to the RF model in a stepwise manner and the mean jump value (mean.jump), the mean absolute difference between mean OOB errors of one model and its first following model, is calculated. Variables are included in the final model if, when they are left out, the mean OOB error decrease is larger than the product nmj * mean.jump. The output of VSURF feature selection was cross-validated again by measuring variable importance based on permutation, impurity or loss function in RF models trained using the functions randomForest::importance and measure_importance in the randomForestExplainer R package.

Next, we evaluated the practical value of the selected clinical parameters and PB biomarkers parameters in predicting disease outcome or progression when measured at or early after hospital admission. For this, we used only the clinical and PB biomarker predictors measured at and up to 3 days of hospitalization. Here, missing data was imputed using proximity from randomForest with the function rfImpute in the randomForest package in R. The predictive RF model for disease outcome was built with the following formula: randomForest(Outcome∼IL12p70+CCL1+IFNlλ2+PD-L1+IL33+IFNβ+Creatinine+Platelets, data=train_data, importance=T, ntree=1000).

The predictive RF model for disease progression was built with the following formula: randomForest(Outcome∼PD-L1+CCL1+LDH+Creatinine,data=train_data, importance=T, ntree=1000). The performance of the models was calculated from a confusion matrix with all performance metrics, including the area under the receiver operating characteristic curves (AUROC), computed using the R packages pROC (v 1.18.0) and ROCR (v 1.0.11) (Robin et al., 2011; Sing et al., 2005). The RF model performed with excellent accuracy when predicting disease outcome (AUROC train set 0.97; test set 1.00) and with lower but good accuracy when predicting disease progression (AUROC train set 0.93; test set 0.82). Examples of decision trees were plotted (using the function plot.getTree in the reprtree v 0.6 R package) and assisted in establishing the cut-off values for specific clinical and PB biomarker parameters measured up to 3 days of hospitalization (plotted with ggplot) to exemplify how these parameters could assist in prediction and patient stratification in the clinical setting with the aim to evaluate best therapeutic strategies.

### 3.3 Imaging Mass Cytometry (IMC) analysis

#### 3.3.1 Preprocessing and imaging denoise

We have employed a modular computational workflow to process and analyze IMC data, using a combination of Python and R packages (Geuenich et al., 2021; Greenwald et al., 2022; Jackson et al., 2020; Lu et al., 2023; Martinelli and Rapsomaniki, 2022; Palla et al., 2022; Rendeiro et al., 2021; Vito RT Zanotelli, 2022; Wolf et al., 2018). First, image data, containing the raw acquisition data for multiple regions of interest (ROIs), optical images providing a slide level overview of the tissue section to be sampled, panoramas, and detailed experiment metadata, were extracted from MCD files acquired with the Fluidigm Hyperion instrument. Images were converted to OME-TIFF and single- and multi-channel TIFF files and (using the ImcSegmentationPipeline in Python) (Vito RT Zanotelli, 2022). Hot pixels artifacts were first filtered during the conversion to multi-channel TIFF files using a default threshold (hpf=50). Each pixel intensity is compared against the maximum intensity of the 3×3 neighboring pixels and if the difference is larger than this threshold, the pixel intensity is clipped to the maximum intensity in the 3×3 neighborhood. Next, to improve the signal-to-noise ratio and remove artifacts, we employed an automated content-aware pipeline performing 2 steps in each of the single-channel TIFF files: (i) it deploys a differential intensity map-based restoration (DIMR) algorithm to detect and remove hot pixels; (ii) then, the pipeline runs a self-supervised deep learning algorithm for filtering shot or background noise (DeepSNiF) (IMC-Denoise Python’s package) (Lu et al., 2023). The denoised single-channel TIFF files were restacked and used in downstream analysis. Denoised images were visualized with cytomapper R package (v 1.9.2) (Nils Eling, 2020).

#### 3.3.2 Cell segmentation and extraction of single-cell features

Cell segmentation was done using a fully automated deep learning-based segmentation approach (DeepCell using the ark-analysis Python package) (Greenwald et al., 2022). This pipeline enables high-throughput segmentation and accurately resolves individual cells across diverse tissues and structures. Briefly, the user first aggregates specific image channels to generate two-channel images representing nuclear and membrane signals. All lineage and functional markers underwent a staining quality check for selection of channels to be used in the cell segmentation. Next, the ark-analysis Python package is used to run Mesmer, a deep learning-enabled segmentation algorithm pre-trained on TissueNet, to automatically obtain cell segmentation masks (Greenwald et al., 2022). Segmentation masks are single-channel TIFF images that match the input images in size, with non-zero grayscale values indicating the IDs of segmented cells. To compare and validate imaging segmentation using the automated deep-learning approach, we also used a semi-automated pixel classification-based approach by training a pixel classifier (Jackson et al., 2020; Vito RT Zanotelli, 2022). Pixels are classified as nuclear, cytoplasmic, or background with ilastik (v 1.4.0), based on a trained random forest classifier using features derived from the image and its derivatives (Berg et al., 2019). Image features used were the Gaussian smoothing, Laplacian of the Gaussian, Gaussian gradient magnitude, difference of Gaussians, structure tensor eigenvalues and the Hessian of Gaussian eigenvalues, each of which had Gaussian kernels of widths from 1to 10 (30 features in total). The outputs of prediction are probability maps for each pixel, which were used to segment the images using a customized CellProfiler (v 4.2.4) pipeline (Stirling et al., 2021; Vito RT Zanotelli, 2022). First the probability map stack is split into nuclear, cytoplasm/membrane and background and using the image-math module an image with the sum of nuclear+cytoplasmic signal is generated. The ‘IdentifyPrimaryObjects’ module is used to segment the nuclear masks to identify nuclei. Next, the ‘IdentifySecondaryObjects’ module is run with the nuclei and the nuclear+cytoplasma image to identify cells and gaps in the identified cells are filled. Ambiguous pixels are removed after imaging rescaling. Finally, segmentation masks are generated for analysis. Following image segmentation, using the segmentation masks together with their corresponding multi-channel images, features of the segmented cells were quantified (using the ark-analysis Python package). These features include the mean pixel intensity of each protein per segmented cell, morphological features, such as cellular area, eccentricity, major and minor axis length and ratio, perimeter, convex area, diameter and cellular spatial coordinates in the X and Y axis (centroid-X, centroid-Y).

#### 3.3.3 Single-cell analysis: pre-processing and cell assignment

For the single-cell analysis, the dataset output from the single-cell feature extraction step was converted into an anndata object. The presented data were not transformed, and all analyses were based on raw IMC measurements. Single-cell marker expressions are summarized by mean pixel values for each channel. The single-cell data were normalized at the 99th percentile to remove outliers, and z-scored cluster means visualized in heatmaps. For PhenoGraph Louvain the data were normalized to the 99th percentile, as is suggested for this clustering algorithm (Levine et al., 2015). We used Scanpy (Single-Cell Analysis in Python v 1.9.1) (Wolf et al., 2018) to perform PCA, batch-correct and integrate the data from each ROI using BBKNN (batch balanced k nearest neighbours) (Polanski et al., 2020), and to compute a UMAP embedding (umap-learn Python package, v 0.5.3) (McInnes, 2018). Next, we performed automated cell type assignment using the Python package Astir (ASsignmenT of sIngle-cell pRoteomics v 0.1.4) (Geuenich et al., 2021). The workflow uses as input an expression matrix containing the measured expression data (anndata) and a priori specified set of marker proteins for all expected cell types to be found (yaml file). Then it employs a statistical machine learning model (deep recognition neural networks) to assign cell type probabilities to each cell. The algorithm assumes cell types express their marker proteins at relatively higher levels than other cell types and it includes a set of post-fit diagnostics to ensure all cell types express their marker proteins at significantly higher levels than other cell types and flags all marker and cell type combinations for which this is violated. Cells that co-express improbable marker combinations are automatically assigned as “other” (for example, co-expression of PanCK and CD3), while cells with a high probability of belonging to more than one cell type are assigned as “unknown”. Assignment to these cell types is, in part, caused by non-specific staining of the individual samples or mis-segmentation of neighboring cells. For cell assignment with Astir, the following information to label cells based on a broad ontogeny (metaclusters and major cell types) and the proteins (lineage markers) to be most expressed in each expected cell type were used. Metaclusters and major cell types: (a) myeloid: macrophage, neutrophil; (b) lymphoid: CD8 T cells, CD4 T cells, B cells; (c) vascular: Endothelium, red blood cells (RBCs); (d) stromal: fibroblast, smooth muscle cell, epithelial. Cell types: (a) macrophage: CD163, CD206, CD14, CD16, CD68, CD11c, Iba1; (b) neutrophil: CD66b, Arginase1; (c) CD8 T cells: CD3, CD8; (d) CD4 T cells: CD3, CD4; (E) B cells: CD20; (f) endothelium: CD31; (g) fibroblast: Collagen1; (h) smooth muscle cell (SMC): SMA; epithelial: PanCK; RBCs: CD235ab.

#### 3.3.4 Single-cell analysis: clustering and annotation of cell identities

After cell assignment, cells labelled as “other” or “unknown” were filtered out from downstream analysis, the anndata object was subset into the major cell types identified, i.e., myeloid, lymphoid, vascular and stromal and Phenograph Louvain clustering (with 200 nearest neighbors) (Levine et al., 2015) was performed for each cell population separately using a small set of specific lineage marker and functional proteins The clusters were then manually annotated to define the populations based on a ranking for the highly differential expressed protein markers in each cluster using Wilcoxon rank-sum (Mann-Whitney-U) and Benjamini-Hochberg corrected adjusted p-values. Annotated cell identities were visualized in heatmaps, UMAP and their spatial coordinated. Next, clusters were merged, resulting in cell type resolutions at two different depths - one to define major cell types (as defined with Astir), and a finer one to define subpopulations (as defined through Phenograph clustering).

#### 3.3.5 Single-cell analysis: differential abundance analysis

The finer annotation was used to evaluate the frequency and absolute counts of cell types. The number and frequency of cells per image (ROI), or patient and disease progression were normalized by the image area (total number of pixels) and displayed as cell frequency or number/mm^2^ and age and/or sex corrected as indicated using linear mixed models as described above. Data normality was checked by the results of the D’Agostino & Pearson, Anderson-Darling, Kolmogorov-Smirnov and Shapiro–Wilk tests. Student’s t-test was used to compare medians between disease progression (early death vs late death) with normally distributed data, and non-normal distributions were compared using Mann–Whitney test. Adjusting for multiple comparisons was applied with the Bonferroni or Holm-Šídák test methods. All tests were performed two-sided using a nominal significance threshold of p<0.05 unless otherwise specified.

Differential abundance analysis was also performed using the miloR R package (v 1.4.0) (Dann et al., 2022), which performs testing by assigning cells to partially overlapping neighborhoods on a k-nearest neighbor (KNN) graph. The workflow includes the following steps: (a) Construction of the KNN graph, computed based on similarities (the Euclidean distance between each cell) in protein expression (log-transformed) and its k nearest neighbors in the principal component (PC) space; (b) Definition of cell neighborhoods; (c) Counting cells in neighborhoods to construct an N x S (neighborhood × experimental sample count) matrix; (d) Testing for differential abundance in neighborhoods, by determining neighborhood counts using the quasi-likelihood (QL) method in edgeR (Lun et al., 2017). In brief, the method fits a negative-binomial generalized linear model (NB GLM) to the counts for each neighbourhood, accounting for different numbers of cells across samples using a trimmed mean of M values (TMM) normalization (Robinson and Oshlack, 2010), and use the QL F-test for the contrast early death vs late death to compute a p-value for each neighborhood; (e) Controlling the spatial FDR in neighborhoods, interpreted as the proportion of the union of neighborhoods that is occupied by false-positive neighborhoods, for multiple testing correction, as previously described (Lun et al., 2017). The method applies a weighted version of the Benjamini–Hochberg method, where the reciprocal of the neighborhood connectivity is used to weight p-values.

Neighborhood connectivity is measured by the Euclidean distance to the kth nearest neighbor of each cell for each neighborhood. The outputs are used to construct a graph where nodes represent neighborhoods, edges represent the number of cells in common among neighborhoods and the size of nodes represents the number of cells in the neighborhood. This graph is visualized in the UMAP plot, where the nodes are positioned accordingly to the position of the sampled index cell in the embedding, which allows qualitative comparison with the single-cell embedding labelled with the cell identities (**Figure S6A**). Finally, we used the R package ggplot for graphical visualization of the log fold-change in the abundance of each cell type in early death vs late death patients (Figure 5B).

#### 3.3.6 Spatial statistics analysis

Spatial statistics analysis based on the coordinates of the cells in the ROIs, were performed using the Python packages Squidpy (Spatial Quantification of Molecular Data in Python v. 1.2.2) (Palla et al., 2022), Athena (Analysis of Tumor HEterogeNeity from spAtial omics measurements v 0.1.3) (Martinelli and Rapsomaniki, 2022), SpOOx (Spatial Omics Oxford Pipeline) (Praveen Weeratunga, 2022) and the R package Giotto (v3.3.0) (Dries et al., 2021). First step was the construction of the spatial graph, which is the graph representation of each cell spatial neighbors, with cells as nodes and neighborhood relations between cells as edges. Here, a generic graph (coord_type = ‘generic’) was created by defining a fixed radius of 20μM (radius = 20) from the centroid of each cell of interest (from Scikit-learn (Pedregosa, 2011), function gr.spatial_neighbors in the Squidpy Python’s package). The spatial analysis is focused on a radius = 20μm because it approximates the distance between the centroids of cells that are in physical contact. After the construction of the spatial graph, neighborhood enrichment scores were determined and visualized (functions gr.nhood_enrichment and pl.nhood_enrichment in Squidpy; pl.spatial in Scanpy Python’s packages). This score computes the spatial organization of each cell type in a quantitative way to inform on the neighbor structure of the tissue, as previously described (Palla et al., 2022) In brief, this score is the result of a permutation-based test determining whether cell types interact more or less frequently compared to a random distribution by permuting cell type labels 1000 times (Schapiro et al., 2017). To derive such a distribution, cell labels are randomized 1000 times, and for each iteration the interaction count is computed. Statistical inference is performed by comparing the actual interaction count to the empirical null distribution: statistical significance for avoidance is defined if more than 990 iterations of random permutations produce larger counts, while statistical significance for association is defined if more than 990 iterations of random permutations produce smaller counts. The z-scores of the neighborhood enrichment of all cell types identified in the post-mortem lung sections with SARS-CoV-2^+^ alveolar macrophages and SARS-CoV-2^+^ epithelial cells were extracted for downstream integrative analysis. In Giotto, the spatial graph was constructed using the function createSpatialNetwork (method = knn, k =4 and maximum_distance_knn = 20), which generated the same results as with the Squidpy function. To identify cell types that are found to be enriched in a spatially proximal manner, as a proxy for potential cell-cell interactions, we use a random permutation (default n = 1000) strategy of the cell type labels within a defined spatial network using the function cellProximityEnrichment. Similar to the function in Squidpy, this function calculates the ratio of observed-over-expected frequencies between two cell types, where the expected frequencies are calculated from the permutations. Then p-values are calculated by observing how often the observed value were higher or lower than the simulated values for respectively increased or decreased frequencies. Next, the function findInteractionChangedFeats was applied to identify all potential protein expression changes associated with specific cell-type interactions in an unbiased manner. For each cell type, the cells are split into two complementary subgroups, with one containing the subset which neighbor cells from another specific cell type. Differential expression is determined by a spatial permutation test followed by adjust for multiple hypothesis testing using a background null distribution reshuffling the cells within the same cell type (Dries et al., 2021). The SpOOx pipeline was also applied to further validate the statistically significant spatial enrichment, which applies a 3-step spatial association analysis, quadrat correlation matrices (QCMs), cross-pair correlation functions (cross-PCFs) and adjacency cell network (ACN) (Praveen Weeratunga, 2022). The first step, QCM, identifies statistically significant cell co-occurrences (FDR < 0.05) based on correlations of the absolute numbers of cell pairs within square quadrats of up to 100uM. The second step, cross-PCFs, examines whether significantly correlated cell pairs (in the range 0-100uM) are in proximity within a radius g(r) (here equals to 20μm) above a spatial randomness. Values of cross-PCF higher than 1 indicates that cell type 1 is observed more frequently at distance r from cell type 2 than expected under complete spatial randomness, and values lower than 1 is indicative of avoidance or exclusion. Finally, the ACN step determines whether the co-locating cell pair is in physically contact with each other.

#### 3.3.6 Publicly available IMC COVID-19 lung data

Publicly available raw IMC COVID-19 data from 7 post-mortem lung samples (79 regions of interest) of a north-American fatal cohort were downloaded from the following: https://zenodo.org/deposit/4139443. Clinical annotations from these patients were obtained from the supplementary information in the published dataset (Rendeiro et al., 2021). The raw IMC dataset was fully re-analyzed and cell identities annotated following the workflow described in the methods sections 3.3.1-3.3.5.

### 3.4 Integration of systemic and tissue data using systems biology and machine learning approaches

#### 3.4.1 Multi-factor analysis (MFA)

To investigate the differences and similarities between patients from a multidimensional point of view, as well as the correlation between variables, we first applied Multiple Factor Analysis (MFA) to integrate clinical data, peripheral blood biomarker profiling and tissue features (based on IMC) from fatal cases (using the function MFA in the missMDA v 1.18 R package)(Josse and Husson, 2016). In brief, MFA consists in performing a global PCA on the dataset concatenating the weighted matrix of each source of variables (Pagès, 2014). Here, we applied MFA with the following aims: (i) to investigate the differences and similarities between patients from a multidimensional point of view, as well as the correlation between variables; (ii) to highlight similarities and differences between groups of variables, i.e., pointing out what is common between the trajectories in disease progression and what is specific; (iii) finally, MFA was also used to balance the influence of the groups of variables (clinical, PB biomarkers and tissue features extracted from the IMC) in the analysis in such a way that no single group, with correlated variables for instance, dominates the top dimensions of variability. For this, the MFA approach calculates for each group of variables (systemic and tissue), a principal component (PC) and then each PC value in the group is divided by the square root of the first eigenvalue. The following tissue features were extracted from IMC and used as inputs: (i) frequency and absolute counts of different cell types; (ii) scores of neighborhood enrichment of cell populations with SARS-CoV-2^+^ alveolar macrophages and SARS-CoV-2^+^ epithelial cells; (iii) expression levels of markers related to antigen presentation (MHCI, MHCII, CD74), cytotoxicity (CD107a, Granzyme B), apoptosis (ClvCaspase3) and endothelial cell activation (ICAM-1, vWF) in SARS-CoV-2^+^ alveolar macrophages, SARS-COV-2^+^ epithelial cells and activated ECs.

Datasets of clinical, PB biomarker and the tissue features extracted from the IMC were merged using shared variables, such as assay-specific sample or patient ID. To indicate the source of the assay of each variable, they were grouped as clinical, PB biomarker or IMC. Samples were ordered based on the day they were collected. We standardized names of the cell types to reflect the measurement, such as frequency (Freq_cell type), counts (Count_cell type) and interacting pairs (cell type_cell type). The final dataset included 53 samples from 9 hospitalized COVID-19 fatal patients following the early death progression and 7 patients following the late death progression with 17 clinical parameters, 72 PB biomarkers (measured by Luminex) and 113 tissue features (analyzed using IMC). The data for each assay was centered and scaled and missing values were imputed using the function imputeMFA (missMDA R package).

#### 3.4.2 MOFA/MEFISTO and survival analysis

##### 3.4.2.1 Method rationale

We used the approaches in Multi-Omics Factor Analysis (MOFA) and Method for the Functional Integration of Spatial and Temporal Omics data (MEFISTO) (Argelaguet et al., 2020; Argelaguet et al., 2018; Velten et al., 2022) to integrate clinical data, peripheral blood biomarker profiling and tissue features (based on IMC) from fatal cases. In brief, this is another computational method we used for integrating multiple modalities of data in an unsupervised fashion for discovering of the principal sources of variation (here we called as “integrated signatures”) in the distinct disease progressions of hospitalized COVID-19 patients with fatal outcome. The method was designed for integrating data modalities via a common sample space (i.e., measurements derived from the same set of samples), where the features (variables) are distinct across data modalities (approaches).

Given several data matrices with measurements of parameters from different approaches on the same or on partially overlapping sets of samples, MOFA infers an interpretable low-dimensional data representation in terms of (hidden) factors (“integrated signatures”); then it disentangles to what extent each factor is unique to a single data modality or is representative of variation from multiple modalities, thereby revealing shared axes of variation between the different approaches’ layers. We added MEFISTO into the model to account for the temporal dependencies between samples that result from factor analysis, as we included longitudinal clinical and PB biomarker data. MEFISTO decomposes the high-dimensional dataset with longitudinal measurements from multiple approaches and groups of samples into a small number of factors in a time-aware manner. This temporally informed dimensionality reduction enables more accurate and interpretable recovery of the underlying patterns of variation by leveraging known temporal dependencies rather than by solely relying on correlations between features (Argelaguet et al., 2020; Argelaguet et al., 2018; Velten et al., 2022).

##### 3.4.2.2 Create object

The first step is the generation of the MOFA object. For this, datasets containing longitudinal clinical, PB biomarker and the tissue features extracted from the IMC were merged in a long data frame object and samples metadata, such as disease progression, day of sampling during hospitalization, days of symptoms at sampling and the source of the parameter measured indicated as clinical, PB biomarker or IMC were added. To avoid variable-specific bias, the data was preprocessed by applying a scaling to zero mean and unit variance to all parameters, using the scale function in R. To ensure results reproducibility, we set the seed for the random number generator in all analysis. The MOFA object was created using the function create_mofa (MOFA2 R package v1.8.4) with default parameters. The group function as used as default (group=NULL) as our aim was to find the systemic and tissue parameters that differentiates the early and late death progression. Because one of the aims is to identify the temporal trajectories of the integrated signatures, as we are integrating the longitudinal clinical and PB biomarker data, the analysis should be run in a time-aware manner. For this, the time points of the averaged (within each disease progression group) day of symptoms at sampling or day of sampling during hospitalization were added into the model by setting the function set_covariates accordingly. This function adds continuous temporal data as covariate to the MOFA object for training with MEFISTO (Velten et al., 2022). A similar approach was applied to integrate the clinical, PB protein marker and lung tissue IMC data from the Amazonian and north American cohorts (Rendeiro et al., 2021). Batch effects from the IMC data and differences in sex and age compositions were regressed out using a linear model with the removeBatchEffect function from the limma R package (v 3.57.6) (Ritchie et al., 2015) before scaling. Because the north American is a cross-sectional cohort, in this analysis only clinical parameters and PB protein markers data from the last sample collected before time of death from patients in the Amazonian fatal cohort were used.

##### 3.4.2.3 Define options, prepare and train model

This step provides a default set of data, model and training options that can be modified and passed to the MOFA object before preparing the object for training. We used default parameters in the function get_default_data_options, the number of factors was set with to num_factors = 5 in the function get_default_model_options, the convergence mode was set to “slow” in the function get_default_training_options and default parameters were used in the function get_default_mefisto_options (MOFA2 R package v1.8.4). The MOFA algorithm implements an Automatic Relevance Determination prior, which automatically learns the effective number of factors. Hence, although we specified the starting number of factors to 5, factors that do not explain any variation will be pruned during model inference (Argelaguet et al., 2020). All defined model options were passed to the object in the preparing step with the function prepare_mofa. Finally, the model was trained with the function run_mofa (MOFA2 R package v1.8.4) and the output saved as a hdf5 file. Before visualisation, missing values in the model were imputed based on the MOFA model using the function impute (MOFA2 R package v1.8.4). The imputed data is then stored in the imputed_data slot of the MOFA object and accessed via the get_imputed_data function.

##### 3.4.2.4 Model quality control, visualization and interpretation

After model training, each factor (here called “integrated signatures) captures a different source of variability in the data, as they are defined by a linear combination of the input features. A good sanity check for model quality control is to verify whether the factors are largely uncorrelated, which confirms a good model fit, the chosen number of factors in the model is appropriate and the normalization was adequate. The variance decomposition by factor (or integrated signature) was also evaluated, as it summarizes the percentage of variance explained by each factor across each data modality (clinical, PB biomarker and IMC) from the heterogeneous dataset. Then, the total variance explained per approach (by combining all factors) is another important metric of quality control of the model as high variances (in %) implies that the factors capture most of the variation for the corresponding approach, whereas values <10% means that the variation is not explained by the model i.e. it is considered as noise with strong non-linearities. Importantly, because MOFA generates a linear and sparse model, which prevents overfitting, it will never explain 100% of the variance in the data, even if using a lot of factors.

Next, the weights of each parameter per source of data (clinical, PB biomarker or tissue features from IMC) per each factor (integrated signature) was inspected and plotted. The weights provide a score for each variable on each factor (integrated signature). Parameters with a strong association with the factor (integrated signature) are expected to have large absolute values: the sign of the weights indicates the direction of the effect, a positive weight indicates that the feature has higher levels in the samples with positive factor (integrated signature) values, and vice-versa. Finally, groups with different signs manifest opposite phenotypes along the inferred axis of variation, with a higher absolute value indicating a stronger effect. Note that the interpretation of MOFA factors is analogous to the interpretation of the principal components in the MFA.

Because the model is generated with a temporal covariate, the longitudinal trajectories of the factors (integrated signatures) can be visualized by plotting the factor loading score of each patient sample against the days of symptoms, days of sampling during hospitalization, and colour coded based on the disease progression. Samples with different signs manifest opposite phenotypes along the inferred axis of variation, with higher absolute value indicating a stronger effect.

##### 3.4.2.5 Building predictive models of clinical outcome

The factors inferred by MOFA can be related to clinical outcomes such as survival times. For this, we used Cox proportional hazards model (Crowley and Breslow, 1984) to estimate the hazard of death as a function of the integrated signatures as covariates. If an integrated signature has a influence on the survival time, it will show a high absolute coefficient in the Cox model. If the coefficient is positive, samples with large values of the integrated signature have an increased hazard of death (low survival probability) compared to samples with smaller values for that integrated signature. To fit these models, we used the function Surv (survival R package v3.5.5) to creates a survival object for use as the response in a model formula considering the MOFA factors and the patients’ days of disease onset until death (DDOUD) with the the coxph function. The survival data is stored in a survival object that contains both the time a sample has been followed up and whether the event has occurred (as 0,1). Kaplan–Meier plot along with log-rank tests (using the functions ggsurvplot and survdiff in the survival v3.5.5 R package) were conducted to assess the differences in survival probabilities between the high and low levels of the integrated signature with p-value < 0.05 in the Cox regression analysis across the days of disease timeframe (Rich et al., 2010).

Aiming at translating discoveries into a clinically actionable assay that is broadly accessible, we also used the systemic and tissue parameters from the MOFA integrated signatures to determine the minimum number of clinical and/or PB biomarkers that could be used to predict progression by using random forests to build predictive models without compromising accuracy. Thus, based on the ranking of factor weights in the integrated signature 2, we combined the top two, three, four or five clinical and/or PB biomarkers measured up to 3 days of hospitalization and tested combinations to predict progression in our validation dataset with high accuracy. Missing data was imputed using proximity from randomForest with the function rfImpute in the randomForest R package (v 4.7.1). The final predictive RF model for disease progression based on the MOFA output (small panel) was built with the following formula: randomForest(Outcome∼Creatinine+CRP +Urea+IL11+E-selectin+TPO, data=train_data, importance=T, ntree=1000). The performance of the models was calculated from a confusion matrix with all performance metrics, including the area under the receiver operating characteristic curves (AUROC), computed using the R packages pROC (v 1.18.0) and ROCR (v 1.0.11)(Robin et al., 2011; Sing et al., 2005). Examples of decision trees were plotted (using the function plot.getTree in the reprtree v0.6 R package) and assisted in establishing the cut-off values for specific clinical and PB biomarker parameters measured up to 3 days of hospitalization (plotted with ggplot) to exemplify how these parameters could assist in prediction and patient stratification in the clinical setting with the aim to evaluate best therapeutic strategies.

#### 3.4.3 Sparse Decomposition of Arrays (SDA)

The third approach used for integration of data from the different approaches (clinical, PB biomarker from luminex, and tissue features from IMC) was the Sparse Decomposition of Arrays), a method for matrix and tensor decomposition, which similar to MOFA/MEFISTO, also fits a latent underlying structure of factors (also called components), explaining the patterns of variability in the data (Chang et al., 2021; Fanaee and Thoresen, 2019; Hore et al., 2016). Datasets of clinical, PB biomarker (measured by Luminex) and the tissue features extracted from the IMC, were merged using shared variables, such as assay-specific sample or patient ID. To indicate the source of the assay of each variable, they were grouped as clinical, PB biomarker or IMC. Samples were ordered based on the day they were collected. We standardized names of the cell types to reflect the measurement, such as frequency (Freq_cell type), counts (Count_cell type) and interacting pairs (cell type_cell type). In total, the integrated dataset contained information on 97 samples from 9 hospitalized COVID-19 fatal patients following the early death progression and 7 patients following the late death progression with 17 clinical parameters, 72 PB biomarkers (measured by Luminex) and 113 tissue features (analyzed using IMC). Missing values were imputed using the function imputeMFA (missMDA R package v 1.18) (Josse and Husson, 2016) and the dataset was converted to a converted to a 4-dimension array, comprising of 16 samples, 202 variables, 3 approaches, 6 time points, using the base R function array). We ran the bayesian tensor and marix decomposition 2000 times for 4 components and setting stopping=FALSE, using the function RunSDA4D (from the SD4D R package v 0.1) (Marchini, 2020). The algorithm can shrink components to zero, so in the output, the algorithm selects the number of components when the number of estimated components is less than the true number. Similar to the defined method (Marchini, 2020), the evidence lower bound (ELBO) was calculated for each iteration, achieving its maximum level at around 200 iterations. After the model was run, we extracted the outputs consisting of a list of the approximate posterior distributions of the main parameters, such as the values of each component (factor) per patient sample (individual Scores matrix A), the scores of each approach per component (matrix B), the scores of each component over time (matrix D) and the values of each component per variable (matrix WS). Components were identified as associated with COVID-19 progression if they showed significant variation determined by Welch’s t-test or Mann–Whitney test using a nominal significance threshold of p<0.05. The overview heatmap (Figure S11A) was generated by combining and applying hierarchical clustering of the variables in the top significant (p < 0.05) components differentiating early and late death COVID-19 progression.

#### 3.4.5 Correlation analysis

Spearman’s rank correlations coefficients and corresponding p-values were calculated with the function rcorr in the hmisc R package (v 5.0.1). Adjusted p-values of the correlation coefficients were calculated using the function rcorr_padjust in the tabletools R package (v 0.1.0). The correlation coefficients and adjusted p-values were visualized using the function corrplot displaying positive correlations in red and negative correlations in blue. Asterisks represent correlations with adjusted p values: *p<0.05, **p<0.01, ***p<0.001.

---

**Table S1:** Panels of multiplex profiling using a bead-based assay (Luminex) and panel information for the IMC experiments.

**Figure S1:**
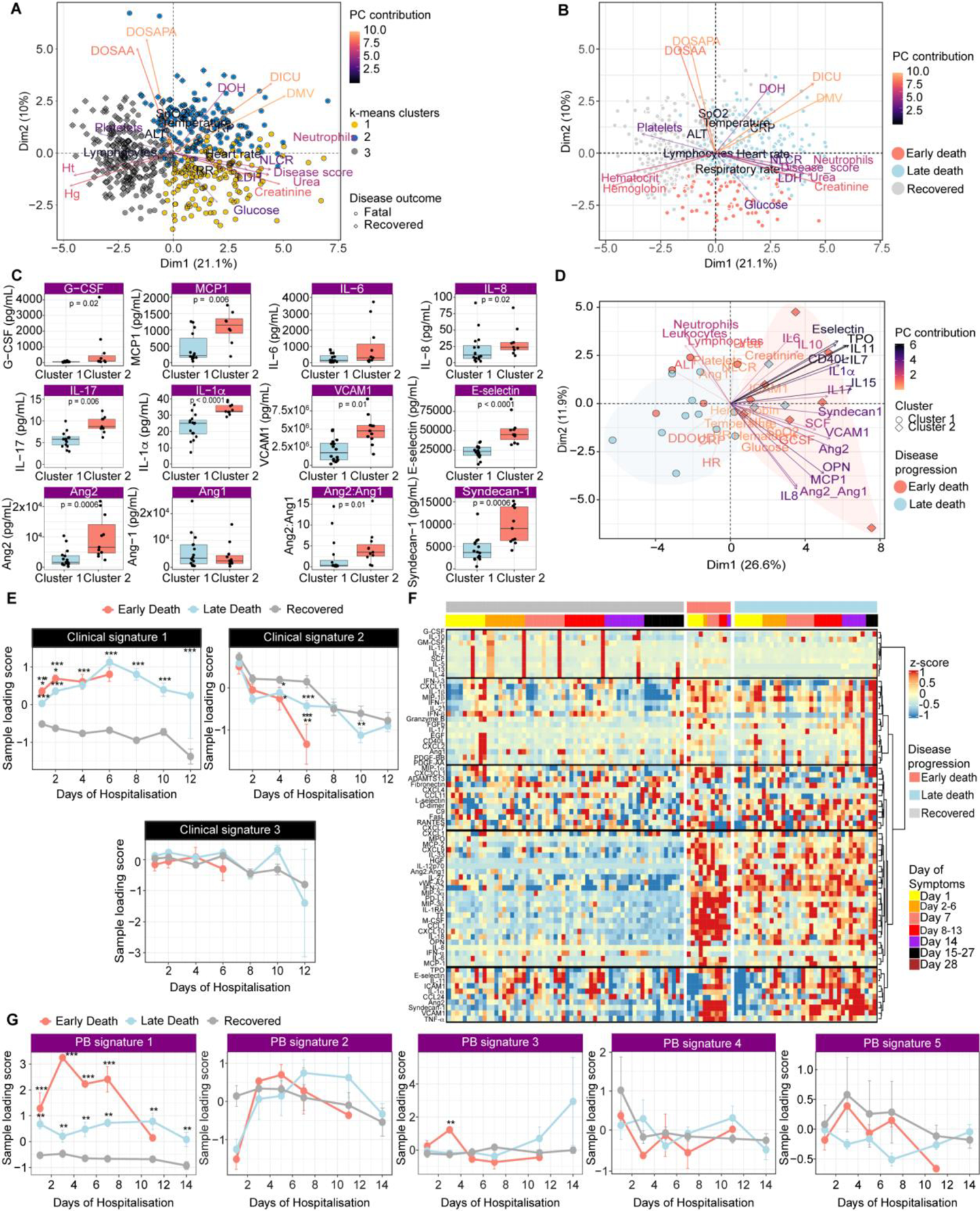
Peripheral blood biomarkers and clinical data measured at hospital admission in severe COVID-19 fatal patients identifies distinct disease trajectories associated with the days of symptoms until death. **(A,B)** PCA biplot of longitudinal clinical variables showing patients’ samples color-coded by unsupervised k-means clustering (left panel) and by disease progression (right panel). Principal component analysis (PCA) shows the relationship between the clinical parameters measured during hospitalization with the individual measurements in recovered (grey dots) and fatal (blue dots) COVID-19 patients. The level of the contribution of the clinical variables, in accounting for the variability in each principal component (Dim1 and Dim2) are colour coded and their directionality in the PCA plot are represented by the arrows. Each dot represents a sample from each patient collected up to 28 days of hospitalization and colour coded by the k-means cluster **(A)** and by disease progression **(B)**. In A, dots are surrounded by a circle, representing patients with fatal outcome, or by a diamond shape, representing patients with recovered outcome. **(C)** Peripheral blood (PB) proteins, measured by the Luminex panel 1, driving the separation between the hierarchical clusters 1 and 2 of hospitalized COVID-19 patients with fatal outcome. Bar plots represent the absolute levels of the proteins enriched in the PB of COVID-19 fatal patients in the hierarchical cluster 2, measured on samples collected at hospital admission. Statistical differences are displayed where observed with adjusted p-values using one-way ANOVA with correction for multiple comparisons (Tukey’s Method) or non-parametric (Kruskal-Wallis) statistical tests with the Dunn’s post hoc tests. Bars indicate the median measurement and the min/max values. **(D)** PCA biplot representing the severe COVID-19 fatal patients accordingly to the early and late death groups. Dimensionality reduction analysis representing the 2 clusters identified by hierarchical clustering analysis. The level of the contribution of the variables in accounting for the variability in each principal component (Dim1 and Dim2) are color coded and their directionality in the PCA plot are represented by the arrows. Each dot represents a patient sample (collected at hospitalization) and color coded accordingly to the corresponding cluster. COVID-19 fatal patients in the cluster 2 (colored in light red) show higher plasma levels and are enriched in markers of EC activation and damage (VCAM-1, E-selectin, Syndecan-1, Ang-2, Ang-2/Ang-1 ratio); myeloid activation and chemoattraction (G-CSF, MCP-1, IL-8) and other pro-inflammatory cytokines and coagulopathy markers (IL-17, IL-1α, TPO, IL-11, CD40L). **(E)** Graph representation of the clinical signatures, determined by based on Exploratory Factor Analysis (EFA), in hospitalized COVID-19 recovered (grey line), early (light red line) and late death (light blue line) progression according to days of sampling during hospitalization. Lines represent the longitudinal trajectory in variation of each clinical signature during hospitalization for each disease progression (recovered in grey, early death in light red and late death in light blue). Dots represent averaged (within disease progression group) loading scores of each patient sample and error bars the standard error. Significance was tested using unpaired t-test with multiple comparisons adjusted by false-discovery rate (FDR) calculated by the two-stage step-up method (Benjamini, Krieger and Yekutjeli). *q-value < 0.05, ** q-value < 0.01, *** q-value < 0.001. **(F)** Longitudinal peripheral blood analysis of proteins (luminex panel 2) in a subset of hospitalized COVID-19 recovered and fatal patients, the latter grouped in early and late death progression. Hierarchical clustering of the peripheral blood (PB) biomarkers measured by multiplex plasma profiling (Luminex Panel 2) in the samples longitudinally collected up to 14 days of hospitalization. The fatal patients were grouped based on the early and late death progression. Disease progression and the day of sampling are indicated by the bars on the top of the heatmap. **(G)** Graph representation of the PB signatures, determined by based on Exploratory Factor Analysis (EFA), in hospitalized COVID-19 recovered (grey line), early (light red line) and late death (light blue line) progression according to days of sampling during hospitalization. Lines represent the longitudinal trajectory in variation of each PB signature during hospitalization for each disease progression (recovered in grey, early death in light red and late death in light blue). Dots represent averaged (within disease progression group) loading scores of each patient sample and error bars the standard error. Significance was tested using unpaired t-test with multiple comparisons adjusted by false-discovery rate (FDR) calculated by the two-stage step-up method (Benjamini, Krieger and Yekutjeli). *q-value < 0.05, ** q-value < 0.01, *** q-value < 0.001.

**Figure S2:**
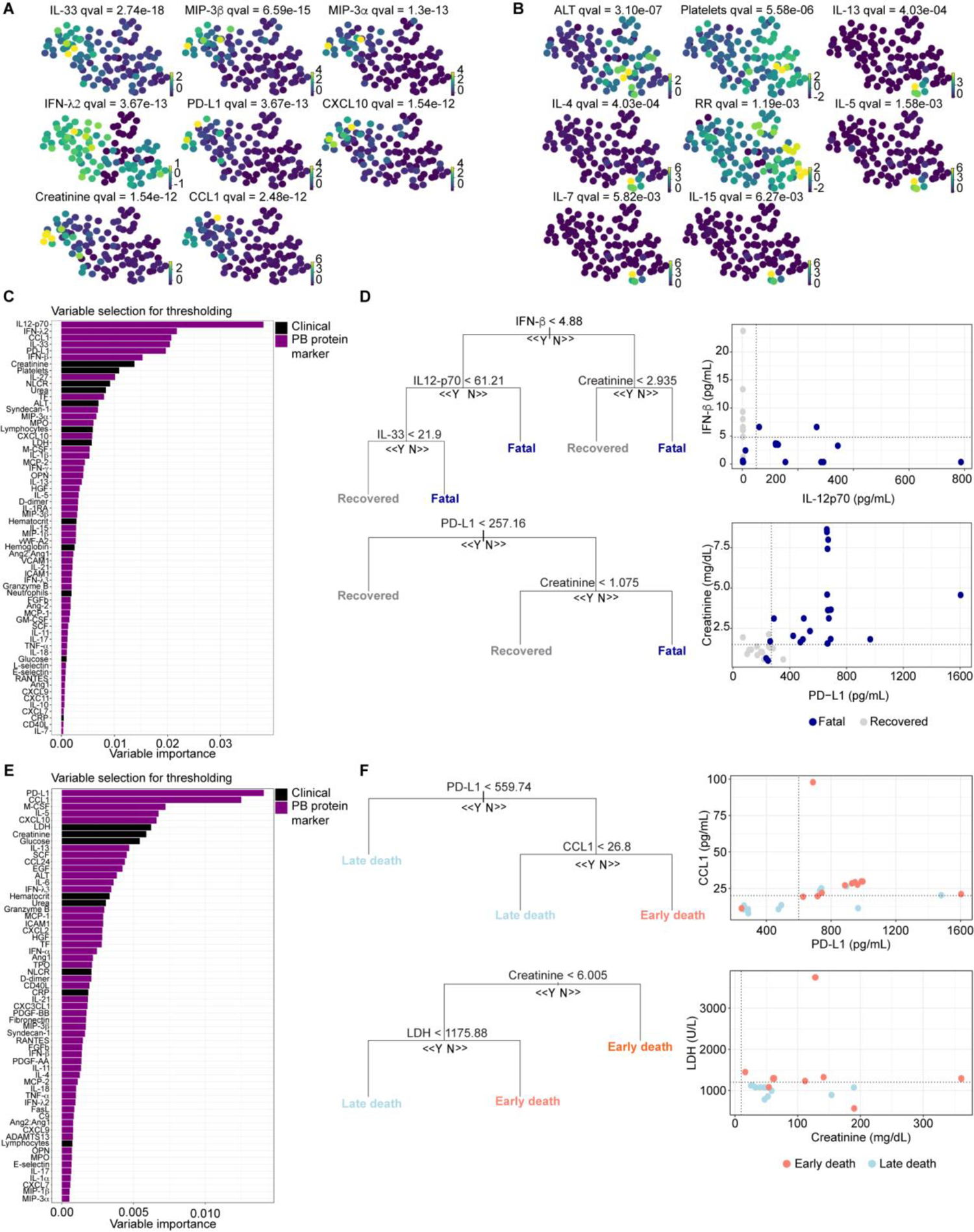
Machine learning models identify clinical parameters and PB biomarkers predicting fatal outcome and disease progression in hospitalized COVID-19 patients. **(A)** Clinical parameters and PB biomarkers driving the terminal state predominantly of early death patients. The main clinical and PB biomarkers driving the terminal states of early death patients were plotted in the UMAP embedding. Dots, representing each sample, are color coded based on the z-score values of each parameter and corrected p-values (qval) for the correlations of these parameters to this terminal state are indicated in the top of each UMAP plot. Analysis of the major clinical and PB protein markers driving these terminal states corroborated the EFA outputs. Protein markers of myeloid activation and chemoattraction and inhibition of T-cell responses in the PB signature 1, such as IL-33, MIP-3α, MIP-3β, CXCL10, CCL1 and PD-L1, and clinical signature 1 (creatine) were significantly correlated with trajectories towards the terminal state characterizing the early death progression. **(B)** Clinical parameters and PB biomarkers driving the terminal state predominantly of recovered patients. The main clinical and PB biomarkers driving the terminal states of recovered patients were plotted in the UMAP embedding. Dots, representing each sample, are colour coded based on the z-score values of each parameter and corrected p-values (qval) for the correlations of these parameters to this terminal state are indicated in the top of each UMAP plot. Parameters in the PB signature 5 (IL-4, IL-5, IL-13, IL-7, IL15), related to Th2 response and lymphopoiesis, were significantly correlated with trajectories towards the terminal state characterising the recovered progression. **(C)** For feature selection, clinical parameters and peripheral blood biomarkers are ordered in descending order of their importance based on mean decreased accuracy or OOR error rate in RF model predicting disease outcome. **(D)** Example decision tree (left panels) based on the top 6 variables important for prediction of disease outcome measured up to 3 days of hospitalization. Cut-off values of the attribute that best divided groups were placed in the root of the tree according to the parameter value. Scatter plots (right panels) representing how the cut-off values of the paired parameters, measured up to 3 days of hospitalization, can separate patients based on the predicted disease outcome. To evaluate the practical value of the 6 parameters in predicting disease outcome when measured at or early after hospital admission, the RF model was trained in 70% of the dataset and then evaluated in a test dataset, using only the clinical and PB biomarker predictors measured at and up to 3 days of hospitalization. The decision trees and cut-off values for specific clinical and PB biomarker parameters measured up to 3 days of hospitalization exemplify how these parameters could assist in prediction and patient stratification in the clinical setting with the aim to evaluate the best therapeutic strategies. **(E)** For feature selection, clinical parameters and peripheral blood biomarkers are ordered in descending order of their importance based on mean decreased accuracy or OOR error rate in RF model predicting disease progression. **(F)** Example decision tree (left panels) based on the top 4 variables important for prediction of disease progression measured up to 3 days of hospitalization. Cut-off values of the attribute that best divided groups were placed in the root of the tree according to the parameter value. Scatter plots (right panels) representing how the cut-off values of the paired parameters, measured up to 3 days of hospitalization, can separate patients based on the predicted trajectory of disease progression. To evaluate the practical value of the 4 parameters in predicting disease progression when measured at or early after hospital admission, the RF model was trained in 70% of the dataset and then evaluated in a test dataset, using only the clinical and PB biomarker predictors measured at and up to 3 days of hospitalization. The decision trees and cut-off values for specific clinical and PB biomarker parameters measured up to 3 days of hospitalization exemplify how these parameters could assist in prediction and patient stratification in the clinical setting with the aim to evaluate the best therapeutic strategies.

**Figure S3.**
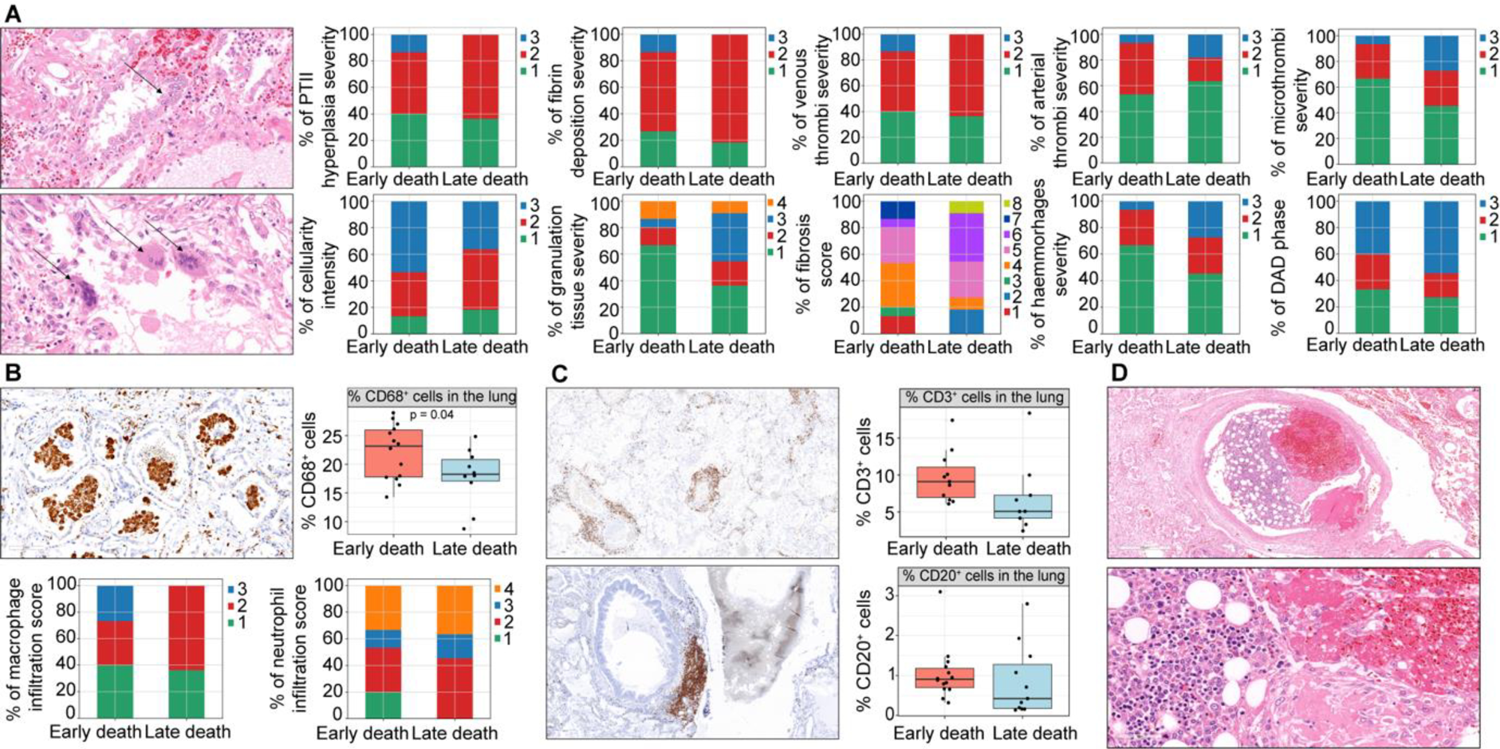
Cellular infiltration and relationship of peripheral blood signatures and lung tissue lesions according to the disease trajectories leading to fatal outcome. **(A)** Frequency of the severity or intensity of the histopathological lesions identified and scored in the lung of fatal COVID-19 patients according to the disease progression. Pathognomonic histological findings of post-mortem lungs from COVID-19 fatal cases are prominent pneumocyte type II (PTII) hyperplasia (arrow in the top left image indicates focal area of proliferated type II penumocytes), syncytia (multinucleated cells - arrows in the bottom left image), vascular thrombi, the so called cellular fibromyxoid exudate, the alveoli obscuring the lumen and hyaline thrombi. According to the enrichment of morphological signatures 1 and 3 in the lung of fatal patients following the early death progression, higher levels of fibrin deposition, venous thrombi, pneumocytes Type II hyperplasia and immune cell infiltration (cellularity) are observed in the lungs of these patients. Meanwhile, accordingly to the enrichment of morphological signatures 3, 4 and 5, fatal patients following the late death progression present in the lung higher levels of fibrosis, hemorrhages, microthrombi, arterial thrombi and granulation tissue. For each parameter, score 1 means rare occurrence (< 5%) in the section analyzed, score 2 = multifocal (6-40%) occurrence, score 3 = coalescing (41-80%) and score 4 = diffuse (>80%) occurrence. For the fibrosis score, 1 = minimal fibrous thickening of alveolar or bronchiolar walls; 2-3 = moderate thickening of walls without obvious damage to the kung architecture; 4-5 = increased fibrosis with definite damage to the lung structure and formation of fibrous bands; 6-7 = severe distortion of structure and large fibrous areas; 8 = total fibrous obliteration of the field. **(B)** Intensity of infiltration of CD68^+^ (myeloid) cells in the post-mortem lung of fatal COVID-19 patients according to the disease progression. For each parameter, score 1 means rare occurrence (< 5%) in the section analyzed, score 2 = multifocal (6-40%) occurrence, score 3 = coalescing (41-80%) and score 4 = diffuse (>80%) occurrence. **(C)** Intensity of infiltration of CD3^+^ and CD20^+^ lymphoid cells in the post-mortem lung of fatal COVID-19 patients according to the disease progression. For each parameter, score 1 means rare occurrence (< 5%) in the section analyzed, score 2 = multifocal (6-40%) occurrence, score 3 = coalescing (41-80%) and score 4 = diffuse (>80%) occurrence. **(D)** Representative case of extramedullary hematopoiesis in the post-mortem lung.

**Figure S4:**
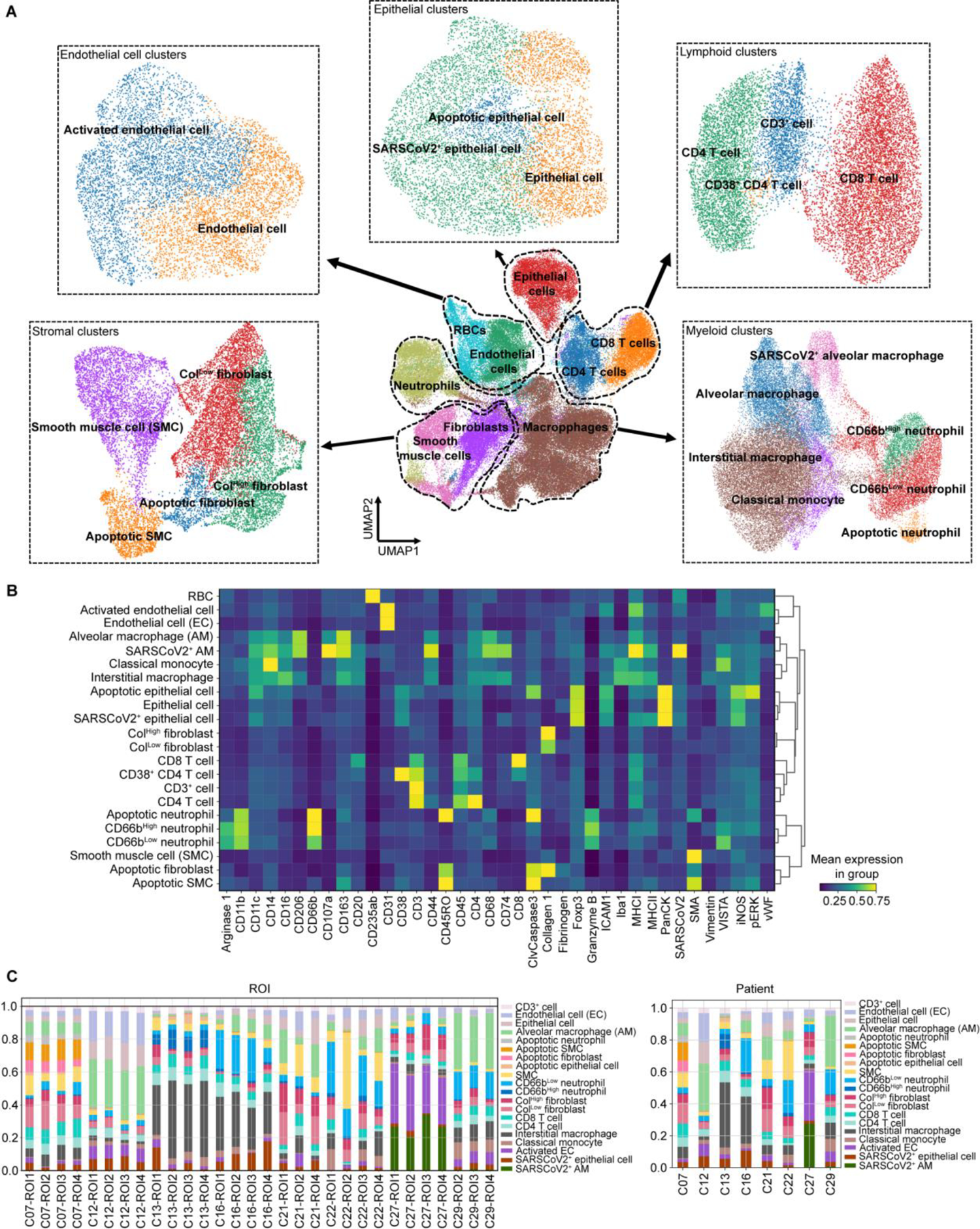
Major cell types in the post-mortem lung tissue of fatal COVID-19 patients determined by Imaging Mass Cytometry (IMC). **(A)** UMAP embedding of the cell types identified in the lung samples by IMC. UMAP embedding of the major cell types identified in the lung samples after supervized cell assignment, using the ASTIR Python’s package. Through ASTIR, we identified and represent in the UMAP embedding epithelial, stromal, vascular, lymphoid and myeloid cell types. Each major cell type was clustered, using Phenograph Louvain clustering, and resulting clusters were annotated and merged to extract the final set of cell types for downstream analysis. The final clusters, for each major cell type, used as an input for the spatial statistics analysis are shown in dashed boxes. **(B)** Heatmap representing the phenotype of the cell types identified in the lung samples. The plot shows the mean expression levels of each marker protein in the IMC panel in each cell type identified in the lung tissue. **(C)** Stacked bar plots showing the frequency of the different cell types across ROIs (left panel) and each patient case (left panel).

**Figure S5:**
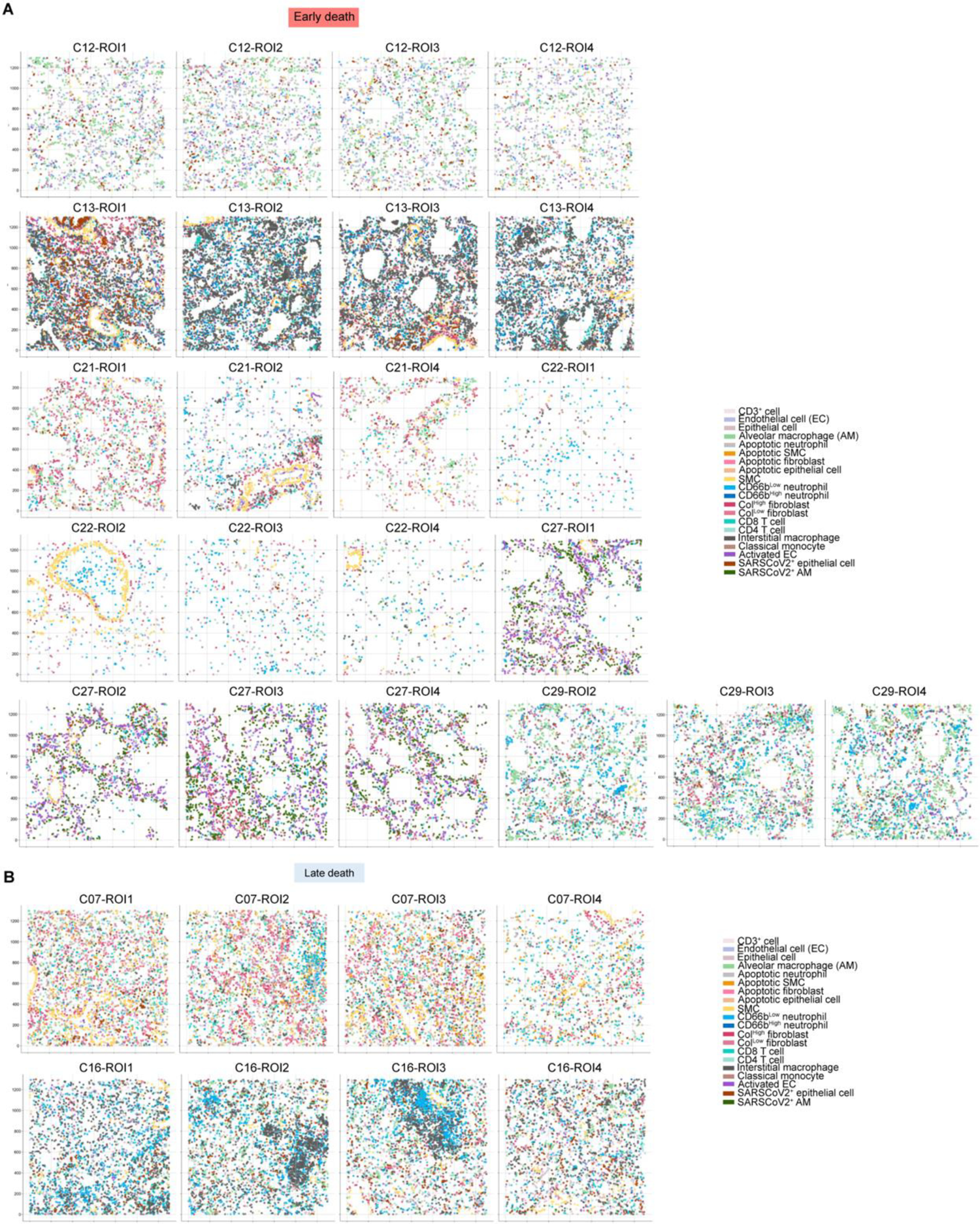
Visualization of the cellular landscape according to the spatial coordinates of cells in regions of interest (ROIs) in sections of post-mortem lung from fatal COVID-19 patients. **(A)** are ROIs from early death progression cases. **(B)** are ROIs from late death progression cases.

**Figure S6:**
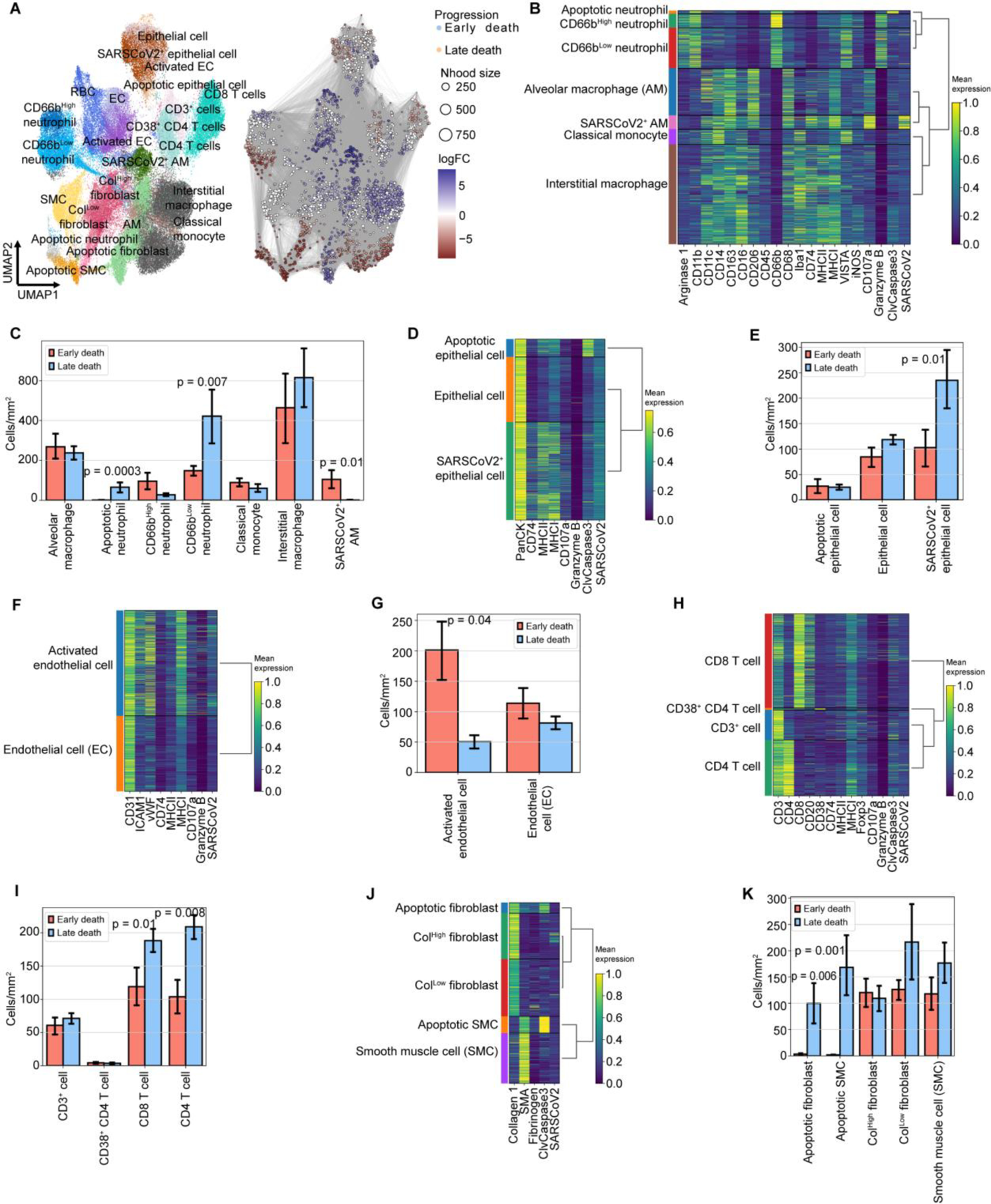
Phenotype, frequency and absolute numbers of the cell types identified by IMC in the post-mortem COVID-19 lung autopsies. **(A)** Independent clustering performed by Milo, in which neighborhood groups are shown for early and late death cases (left UMAP panel). Annotation of each neighborhood group based on our analysis is shown on the right panel. The logarithm of the fold-change comparing early death to late death is represented on the vertical axis. **(B)** Heatmap representing the mean expression of marker proteins used in the Phenograph clustering of myeloid cells. The mean expression values of the marker proteins were used to determine the merging of similar cell clusters and their annotation, as shown in the lateral rows of the heatmap. **(C)** Absolute numbers of the different myeloid cells according to the disease progression. **(D)** Heatmap of the mean expression of marker proteins used in the Phenograph clustering of epithelial cells. The mean expression values of the marker proteins were used to determine the merging of similar cell clusters and their annotation, as shown in the lateral rows of the heatmap. **(E)** Absolute counts of the different epithelial cells according to the disease progression. **(F)** Heatmap of the mean expression of marker proteins of the endothelial population. The mean expression values of the marker proteins were used to determine the merging of similar cell clusters and their annotation, as shown in the lateral rows of the heatmap. **(G)** Absolute counts of the different endothelial cells according to disease progression. **(H)** Heatmap of the mean expression of marker proteins used in the Phenograph clustering of lymphoid cells. The mean expression values of the marker proteins were used to determine the merging of similar cell clusters and their annotation, as shown in the lateral rows of the heatmap. **(I)** Absolute counts of the different lymphoid cells according to disease progression. **(J)** Heatmap of the mean expression of marker proteins used in the Phenograph clustering of stromal cells. The mean expression values of the marker proteins were used to determine the merging of similar cell clusters and their annotation, as shown in the lateral rows of the heatmap. **(K)** Absolute counts of the different stromal cells according to disease progression.

**Figure S7:**
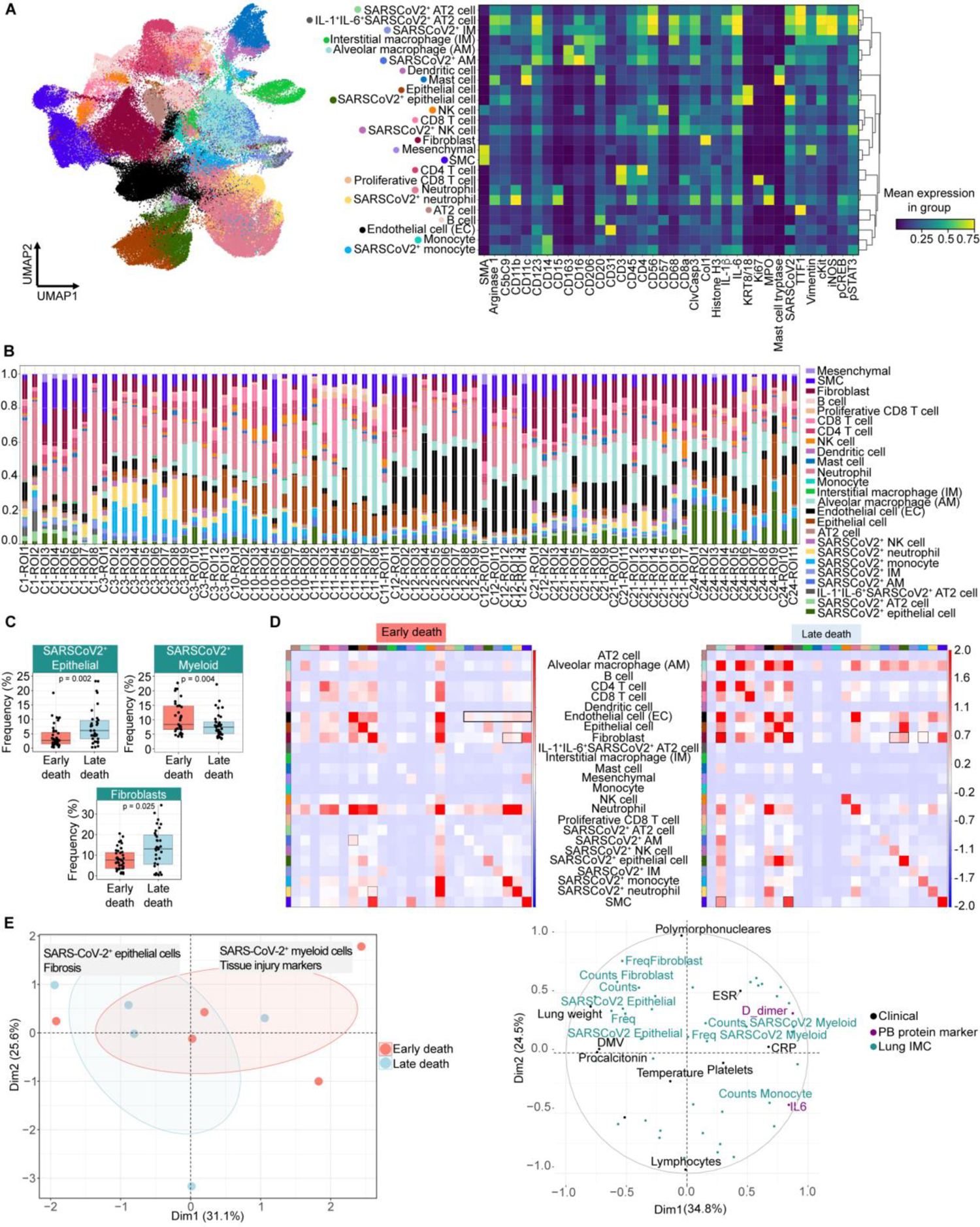
IMC Analysis of post-mortem lung from a North American fatal COVID-19 cohort (Rendeiro et al.). **(A)** UMAP embedding and phenotype of the cell types identified in the post-mortem lung ROIs from a North American fatal COVID-19 cohort (Rendeiro et al.). Clusters were determined using Phenograph Louvain clustering on each of the major cell types identified using the ASTIR Python’s package, such as myeloid, lymphoid, endothelium, stromal and epithelial cells. Each cluster was annotated based on expression levels of specific marker proteins, followed by merging of clusters of similar phenotypes. The UMAP embedding (left panel) shows the cell types identified. The heatmap (right plot) shows the mean expression levels of each protein marker in the IMC panel for each cell type identified in the post-mortem lungs. **(B)** Frequency of cell types in each ROI. Red light shading indicates the ROIs from early death cases. Blue light shading indicates the ROIs from late death cases. **(C)** Frequency of SARS-CoV-2+ cell types and fibroblasts according to disease progression. **(D)** Spatial neighborhood enrichment analysis in ROIs from early (left panel) and late death (right panel) cases. **(E)** Clinical and tissue signatures driving variance between early and late death progression. We used Multiple Factor Analysis (MFA) to integrate the clinical parameters, PB biomarkers and lung tissue features extracted from the IMC analysis. In the dimensionality reduction plot (left panel), each dot represents a patient and they are color-coded accordingly to the disease progression (light red, early death; light blue, late death). The ellipses (color coded accordingly to disease progression) represent point concentration ellipses with their size defined by the 95% confidence interval around the group mean points. A summary label of the underlying factors driving the early and late fatal progression are indicated in the boxes. In the dimensionality reduction plot (right panel), the underlying clinical parameters (in black), PB biomarkers (in purple) and tissue features (in green) driving the early and late fatal progression in this cohort are indicated. The data indicate that signatures contributing to the variance in the first principal component are the main determining factors in segregating the patients following the early death progression from those following the late death progression.

**Figure S8:**
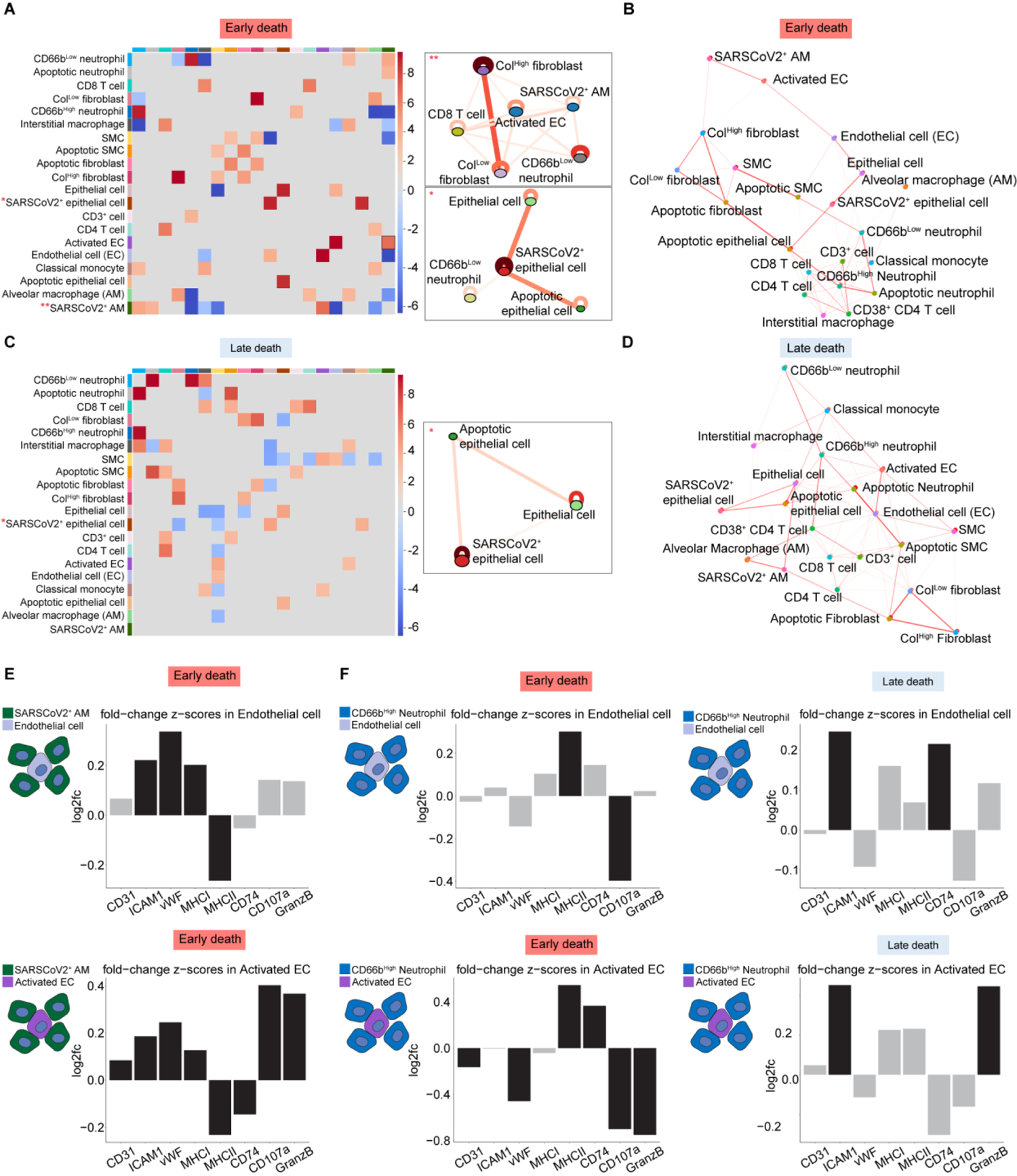
Different spatial statistics analyses in the post-mortem lung of COVID-19 early and late death patients. **(A)** Different spatial statistics methods were applied to quantify proximal homotypic (within cell-types) and heterotypic (between cell-types) cellular interactions and to generate disease progression-specific interaction maps. Heatmap (in the left) showing statistically significant interacting pairs of cells within a 20μM radius from the anchor cell type 1 in the early death lungs, as determined by the SpOOx pipeline. Red boxes indicate cell types with significant (FDR < 0.05) interactions (enrichment proximity), while blue boxes indicate cell types with significant (FDR < 0.05) avoidance. Spatial connectivity plots (in the right) for SARS-CoV-2^+^ alveolar macrophages (SARSCoV2^+^ AM) and SARS-CoV-2^+^ epithelial cells (SARSCoV2^+^ epithelial cell) show the cells that are significantly co-located with them within a 20μm radius [g(r) > 1], represented by the connecting lines, as determined by the cross-pair correlation analysis (cross-PCF) in the SpOOx pipeline. **(B)** Network representation of the pairwise interacting cell types identified by Giotto in the early death lungs. Only enriched interactions, depicted by the red lines, are shown. Width of the edges indicates the strength of spatial enrichment. **(C)** Heatmap (in the left) showing statistically significant interacting pairs of cells within a 20μM radius whiting the anchor cell type 1, in the late death lungs. Spatial connectivity plots (in the right) for SARS-CoV-2^+^ epithelial cells showing the cells that are significantly co-located with them within a 20μm radius [g(r) > 1], represented by the connecting lines, as determined by the cross-pair correlation analysis (cross-PCF) in the SpOOx pipeline. **(D)** Network representation of the pairwise interacting cell types identified by Giotto in the late death lungs. Only enriched interactions, depicted by the red lines, are shown. Width of the edges indicates the strength of spatial enrichment. **(E)** Barplots showing protein expression changes (log2fc) in endothelial (top panel) and activated endothelial cells (bottom panel) based on their spatial interaction with SARS-CoV-2^+^ alveolar macrophages (SARSCoV2^+^ AM) in the early death lungs. Differential expression is determined by a spatial permutation test followed by adjust for Bonferroni multiple hypothesis testing using a background null distribution reshuffling the cells within the same cell type. Black bars represent p-adjusted values < 0.05; grey bars represent p-adjusted values ≥ 0.05. **(F)** Barplots showing protein expression changes (log2fc) in endothelial (top panel) and activated endothelial cells (bottom panel) based on their spatial interaction with CD66b^High^ neutrophils in the early death (left) and late death (left) lungs. Differential expression is determined by a spatial permutation test followed by adjust for Bonferroni multiple hypothesis testing using a background null distribution reshuffling the cells within the same cell type. Black bars represent p-adjusted values < 0.05; grey bars represent p-adjusted values ≥ 0.05.

**Figure S9:**
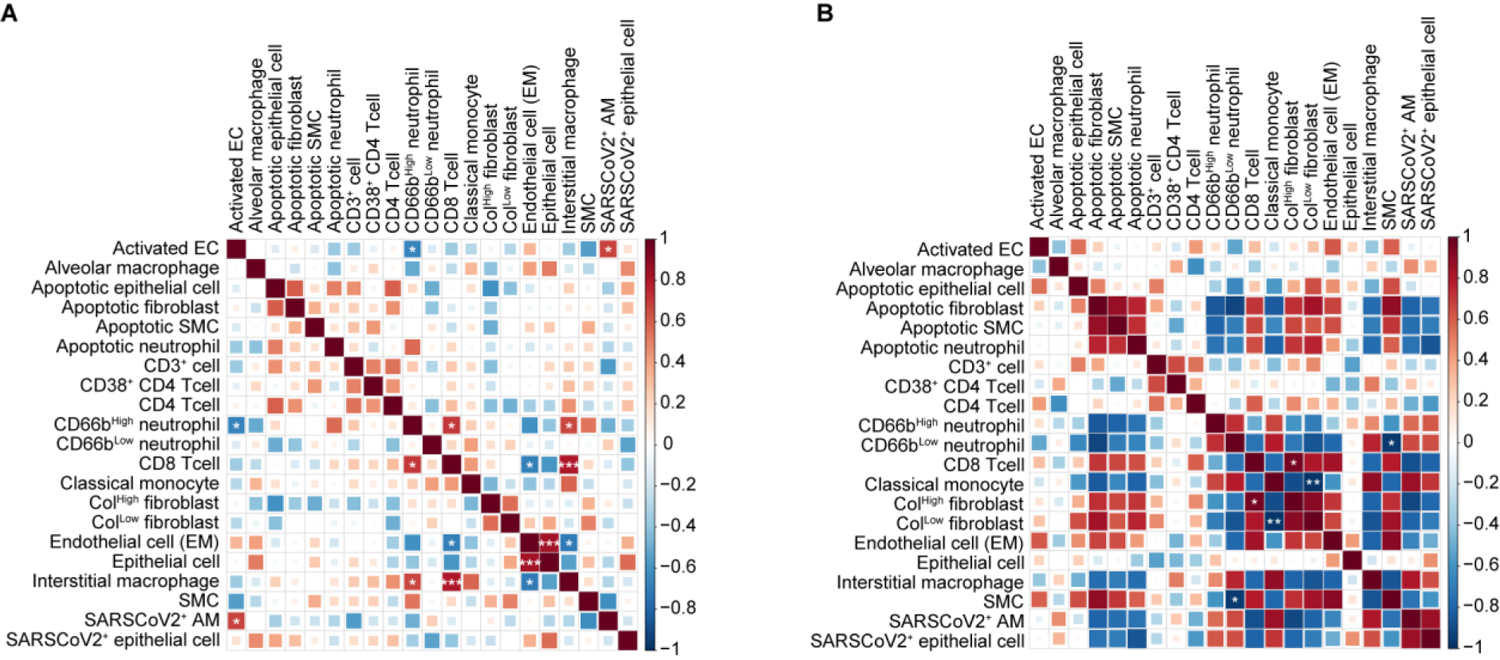
Correlation analysis between IMC data and other approaches. **(A)** Matrix plot of the Spearman’s rank correlations between the frequencies of the cell types identified by IMC in the post-mortem lung samples from COVID-19 patients following the early death progression. Asterisks represent the significant correlations (*p<0.05, **p<0.01, ***p<0.001). **(B)** Matrix plot of the Spearman’s rank correlations between the frequencies of the cell types identified by IMC in the post-mortem lung samples from COVID-19 patients following the late death progression (right panel). Asterisks represent the significant correlations (*p<0.05, **p<0.01, ***p<0.001).

**Figure S10:**
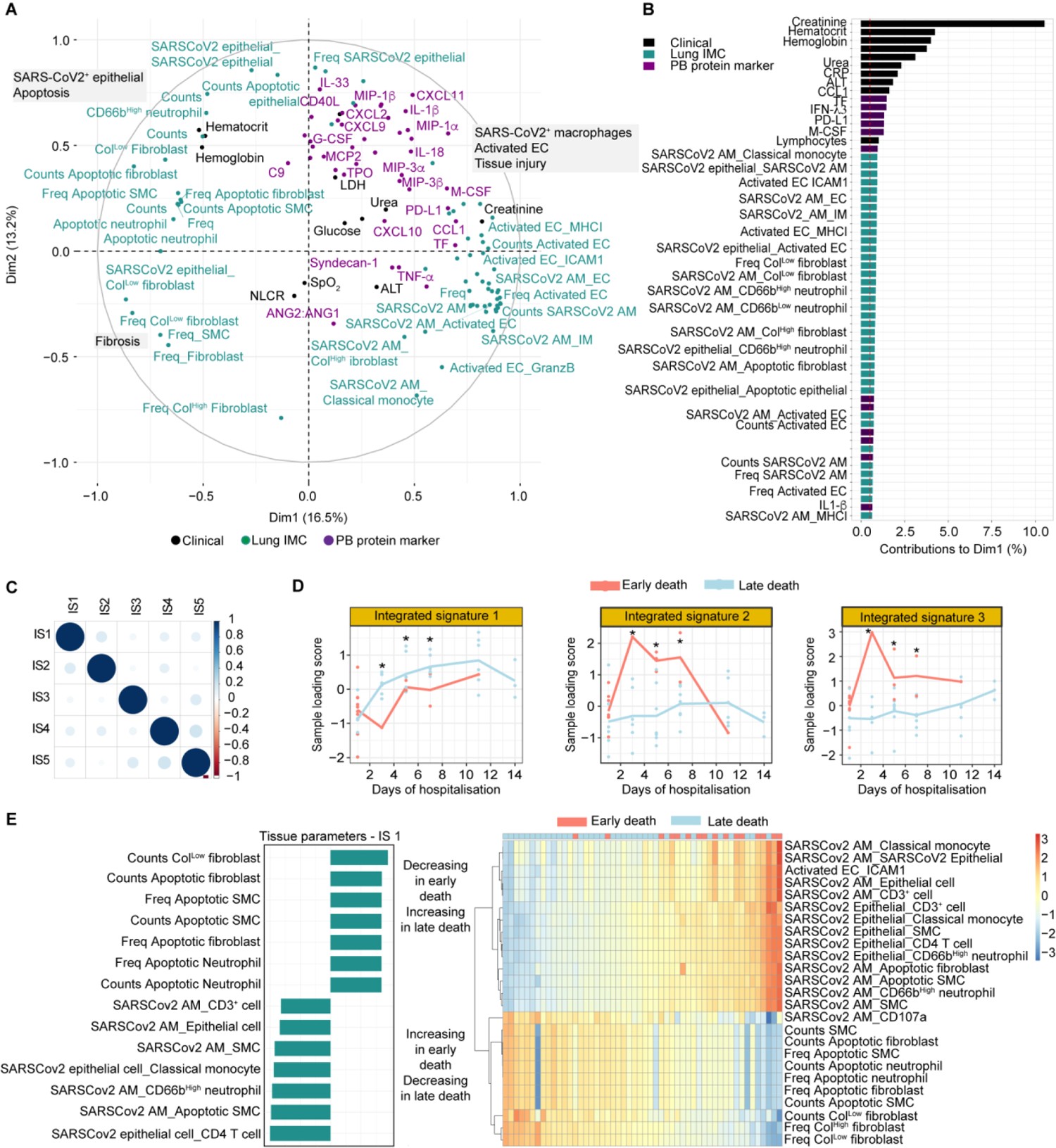
Integration of systemic and tissue signatures reveal the mechanisms driving distinct trajectories in disease progressions in fatal COVID-19 patients. **(A)** Systemic and tissue signatures defining the distribution of the early and late death patients in the MFA plot. In the dimensionality reduction plot, the underlying clinical parameters (in black), PB biomarkers (in purple) and tissue features (in green) driving the early and late fatal progression in this cohort are indicated. Freq = frequency; the underscore sign “_” between cell type labels indicates interacting cell pairs; the underscore sign “_” between a cell type label and a protein marker indicates protein expression in the corresponding cell type. **(B)** Variables contribution to dimensions 1 (Dim1) and 2 (Dim2) in the MFA. Contributions (in %), sorted in descending order of importance, of the tissue (green bars), clinical (black bars) and PB biomarkers (purple bars) on the variability retained by the dimension 1 (top panel) and by the dimension 2 (bottom panel) of the MFA are represented. Parameters contributing to the variance in the first dimension are the main determining factors in segregating the patients following the early death progression from those following the late death progression. Meanwhile, the parameters contributing to the variance in the second dimension, segregate the patient samples according to their day of sampling or day of symptoms. Freq = frequency; the underscore sign “_” between cell type labels indicates interacting cell pairs; the underscore sign “_” between a cell type label and a protein marker indicates protein expression in the corresponding cell type. **(C)** Correlation between the integrated signatures extracted from MOFA/MEFISTO analysis. The analysis of correlation between the integrated signatures (IS) is a good sanity check to confirm that there is a good enough number of factors, they are largely uncorrelated, the model has a good fit, and the normalisation is adequate. **(D)** MOFA factors (integrated signatures) values along the days of sampling (hospitalization). Sample loading scores for each integrated signature along the days of sampling (during hospitalization). The loading scores of each patient sample (dots) for each integrated signature were plotted against the days of sampling in line plots. Lines represent the averaged (within disease progression group) longitudinal trajectory of each integrated signature during hospitalization (early death in light red and late death in light blue). Dots represent loading scores of each patient sample. Significance was tested using unpaired t-test with multiple comparisons adjusted by false-discovery rate (FDR) calculated by the two-stage step-up method (Benjamini, Krieger and Yekutjeli). *q-value < 0.05, ** q-value < 0.01, *** q-value < 0.001. **(E)** Inspection of the top tissue features (from IMC) associated with the integrated signatures 1.

**Figure S11:**
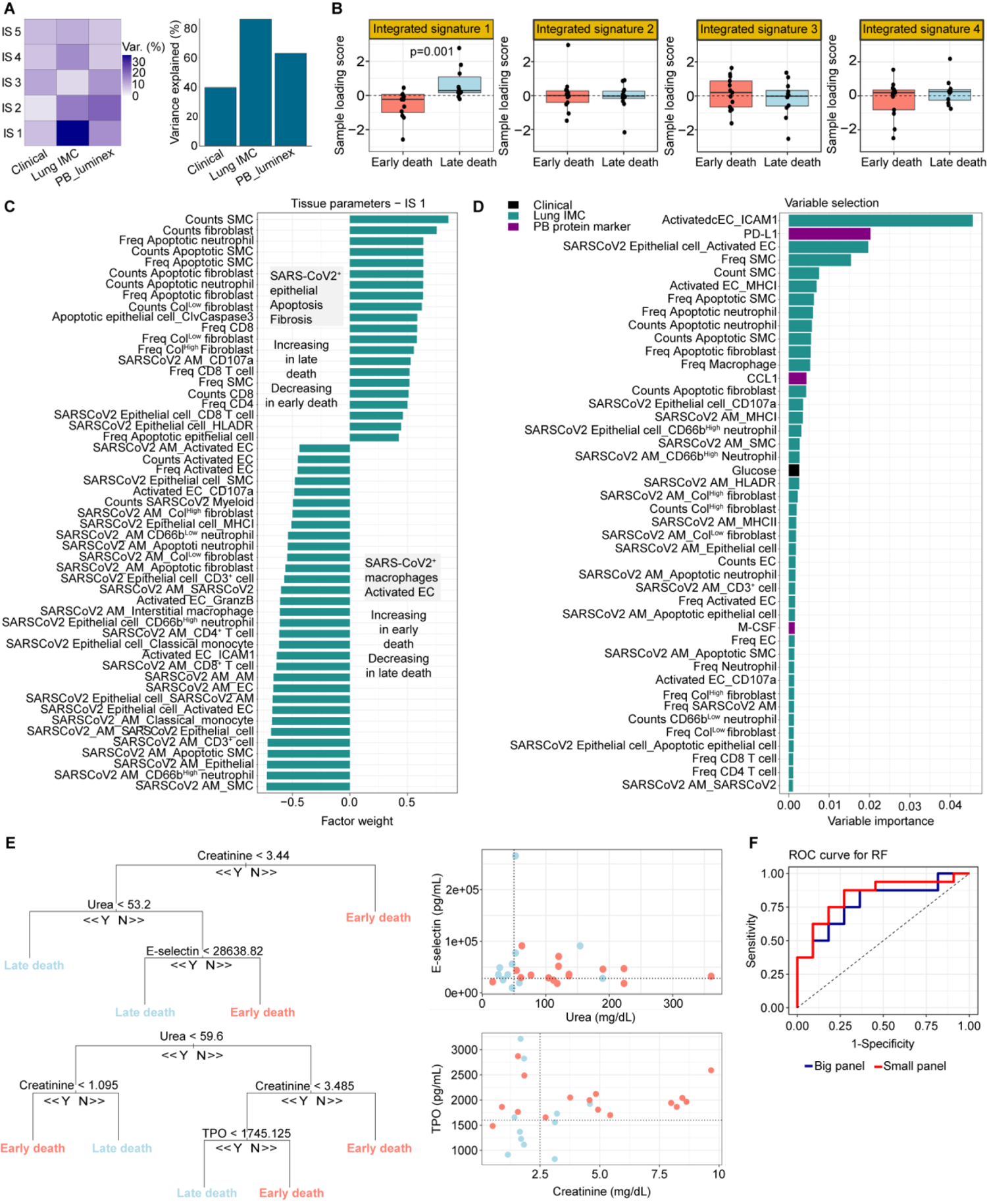
Integration of signatures in the Amazonian and north American COVID-19 fatal cohorts with MOFA and validation of the predictive potential of the integration signatures in the Amazonian fatal COVID-19. **(A)** MOFA quality control metrics. The further validate our data, MOFA/MEFISTO algorithms were applied to integrate the clinical, PB biomarker and tissue features (extracted from IMC) from the early and late death patients in the Amazonian and in a north American COVID-19 fatal cohort. Because the north American is a cross-sectional cohort, in this analysis, only clinical parameters and PB protein markers data from the last sample collected before time of death from patients in the Amazonian fatal cohort were used. Tissue features, extracted from IMC in both cohorts, were also used. The left panel represents the variance decomposition by integrated signature (IS), summarizing the sources of variation by showing the percentage of variance explained by each IS across each data modality (clinical, PB biomarker and IMC). A high variance (%) implies that the MOFA factors (here called integrated signatures) capture most of the variation for a specific approach, whereas small values means that the variation is not explained by the model (i.e. it is considered as noise). The right panel represents the total variance explained per approach, and it shows the total variance captured by all IS from each data source approach. **(B)** Integrated signatures derived from the integration of the Amazonian and north American (Rendeiro et al., 2021) COVID-19 fatal cohort clinical, PB and lung data. The loading scores of each patient sample (dots) for each integrated signature (IS) were plotted. The bars represent the median values of samples loading score and the error bars represent the interquartile range. Student’s t-test was used to compare medians between normally distributed data, and data sets with non-normal distributions were compared using Mann–Whitney test. All tests were performed two-sided using a nominal significance threshold of p<0.05 (when significant, the p-value is stated in the plot). **(C)** Inspection of the top lung tissue features associated with integrated signature 1, predicted to be driving most of the variance underlying the early and late death progression in both Amazonian and north American cohorts. The top lung tissue features with positive weights in the integrated signature 1 are enriched in the late death group and reduced in the early death progression and vice-versa. Freq = frequency; the underscore sign “_” between cell type labels indicates interacting cell pairs; the underscore sign “_” between a cell type label and a protein marker indicates protein expression in the corresponding cell type. **(D)** Random forests (RF) variable selection cross-validate the integrated signatures driving disease progression to the fatal outcome in the Amazonian COVID-19 cohort. To orthogonally validate the MOFA/MEFISTO and survival analysis findings, other systems biology integration approaches were applied, such as machine learning (ML) using random forests (RF). The same set of clinical (black bars), PB biomarker (purple bars) and tissue features (green bars) used in the MOFA/MEFISTO analysis were ranked by applying a feature selection method for building predictive models using random forests (RF)-based algorithms. Clinical parameters, PB biomarkers and tissue features are ordered in descending order of their importance based on mean decreased accuracy or OOR error rate in RF model predicting disease progression. The output of the feature selection step showed that the most important features predicting disease progression in our cohort are tissue features (green bars) in the integrated signature 2, and PB biomarkers in the PB signature 1. Freq = frequency; the underscore sign “_” between cell type labels indicates interacting cell pairs; the underscore sign “_” between a cell type label and a protein marker indicates protein expression in the corresponding cell type. **(E)** Example decision trees based on the top 3 clinical and top 3 PB biomarkers (small panel) in the integrated signature 2, measured up to 3 days of hospitalization, in predicting COVID-19 progression in hospitalized patients in the Amazonian COVID-19 cohort. To evaluate the practical value of the top 3 clinical parameters and top 3 PB biomarkers in the integrated signature 2 to predict disease progression when measured at or early after hospital admission, the RF model was trained in 70% of the dataset and then evaluated in a test dataset, using only the clinical and PB biomarker predictors measured at and up to 3 days of hospitalization. The decision trees and cut-off values for specific clinical and PB biomarker parameters measured up to 3 days of hospitalization exemplify how these parameters could assist in prediction and patient stratification in the clinical setting with the aim to evaluate the best therapeutic strategies. Cut-off values of the attribute that best divided groups were placed in the root of the tree according to the parameter value. Scatter plots (right panels) representing how the cut-off values of the paired parameters, measured up to 3 days of hospitalization, can separate patients based on the predicted disease progression. **(F)** Random forests (RF) model performance. Area under the receiver operating characteristic curves (AUROC) representing the performance of the RF models trained for prediction of disease progression. Models were trained and tested using the top 10 clinical and top 10 PB biomarkers in the integrated signature 2 (big panel – blue curve) or the top 3 clinical and top 3 PB biomarkers in the integrated signature 2 (small panel – red curve).

**Figure S12:**
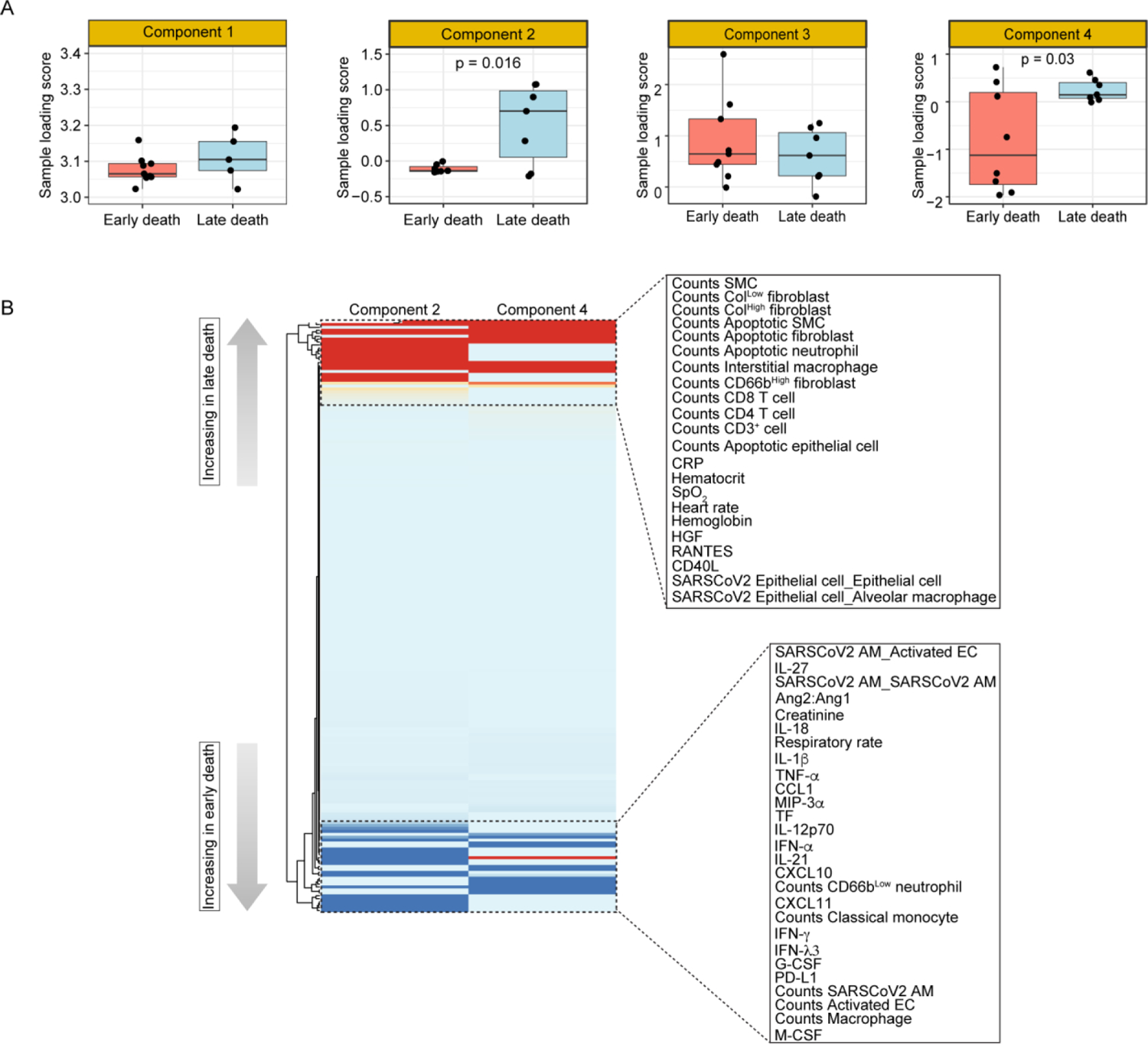
Integration of peripheral and tissue signatures in COVID-19 fatal patients using Tensor and matrix decomposition into latent SDA components further corroborates the mechanisms of COVID-19 progression to fatal outcome. **(A)** Sample loading scores per each latent SDA component. Tensor and matrix decomposition approach was also used to integrate the datasets originated from the different sources of data (clinical, PB and post-mortem lung). Components derived from the integration of clinical parameters, PB biomarkers and tissue features from the Amazonian COVID-19 fatal cohort. Dots represent the sample loading score for each patient in each component. The bars represent the median values and error bars represent the minimum-maximum range. Student’s t-test was used to compare medians between normally distributed data, and data sets with non-normal distributions were compared using Mann–Whitney test. All tests were performed two-sided using a nominal significance threshold of p<0.05 (when significant, the p-value is stated in the plot). **(B)** Scaled loading scores of clinical parameters, PB biomarker and post-mortem lung tissue features in the SDA components 2 and 4 were clustered and represented in a heatmap. The heatmap details the top clinical parameters, PB biomarker and tissue features with high posterior inclusion whose variance contributes to the components 2 and 4, identified as significantly driving variance between early and late death progression. Positive factor weights represented in red, are upregulated, while negative factor weights, in blue, are downregulated in the late death progression. The opposite is true for the early death progression. Freq = frequency; the underscore sign “_” between cell type labels indicates interacting cell pairs; the underscore sign “_” between a cell type label and a protein marker indicates protein expression in the corresponding cell type.

